# Genetic evidence for T-wave area from 12-lead electrocardiograms to monitor cardiovascular diseases in patients taking diabetes medications

**DOI:** 10.1101/2023.08.30.23294832

**Authors:** Mengling Qi, Haoyang Zhang, Xuehao Xiu, Dan He, David N. Cooper, Yuanhao Yang, Huiying Zhao

## Abstract

**Aims:** Many studies indicated use of diabetes medications can influence the electrocardiogram (ECG), which remains the simplest and fastest tool for assessing cardiac functions. However, few studies have explored the role of genetic factors in determining the relationship between the use of diabetes medications and ECG trace characteristics (ETC).

**Methods:** Genome-wide association studies (GWAS) were performed for 168 ETCs extracted from the 12-lead ECGs of 42,340 Europeans in the UK Biobank. The genetic correlations, causal relationships, and phenotypic relationships of these ETCs with medication usage, as well as the risk of cardiovascular diseases (CVDs), were estimated by linkage disequilibrium score regression (LDSC), Mendelian randomization (MR), and regression model, respectively.

**Results:** The GWAS identified 124 independent single nucleotide polymorphisms (SNPs) that were study-wise and genome-wide significantly associated with at least one ETC. Regression model and LDSC identified significant phenotypic and genetic correlations of T-wave area in lead aVR (aVR_T-area) with usage of diabetes medications (ATC code: A10 drugs, and metformin), and the risks of ischemic heart disease (IHD) and coronary atherosclerosis (CA). MR analyses support a putative causal effect of the use of diabetes medications on decreasing aVR_T-area, and on increasing risk of IHD and CA.

**Conclusion:** Patients taking diabetes medications are prone to have decreased aVR_T-area and an increased risk of IHD and CA. The aVR_T-area is therefore a potential ECG marker for pre-clinical prediction of IHD and CA in patients taking diabetes medications.

## Introduction

It has been blithely assumed that glycemic control would be beneficial in reducing the risk of cardiovascular disease (CVD). To this end, a large number of clinical trials have investigated the potential use of diabetes medications in improving cardiac health. These studies have demonstrated that many diabetes medications are not associated with a reduction in the rate of heart failure(1, 2, 3, 4), and that rosiglitazone and pioglitazone are actually associated with increasing the risk of heart failure(5, 6). The effects of the commonly used first line therapy, metformin, on cardiovascular end points remain unclear(7).

Heart functions are usually measured by the electrocardiogram (ECG). The ECG provides an electrical tracing of the heart that is recorded non-invasively by placing electrodes on the chest, arms and legs. It is the simplest and fastest clinical method available to assess cardiac abnormalities(8). A greater dispersion of QT intervals has been reported to be related to a high dose of metformin (9) whereas the use of metformin has been found to be associated with a reduced heart rate(10). However, the challenge remains as to how to identify ECG markers associated with the use of diabetes medications due to our lack of knowledge of the shared genetic landscape between them.

Recently, genome-wide association studies (GWAS) have been performed to identify variants associated with the traditional ECG trace characteristics (ETCs) known to represent risk factors for particular cardiac disorders. Importantly, the advent of the UK Biobank (UKB), which collated genotype data from a large population, allowed us to perform large-scale GWAS for less conventional ETCs, such as wave-related ETCs. The most recent GWAS on the use of diabetes medications (11), and a recently established FinnGen Biobank, included GWAS for various cardiac disorders offering us an opportunity to investigate the genetic relationships between ETCs, diabetes medication use and CVDs, which is important for identifying key ETCs of use in monitoring heart functions in patients using diabetes medications.

In this study, GWAS were performed to identify genetic variants associated with ETCs, and further detect their phenotypic and genetic relationships with diabetes medication use and CVDs. The phenotypic and genetic relationships of these ETCs with medication use and CVDs were investigated by regression models, LDSC(12) and seven different MR approaches(13, 14, 15, 16, 17, 18, 19). This study provides genetic evidence to support the use of ECG in monitoring CVDs in patients taking diabetes medications.

## Methods and Materials

### Electrocardiogram morphology phenotypes and quality control

The 12-lead electrocardiogram (ECG) records of 42,340 Europeans were downloaded from the UK-Biobank (UKB). The ECG signal from each sample was a 10-second 12-lead signal with a sampling rate of 500 Hz. The preprocessing and feature extraction of ECG can be found in the supplementary materials.

### The phenotypic relationships between traits

Regression models and the difference test were used to examine the phenotypic relationships between ETCs, diabetes medication use, and cardiovascular diseases through medical records in the UKB. The regression models were constructed using 38,953 samples from the UKB, after removing samples with missing information, which were adjusted by age, sex, diabetes, genotype batch and assessment centre.

### Genome-wide association studies for ETCs

A genome-wide association study (GWAS) was performed for each ETC of European ancestry using UKB data. European ancestry was selected by a two-stage approach as described in Supplementary Material. The GWAS analysis was carried out by BOLT-LMM(20, which is a Bayesian-based linear mixed model (LMM) that makes allowance for the relatedness of individuals by adjusting the population structure and cryptic relatedness. BOLT-LMM was corrected for age, sex, genotype batch and assessment center, and was fitted by random effects of the restricted single nucleotide polymorphisms (SNPs) (with LD *r*^2^ < 0.9). The SNPs used in BOLT-LMM were calibrated by the LD scores calculated from the 1000 Genomes Project Europeans reference(21). Genotyped SNPs were imputed according to the Haplotype Reference Consortium reference panel(22). SNPs with minor allele frequency (MAF) < 0.01, imputation INFO score < 0.3, P-value of Hardy-Weinberg test < 1×10^-6^, minor allele counts < 5, or the call rate < 0.05 were excluded, yielding a final set of 8.54M SNPs.

### Causal relationship inference

For the pairs of phenotypes with significant genetic correlations, Mendelian randomization (MR) analysis was performed to detect their putative causal relationships through multiple MR models (inverse-variance-weighted (IVW), MR-Egger, weighted mode, weighed median, generalized summary-data-based Mendelian randomization (GSMR), causal analysis using summary effect estimates (CAUSE), and MRlap). MRlap(23) was employed to correct the sample overlap and is a method to approximate the sample overlap using cross-trait LD-score regression (LDSC). It has been used in many studies for MR analysis(24, 25, 26, 27, 28). The detailed descriptions of all the MR approaches are given in the Supplementary Material.

The putative causal relationship between two traits was considered to be determined if consistent results were identified by more than four MR models with FDR < 0.05. Depending on the data type of the outcome and exposure trait, the MR estimates (i.e., *β_xy_*) were converted to liability scale using one of the methods described by Byrne et al.(29, 30). The related formulae of the methods were described in the Supplementary Material.

## Results

### Overview of this study

Figure 1 depicts the overall workflow of this study. The study comprises three modules: (1) extracting 168 ETCs from 42,340 UKB samples with 12-lead ECG data, and performing genome-wide association studies (GWAS) for each ETC; (2) phenotypic relationship analysis of the ETCs, cardiovascular diseases and diabetes medication use by leveraging the electronic medical records of samples in UKB; (3) employing GWAS summary data of the ETCs, diabetes medication use and CVDs to investigate the genetic correlations and genetic causal links between them by LDSC and MR analysis. The GWAS data sources used in this study were described in Supplementary Material.

**Figure 1.**
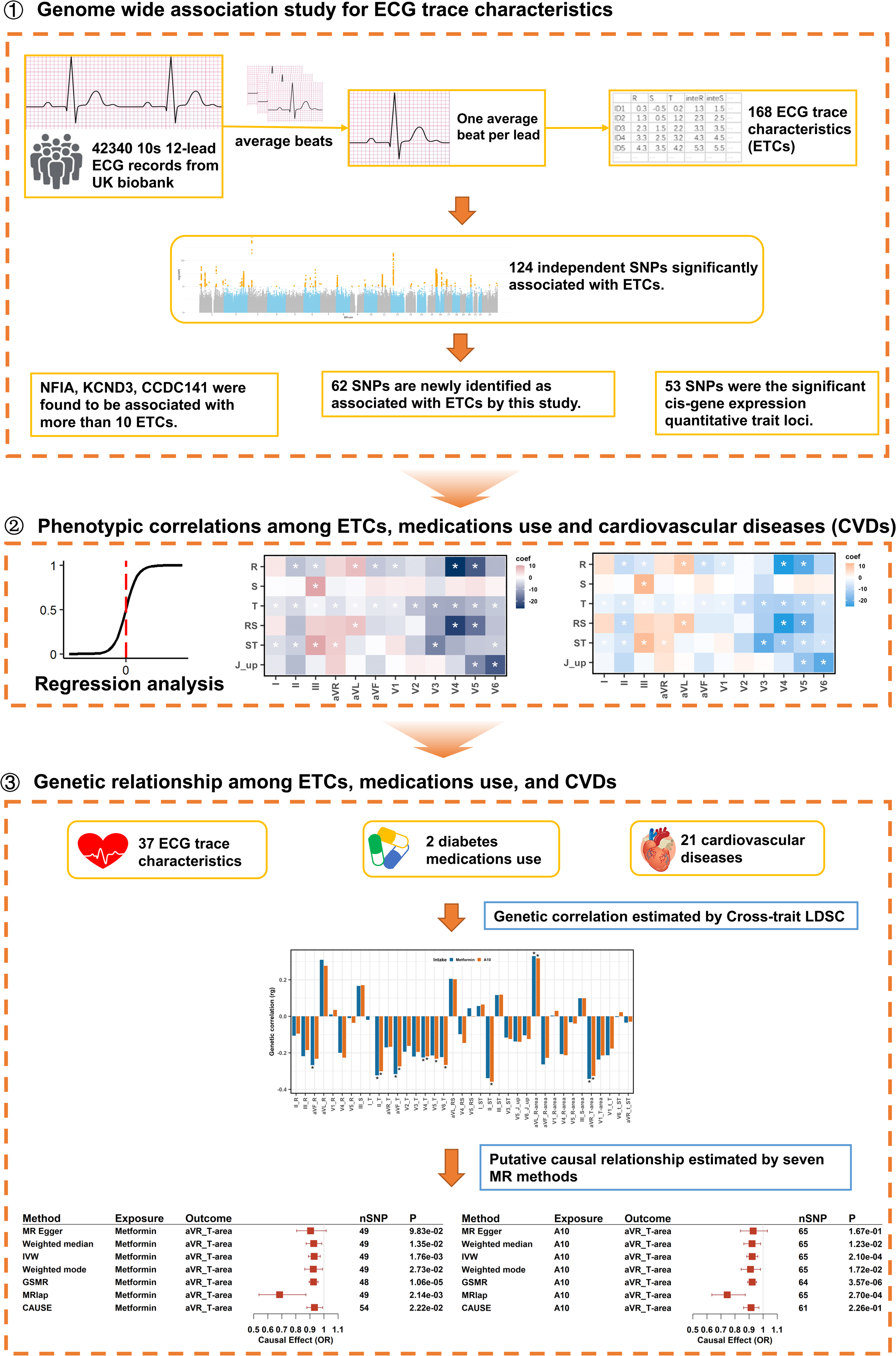
Schematics for genetic and phenotypic analysis of relationships between ETCs, medication use, and CVDs. Step 1: Genome-wide association study for 168 ETCs by BLOT-LMM; Step 2: Analyzing phenotypic relationships among ETCs, taking A10/metformin and cardiovascular diseases, by employing medical records from the UK-Biobank; Step 3: Analyzing genetic relationships between the ETCs, medication usage and CVDs through LDSC and seven MR methods.

### Genome-wide association study of ETCs

The descriptions and short names of all the 168 ETCs are given in Supplementary Table 2. Through the GWAS for these 168 ETCs, 124 independent SNPs were identified as study-wise and genome-wide significantly (*P-value*<5×10^-8^/168, LD *r^2^*<0.05) associated with at least one ETC (Figure 2, Supplementary Table 4). These 124 SNPs were systematically analyzed by using both the extensive GWAS Catalog(31) and more recent phenome-wide associations (PheWeb) from UKB(32) (Supplementary Table 4). Out of the 124 loci, 45 have been reported to be associated with at least one type of heart function, such as electrocardiogram morphology, QRS duration, left ventricular structure and function (Supplementary Table 4), 38 loci were found by previous studies to be associated with CVDs, such as atrial fibrillation and flutter, atrioventricular block and heart failure, and 53 SNPs were significant *cis*-gene expression quantitative trait loci (*cis*-eQTLs)(33)(Supplementary Table 4). These *cis*-eQTLs were found to regulate genes enriched in phospholipid binding, phosphatidylinositol-5-phosphate binding, transmembrane receptor protein kinase activity, and phosphatidylinositol-4-phosphate binding functions (P-value<0.01, Supplementary Figure 1) according to clusterProfiler analysis (34). In total, 62 SNPs had not been previously reported as being associated with any ETCs or heart disease (Figure 2A).

**Figure 2.**
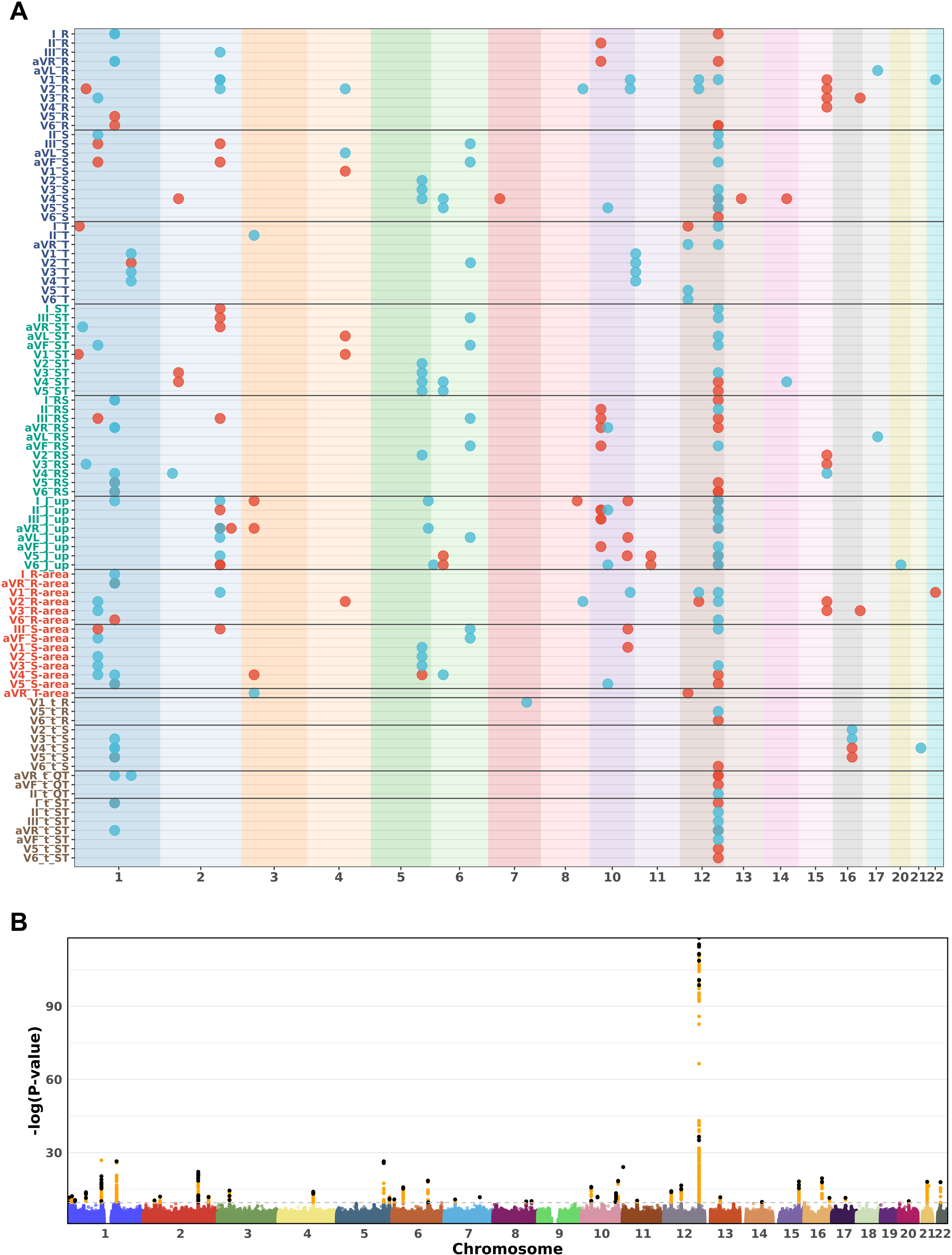
Genome-wide association studies for the ETCs. A. GWAS identified 124 independent SNPs as being significantly associated with at least one ETC 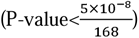), half of which have been reported to be associated with ECG or CVDs (red points), and 62 SNPs that were novel findings from this study (blue points). The blue text on the *Y* axis refers to the ETCs of R/S/T wave amplitudes in each lead; the green text on the *Y* axis refers to the ETCs of combinations of amplitudes; the red text on the *Y* axis denotes the ETCs of wave areas for R/S/T in each lead; the brown text on the *Y* axis refers to the ETCs of time interval in each lead. The background color represents different chromosomes. The chromosome index is shown on the *X* axis. B. Manhattan plot of the ETCs and the smallest P-value of each SNP across all the traits is shown. The red dashed line shows the threshold P-value = 5×10^-8^/168. Orange points represent SNPs with P-value <5×10^-8^ /168, whereas black points represent SNPs with LD *r^2^*<0.05 with at least one SNP within 1000 bp.

These 124 independent loci were assigned to 37 genes (Figure 2, Supplementary Table 4). The SNPs in *NFIA* (7 SNPs), *KCND3* (10 SNPs), *CCDC141* genes (6 SNPs), and 12q24.21 region (18 SNPs) were significantly associated with more than 10 ETCs, suggesting key roles for these genomic regions in characterizing ECG (Figure 2B, Supplementary Table 4). More detailed information on these SNPs is provided in the Supplementary Material and Supplementary Figure 2. Genes associated with each ETC, which were analyzed by gene-level GWAS are given in the Supplementary Material.

### Phenotypic association among ETCs, use of diabetes medication, and CVDs

The phenotypic relationship among ETCs, diabetes medication use, and CVDs were analyzed by means of difference test and regression model using ECG records of 38,953 samples (the sample sizes used for the phenotypic analysis are given in Supplementary Table 7). The regression analysis indicated 62 ETCs were significantly (Bonferroni adjusted P-value < 0.05) correlated with use of diabetes medication (A10) and metformin (Figure 3A-B). Among them, the ETCs related with R, T, ST, t_T, t_ST, and t_QT were significantly correlated with both A10 and metformin use in more than 6 leads. Detailed information is given in Supplementary Table 8. In addition, 6 ETCs were significantly (P-adjust < 0.05) correlated with metformin use only (Figure 3B). Further analysis indicated that 125 ETCs were significantly correlated with 10 CVDs (Bonferroni adjusted P-value < 0.05, Figure 3C). Additionally, coronary atherosclerosis, major coronary heart disease events, cardiomyopathy, cardiovascular diseases, heart failure and ischaemic heart disease were found to be significantly correlated with more than 50 ETCs.

**Figure 3.**
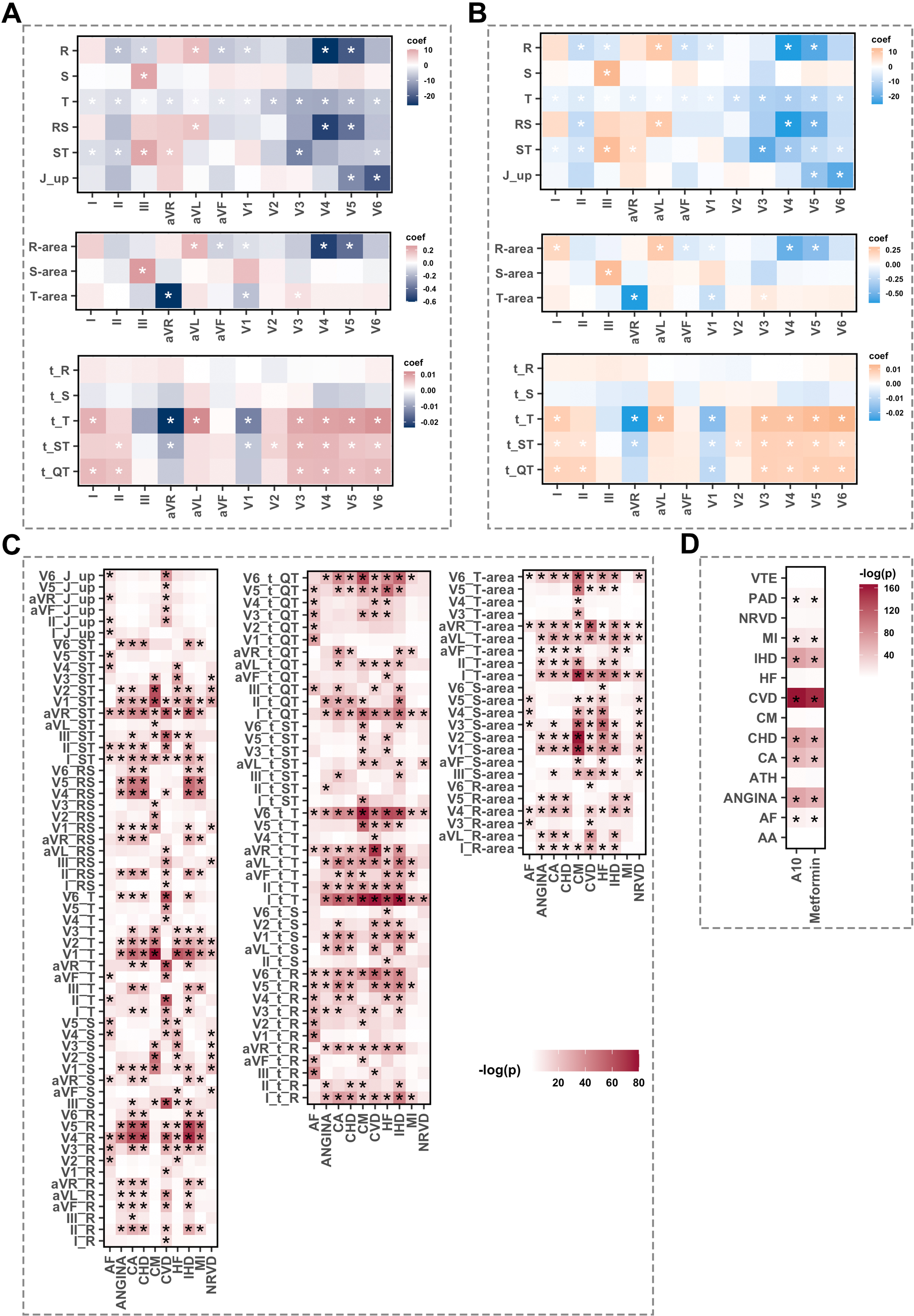
Phenotypic relationship among ETCs, A10/metformin use, and CVDs. A. The coefficients of the regression analysis between ETCs and A10 use. The markers on the horizontal axis denote the names of leads, whilst the labels on the vertical axis denote the ETCs. The darker colors represent higher values of coefficients *(beta)*. B. Coefficients of the regression analysis between ETCs and metformin use. The markers on the horizontal axis are the lead name, the labels on the vertical axis are the ETCs. The darker colors represent higher values of coefficients *(beta)*. C. Relationships between ETCs and CVDs. The markers on the horizontal axis are the abbreviations of CVDs, whilst the labels on the vertical axis are the ETCs. The darker colors represent lower P-values. D. Relationships between medicine use and CVDs. The markers on the horizontal axis denote medicine use, the labels on the vertical axis are the CVDs. The darker colors represent lower P-values. “*” denotes Bonferroni adjusted P-value < 0.05.

When we analysed the correlations between the use of diabetes medications and CVDs, we found 8 CVDs (atrial fibrillation and flutter, angina pectoris, coronary atherosclerosis, major coronary heart disease event, cardiovascular diseases, ischaemic heart disease, myocardial infarction, peripheral artery disease) were significantly (Bonferroni adjusted P-value < 0.05) correlated with A10 use and metformin use (Figure 3D).

### Genetic heritability and genetic correlations of ETCs, use of diabetes medications, and cardiovascular diseases

The methods for estimation of genetic heritability and genetic correlations were described in Supplementary Material. The SNP-based heritability (*h^2^*) of the 168 ETCs was estimated using single-trait LDSC (Figure 4A, Supplementary Table 9). Of 168 ETCs, 143 showed an FDR significance (FDR<0.05) and non-zero heritability, ranging from 0.03 to 0.19. The heritability of 101 ETCs also reached the Bonferroni-corrected level of significance (P-values <0.05/168; Figure 4A). The Liability-scale heritability (*h*^2^) of A10 and metformin use were 0.046 (P= 5.88 × 10^-52^) and 0.030 (P= 3.68 × 10^-56^), respectively. The heritability of cardiovascular diseases (CVDs) was estimated using 22 GWAS summary statistics of CVDs from FinnGen (Supplementary Table 1). The Liability-scale *h*^2^ for these CVDs ranged from 0.015 to 0.088 and P < 0.05/22 for 21 CVDs (Figure 4B, Supplementary Table 10).

**Figure 4.**
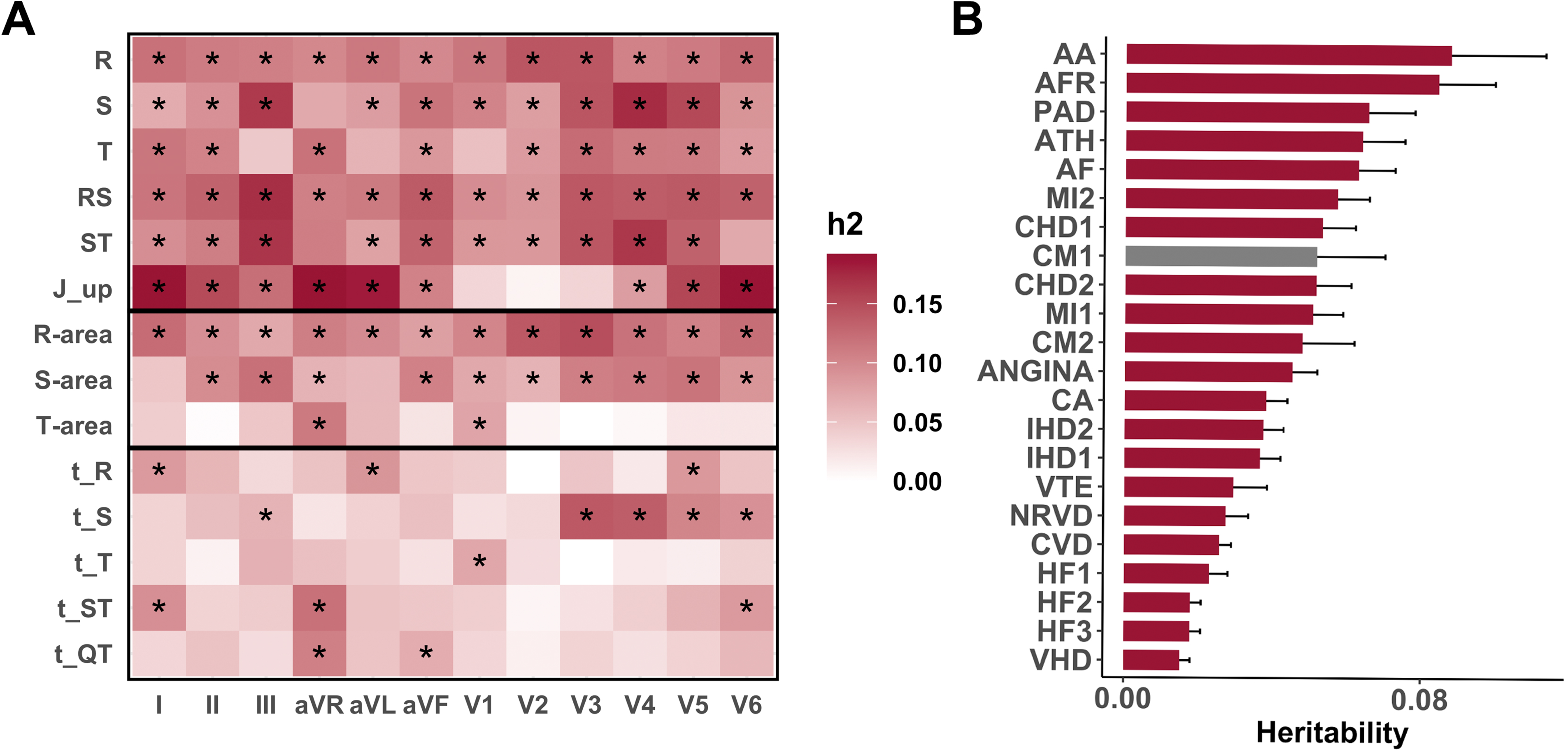
The heritability (*h^2^*) of ETCs and CVDs. A. The heritability of each ETC from different leads, estimated by LDSC with unconstrained intercept. The markers on the horizontal axis are the lead name, the labels on the vertical axis are the ETCs. The darker colors represent higher values of *h^2^*. “*” denotes a P-value < 0.05/168. B. Heritability of 22 CVDs was calculated by single-trait LDSC. Red bars denote P-values for heritability estimation lower than 0.05/22. The labels on the vertical axis are the Abbreviations which can be checked in Supplementary Table 1.

From 62 ETCs that were found to be significantly correlated with A10 and metformin use in phenotypic analysis, we found 37 had significant heritability (Bonferroni corrected P-adjust < 0.05, *h^2^* > 0). We firstly used the cross-trait LDSC analysis to estimate the genetic correlations between these 37 ETCs and the use of diabetes medications (Figure 5A). As shown in Figure 5A, the use of A10 was found to be significantly correlated (Bonferroni corrected P-adjust < 0.05) with 8 ETCs. Among these ETCs, five (II_T, aVF_T, V4_T, aVL_R-area, and aVR_T-area) were also genetically correlated with metformin use. Detailed information is given in Supplementary Table 11.

**Figure 5.**
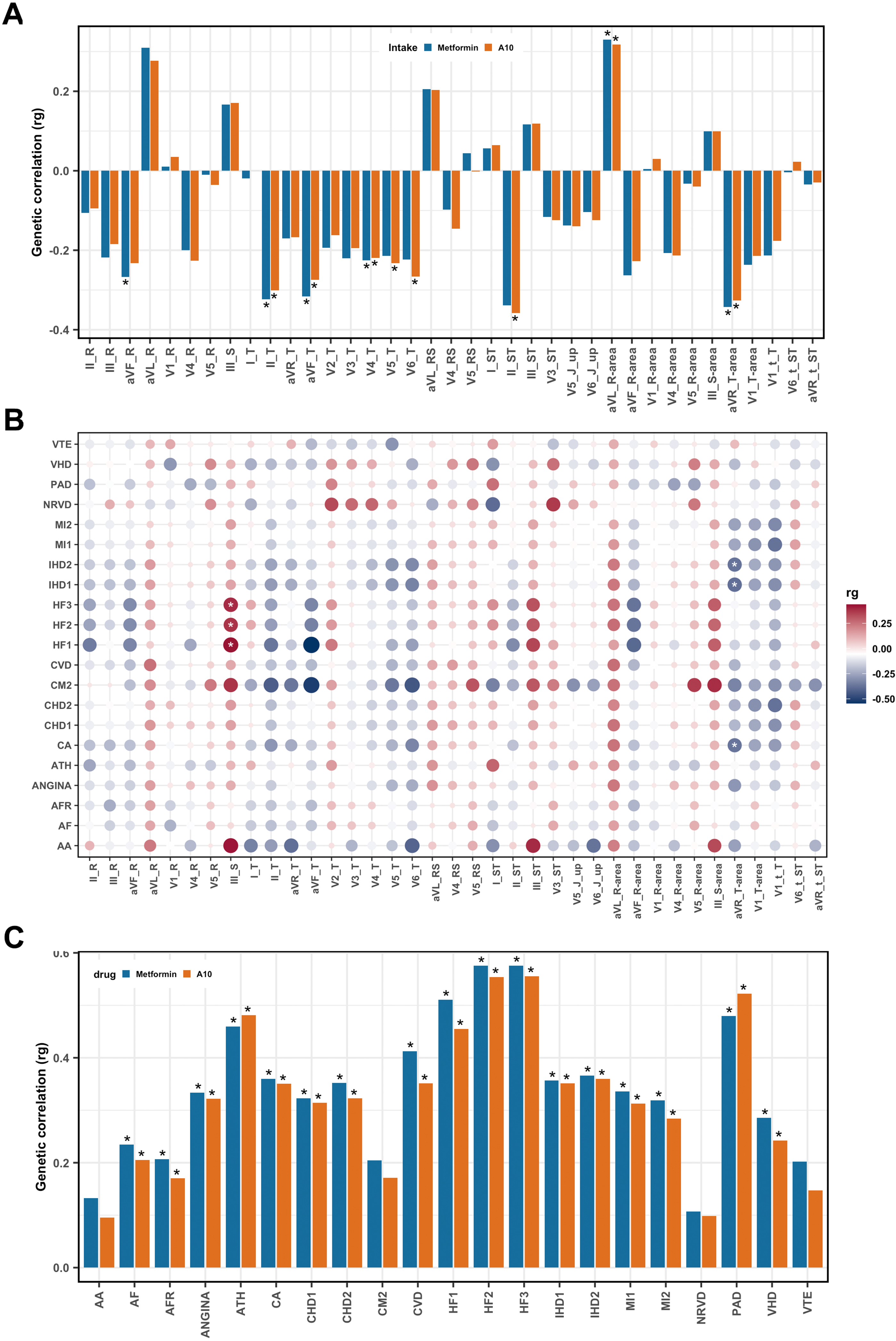
Genetic correlations among ETCs, medication use and CVDs. A. Genetic correlation (*r_g_*) between ETCs and medication use. Blue and red bars represent metformin use and A10 use respectively. “*” denotes a Bonferroni corrected P-value < 0.05. B. LDSC analysis was used to calculate genetic correlation (*r_g_*) between ETCs and CVDs. The larger and darker dots represent higher *r_g_*. Blue and red represent negative and positive correlations respectively. “*” denotes a FDR < 0.05. C. Genetic correlation (*r_g_*) between medication use and CVDs by LDSC analysis. The larger and darker dots represent higher absolute *r_g_*. Blue and red colors represent negative and positive correlations respectively. “*” means Bonferroni corrected P-value < 0.05.

Next, the genetic correlation between the ETCs and cardiovascular diseases (CVDs) was examined using 21 GWAS summary statistics of CVDs from FinnGen (Supplementary Table 1). The genetic correlations between the 37 ETCs and 21 CVDs (Bonferroni corrected *P_heritability_* < 0.05/22) were analyzed by cross-trait LDSC, indicating that 6 ETC-CVD pairs had significant genetic correlations (absolute *r_g_* ranging from 0.37 to 0.44, FDR < 0.05) (Figure 5B, Supplementary Table 12). Among them, heart failure (HF) was significantly correlated with increased III_S whereas ischemic heart disease (IHD) and coronary atherosclerosis (CA) were found to be significantly correlated with decreased aVR_T-area. The phenotypic correlation analysis also indicated that HF is significantly (Bonferroni adjusted P-value = 0.009) correlated with increased III_S and IHD. Further, CA was found to be significantly (P-adjust = 2.34 × 10^-12^ and P-adjust = 6.74 × 10^-12^) correlated with decreased aVR_T-area by phenotypic correlation analysis (Figure 3C).

Finally, the genetic correlations between CVDs and the use of diabetes medication were estimated by cross-trait LDSC (Figure 5C), indicating 17 CVDs have significant genetic correlation with the use of both A10 and metformin (Bonferroni adjusted P<0.05, Supplementary Table 13). Among them, 8 CVDs were found to be significantly (P-adjust < 0.05) correlated with both A10 and metformin use by phenotypic analysis (Figure 3D).

### Mendelian Randomization analysis to identify putative causal relationships between ETCs, use of diabetes medications and CVDs

The putative causal relationship between the 14 ETCs and the use of diabetes medications that have shown significant phenotypic (Figure 3A-B) and genetic correlations (Figure 5A) was analyzed by means of seven different MR approaches (see Methods). Six out of seven MR models provided evidence supporting use of A10 has a putative casual effect on aVR_T-area with estimated liability-scale odds ratios (ORs) ranging from 0.74-0.92 (Figure 6) (weighted median: FDR = 0.018, IVW: FDR= 5.92 × 10^-4^, weighted mode: FDR=0.018, GSMR: FDR= 2.50 × 10^-5^, MRlap: FDR=5.92 × 10^-4^, and CAUSE: FDR=5.74 × 10^-3^). Furthermore, the use of metformin also had a putative causal effect (FDR < 0.05) on aVR_T-area as indicated by six out of seven MR methods (the exception being MR-Egger), with estimated liability-scale ORs ranging from 0.68-0.93 (Figure 6). These results suggested that those samples associated with the use of A10 or metformin were prone to have an approximately 12% smaller aVR_T-area as compared to individuals taking neither A10 or metformin.

**Figure 6.**
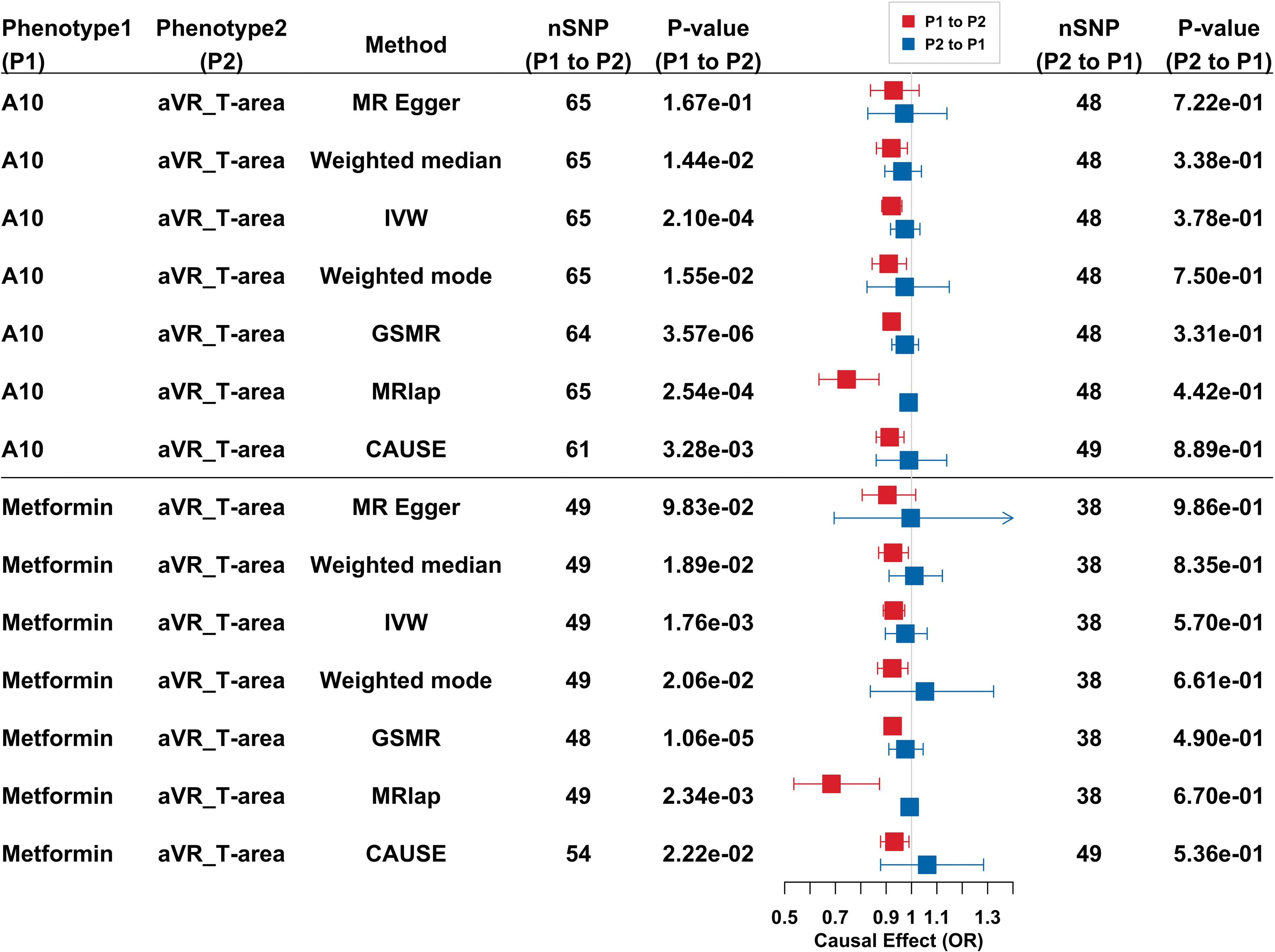
The causal relationship (in log-scale odds ratio [OR]) between A10 or metformin use and aVR_T-area was estimated by seven MR methods. The value of ORs and 95% confidence intervals (CIs) indicates the change of aVR_T-area for samples using A10 or metformin compared to the samples associated with never having taken these two medicines. CAUSE: causal analysis using summary effect estimates; GSMR: generalized summary-data-based Mendelian randomization; IVW: inverse variance weighted. Red and blue colors denote different directions of MR analysis.

When the MR methods were employed to identify putative causal relationships between the ETCs and CVDs, a causal effect (OR:1.02∼1.03) of III_S on HF was identified by five MR methods (Figure 7, Supplementary Table 14), suggesting that an increase of 1*uV* in III_S led to an ∼2% increase in the risk of HF. No putative causal effect was observed in the reverse direction by MR analysis.

**Figure 7.**
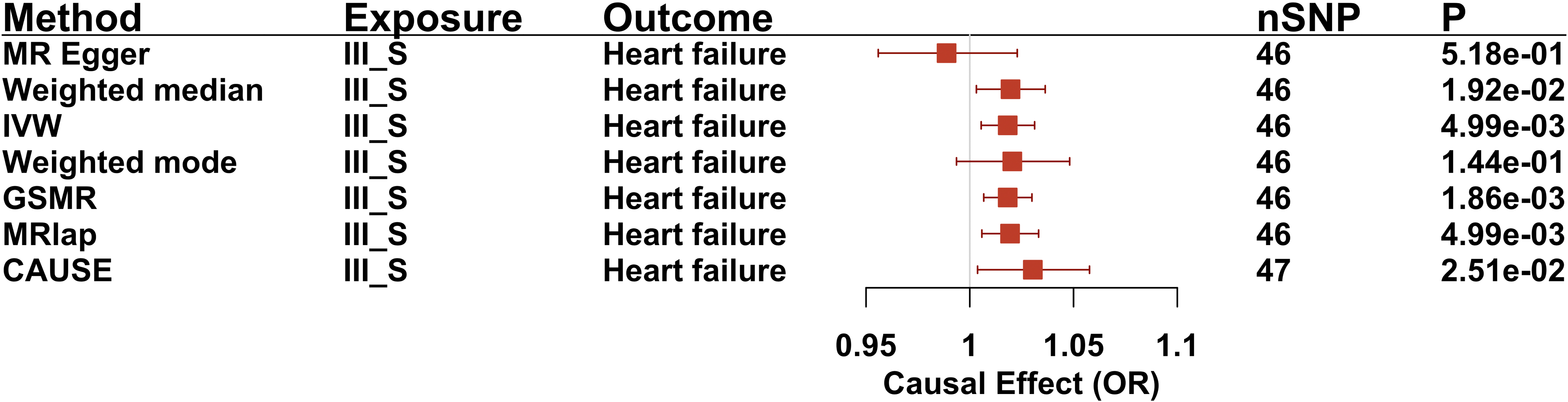
The causal relationship (in log-scale odds ratio [OR]) of III_S on HF was estimated by seven MR methods in two directions. Odds ratios (ORs) and 95% confidence intervals (CIs) express the risk of HF for the change of III_S compared to the samples without HF.

The causal relationship between CVDs and A10/metformin use was further explored. Specifically, the role of genetics to ascertain the impact of A10 on IHD risk was supported by six out of the seven MR methods (OR: 1.14-2.30; FDR = 3.60 × 10^-8^-2.54 × 10^-2^ for IHD1; OR: 1.13-2.14; FDR = 4.80 × 10^-7^-4.52× 10^-2^ for IHD2) (Figure 8, Supplementary Figure 6). Further, all seven MR methods provided consistent evidence in support of a causal effect of metformin use on IHD (OR: 1.13-2.14 for IHD1; OR: 1.11-2.66 for IHD2) (Figure 8, Supplementary Figure 6). In addition, the use of genetics to ascertain the impact of A10 or metformin use also indicated a putative causal effect on CA (Figure 8, OR: 1.11-2.66 for A10; OR: 1.13-3.85 for metformin). More results about the genetics of A10/metformin use and CVDs are summarized in Supplementary Table 15.

**Figure 8.**
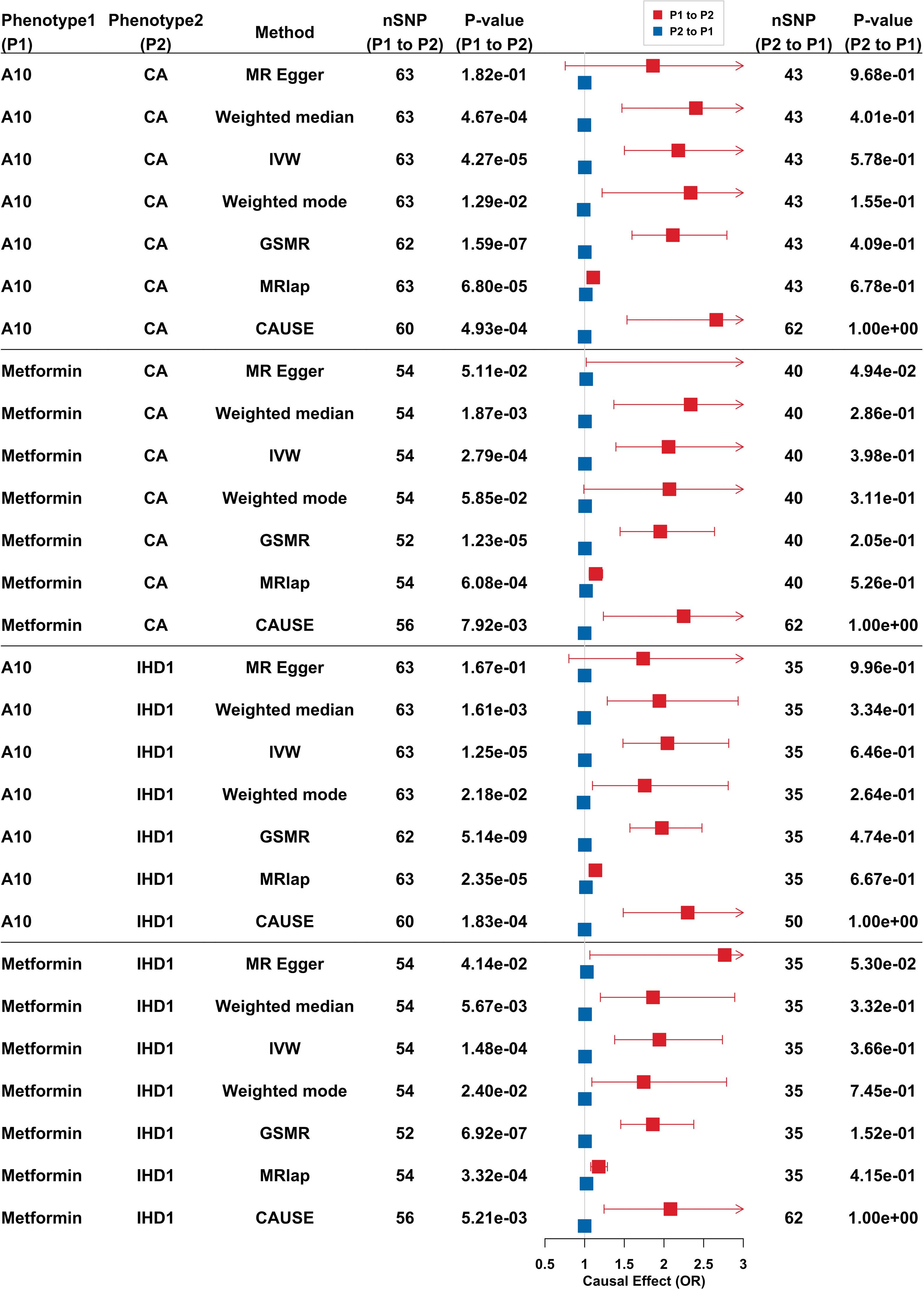
The causal relationship (in log-scale odds ratio [OR]) of A10 or metformin use on IHD or CA was estimated by seven MR methods. Odds ratios (ORs) and 95% confidence intervals (CIs) express the risk of IHD for samples associated with A10 or metformin use compared to the samples associated with never take these two medicines. IHD1: I9_ISCHHEART, ischemic heart disease; CA: I9_CORATHER, coronary atherosclerosis.

Taken together, the use of metformin or A10 has a putative causal effect on aVR_T-area, as well as on the risk of CA and IHD, whilst aVR_T-area exhibits a genetic correlation with IHD and CA (only *r_g_* was significant). In addition, the III_S has a putative causal effect on HF. These relationships provided genetic evidence in support of the results of the phenotypic analysis in Figure 3.

## Discussion

In this study, GWAS were performed for 12-lead ECG that was characterized by 168 ETCs, including not only the classical ETCs but also wave area-related features. The GWAS identified 124 independent SNPs associated with at least one ETC. Half of these have not been reported as being associated with CVDs or classical ETCs (Supplementary Material, Supplementary Table 5). Many variants near genes with calcium or potassium channel functions were identified as being associated with R-area (Supplementary Table 4) whereas variants near genes known to be related with autism spectrum disorder or schizophrenia were associated with S-area (Supplementary Table 4). Moreover, variants identified as being associated with T-area tended to be located near sodium channel genes (Supplementary Table 4). For example, *HAND1* and *SAP30L* were found to be associated with many S-wave related traits. *HAND1* is already known to be linked to developmental heart chamber defects(35) and SNPs near to *HAND1* have been reported to be associated with the QR-interval(36). Another gene, *KCND3,* was identified as being significantly associated with 29 ETCs. Although previous studies have demonstrated that *KCND3* is associated with early repolarization on electrocardiograms(37, 38), we provide further evidence to support a genetic association between *KCND3* with various ETCs, suggesting an important role for *KCND3* in cardiac function.

The heritability estimation of ETCs indicated that the highest heritability 0.192 (SE=0.027 and SE = 0.029) for the difference between R start and S end in voltage in aVR lead (aVR_J_up) and the difference between R start and S end in voltage in I lead (I_J_up) (Supplementary Table 7). The 10 highest heritability estimates of ETCs were all related to the ST segment (Supplementary Table 7). Interestingly, we found that the 10 ETCs with the lowest heritability included nine T-wave related features. Thus, T-wave related features may be characterized by lower heritability than features associated with other ECG waves.

A recent clinical study investigated the effects of long-term metformin use on heart disease outcome by following 3,234 participants for 21 years, and demonstrated that metformin had no impact on cardiovascular survival(39). Another randomized clinical trial in 368 samples investigating the effects of metformin on coronary heart disease indicated that taking metformin had no effect on cardiovascular disease(40). This study has provided the first genetic evidence that patients taking metformin (*r_g_* = -0.34 and P-value = 3.00 × 10^-4^) or A10 (*r_g_* = -0.33 and P-value = 8.82 × 10^-6^) tend to display a decreased aVR_T-area. Further MR analysis indicated that the genetic factors associated with the use of metformin or A10 have a putative causal effect in terms of an 11% decrease in T-wave area compared to individuals not taking these medications.

This study had several limitations. Firstly, GWAS of ECG was based on the earliest ECG of the samples in UKB. Further studies are required to leverage the longitudinal ECG data of samples in order to consider ETC changes during aging. Secondly, the limited number of samples for which ECG information was available reduced the power of our GWAS analysis to detect SNPs associated with ETCs and in the MR analysis. Thirdly, all the analyses in this study were based on data from Europeans because ECG and genetic data from other ethnicities are quite limited.

## Conclusions

GWAS was performed by extracting comprehensive traditional and non-traditional ETCs and identifying 62 novel genetic loci as being significantly associated with them. The genetic relationship between ETCs, the use of diabetes medication and CVDs was investigated, and supported the putative causal effects of the use of diabetes medication on the aVR_T-area and the risk of IHD and CA. These findings suggest new avenues for the use of ECG in the monitoring or early diagnosis of IHD or CA risk in patients taking diabetes medications.

## Data Availability

All data produced in the present study are available upon reasonable request to the authors

## Acknowledgments

Data for this study were obtained from the UK-Biobank (application number #65805).

## Legends

**Graphical abstract.** Firstly, genome-wide association studies (GWAS) were performed for 168 ECG trace characteristics (ETCs) extracted from the 12-lead ECGs of 42,340 Europeans in the UK Biobank which identified 124 independent single nucleotide polymorphisms (SNPs) significantly associated with at least one ETC (62 were novel). Secondly, the phenotypic relationships were analyzed by regression model and found significant phenotypic association between 62 ETCs, 8 CVDs and A10/metformin use, and between 125 ETCs and 10 CVDs. Thirdly, pairwise genetic correlations of diabetes medications use (ATC code: A10 drugs, and metformin), T-wave area in lead aVR (aVR_T-area), and the risks of ischemic heart disease (IHD) and coronary atherosclerosis (CA) were identified by linkage disequilibrium score regression (LDSC). Then, Mendelian randomization (MR) support a putative causal effect of diabetes medications use on decreasing aVR_T-area, and on increasing risk of IHD and CA.

