## Supplementary material for "Genetic evidence for T-wave area from 12-lead electrocardiograms to monitor cardiovascular diseases in patients taking diabetes medications": supple_figure0718ya.docx

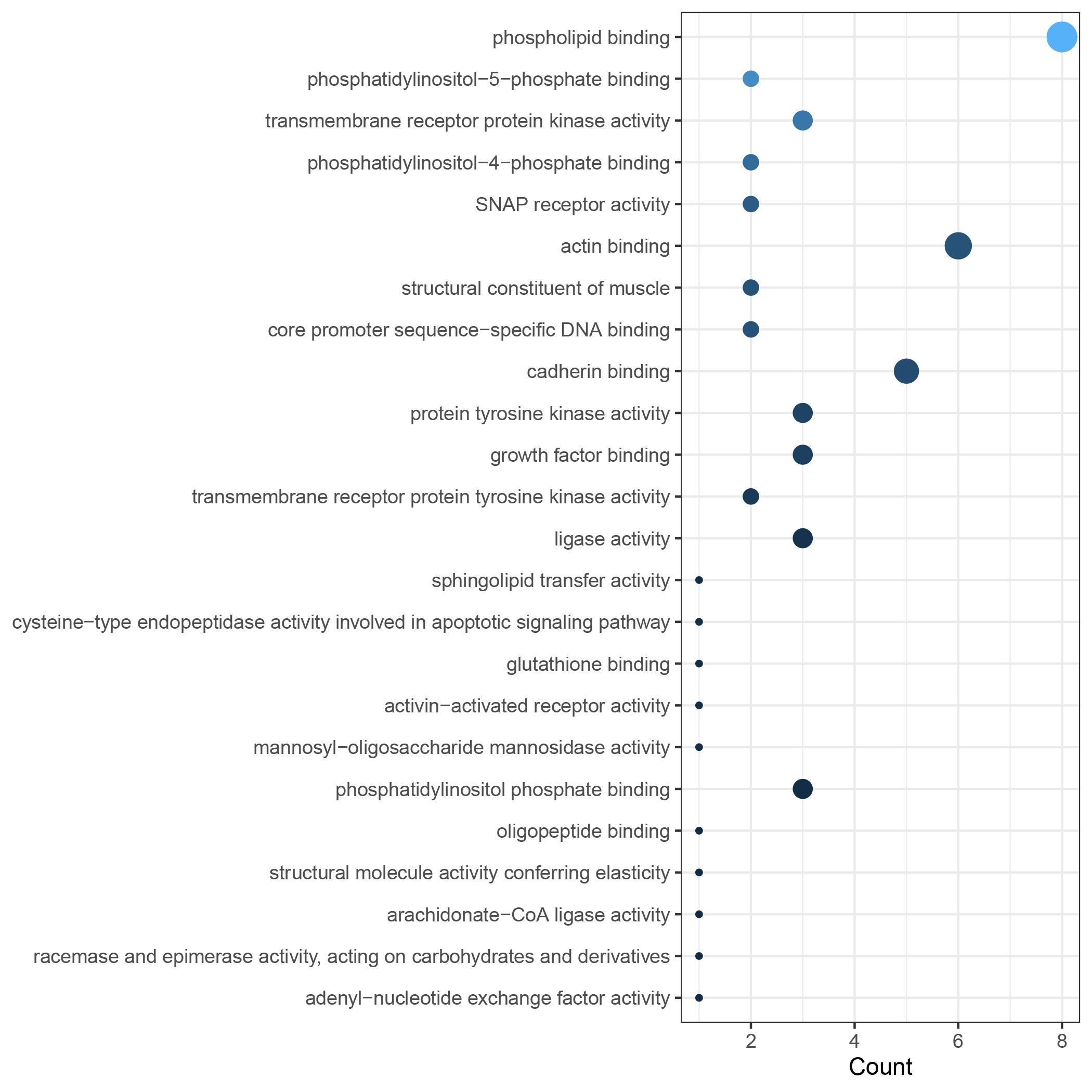

Supplementary Figure 1. The molecular function of gene ontology enrichment analysis for gene set of the significant SNPs regulatory targets. The *x* axis and size of the circles represent the enriched gene number. All circles in this Supplementary Figure shows P-value <0.05.

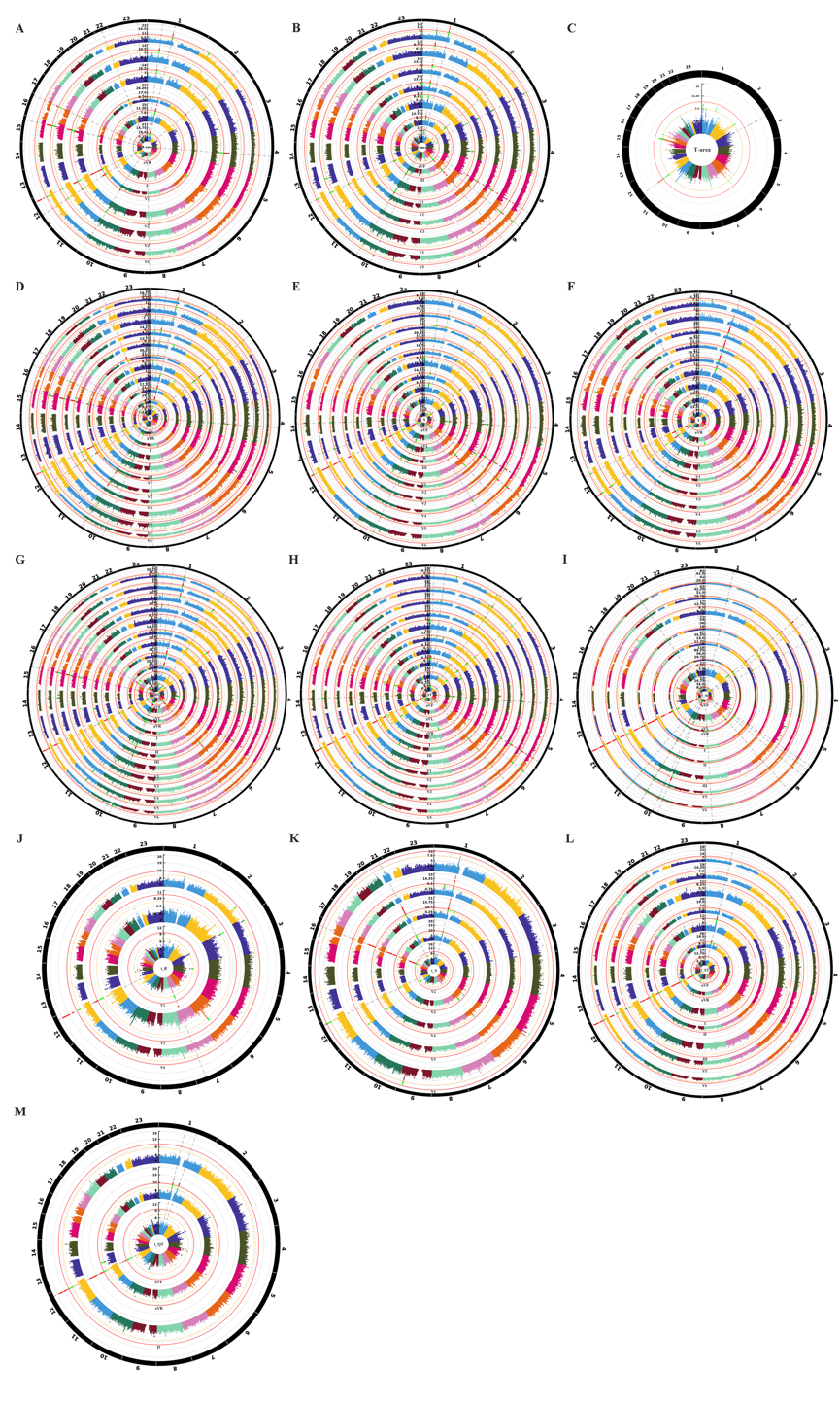

Supplementary Figure 2. Genome wide association study for the ETCs. A-M represent the ETCs of R-area, S-area, T-area, R, S, T, RS, ST, J_up, t_R, t_S, t_ST, and t_QT respectively. The name of ETC was marked in the center. The number outside the outermost circle is chromosome number. The text (corresponding to the nearest endocentric circle) between each circle is the lead name. Each circle represents a lead (except the outermost circle). The number (corresponding to the nearest endocentric circle) between each circle is the –log10(5e-8) for the most significant SNP. yellow dotted line shows the threshold P-value < 5e-8 and red solid line shows the threshold P-value < 5e-8/168. Green points represent SNPs with P-value<5e-8 but >5e-8/168, red points represent SNPs with P-value < 5e-8/168.

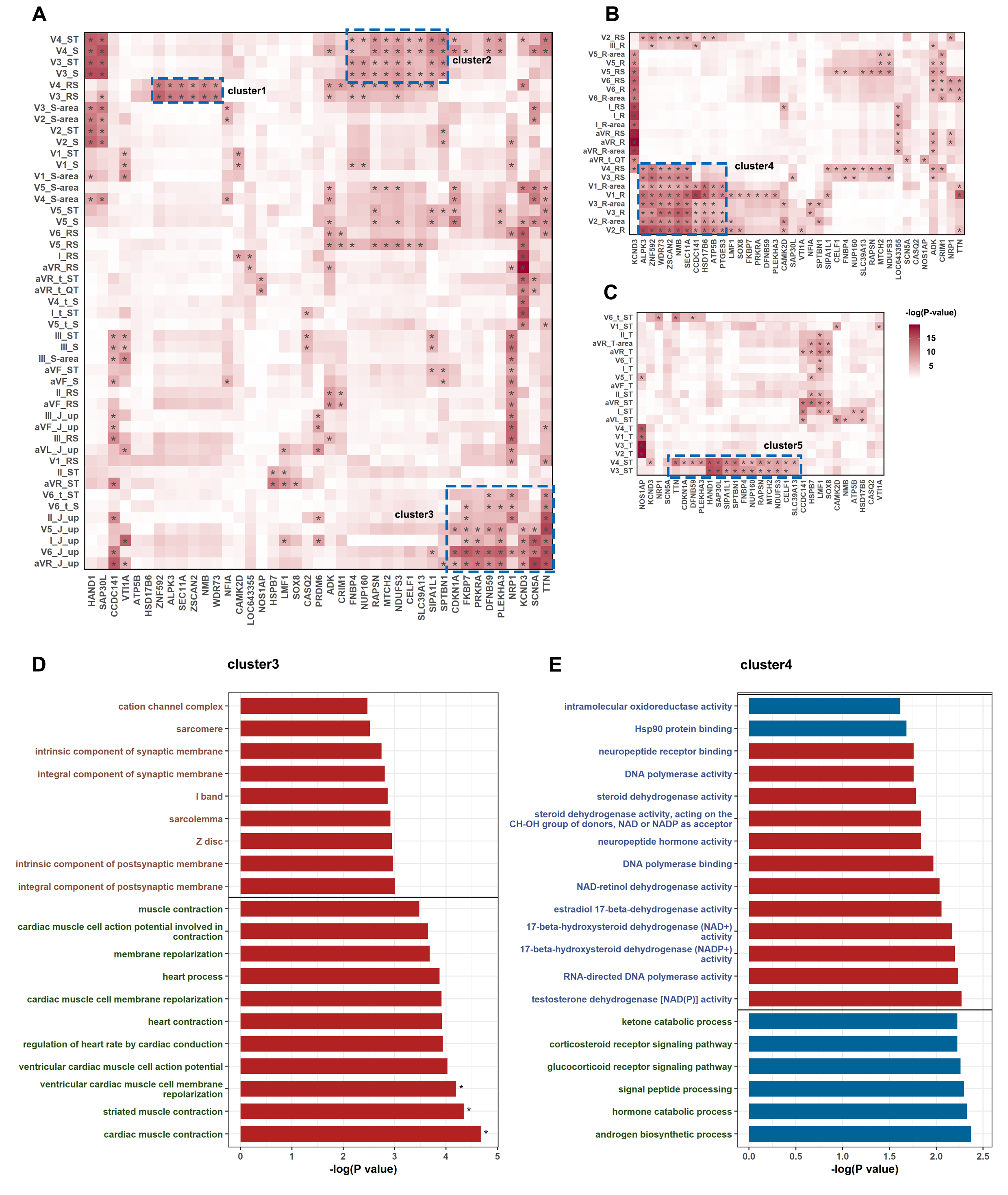

Supplementary Figure 3. The genome-wide gene-base association analysis for ETCs. The association between genes and S wave related ETCs (A), R wave related ETCs (B), T wave related ETCs (C). Darker in color means lower P-value. The symbol “*” represents the P-value < 0.05/18763. The genes and ETCs in one blue box are the ETCs significantly associated with the same genes. One blue box represents one cluster. The 20 gene ontologies with FDR <0.05 in enrichment analysis for cluster3 (D), cluster4 (E), from Figure 3A, B. The color in text of axis of (D) and (E) means different types of gene ontology. Red represents cellular component; blue represents molecular function and green represents biological process. The bar in red means FDR < 0.05 and the mark “*” on the right side of bar represents Bonferroni adjusted P-value < 0.05. Correction of P-value was in respective to the number of genes involved in the gene ontology analysis.

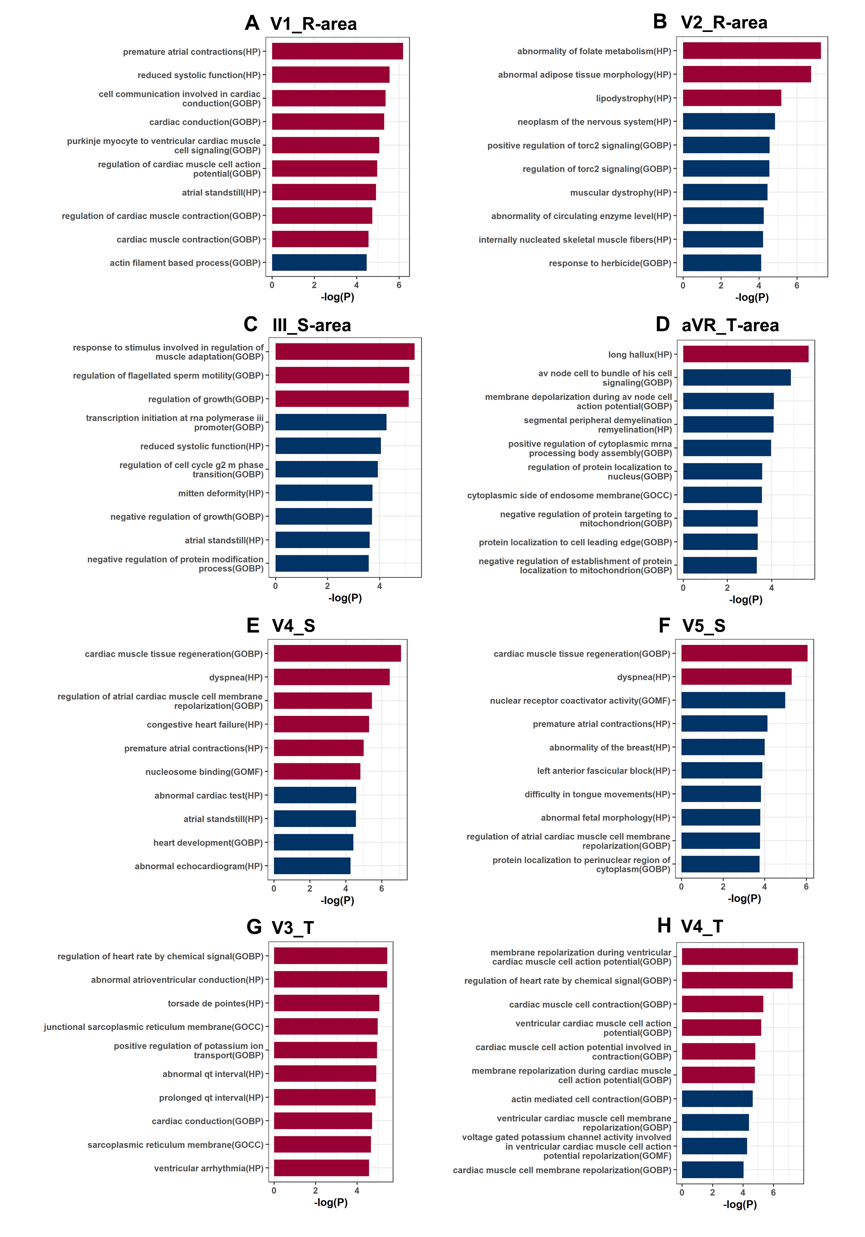

Supplementary Figure 4. Gene-set analysis for ETCs by MAGMA. The names of ETCs were displayed on each plot. *Y* axis shows the top 10 gene sets with lowest P-value and X axis is the -log10(P-value). Bar in red means FDR < 0.05.

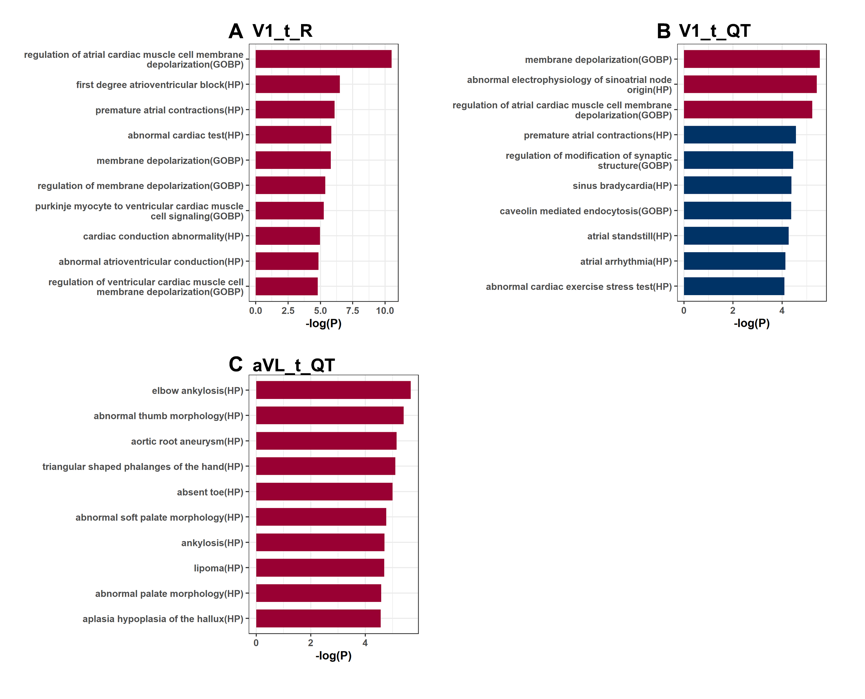

Supplementary Figure 5. Gene-set analysis for ETCs by MAGMA. The names of ETCs were displayed on each plot. *Y* axis shows the top 10 gene sets with the lowest P-value and *x* axis is the -log10(P-value). Bar in red means FDR < 0.05.

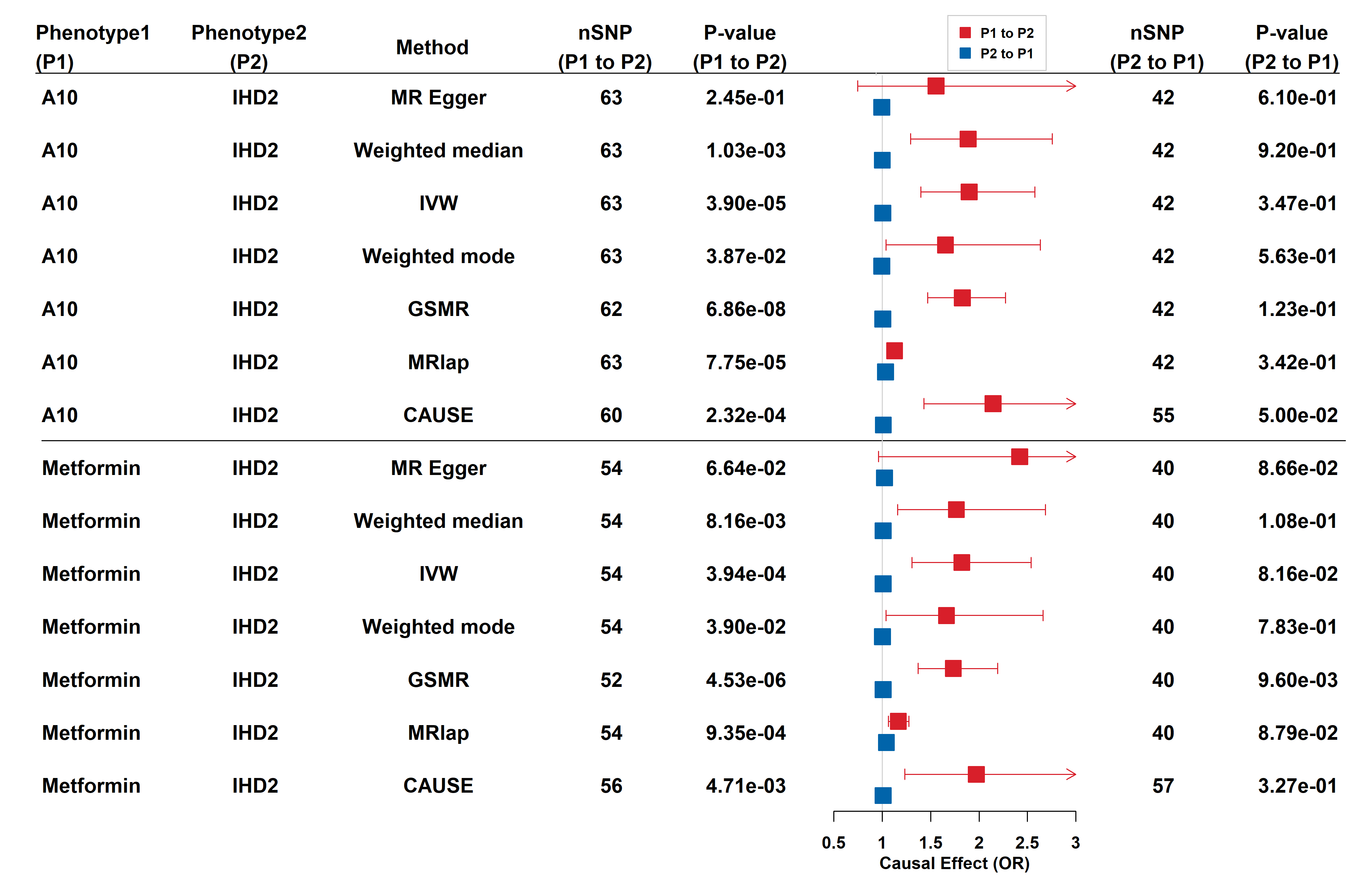

Supplementary Figure 6. MR analysis of A10 and metformin on IHD2. The causal relationship (in log-scale odds ratio [OR]) of taking A10 or metformin on IHD2 was estimated by seven MR methods. IHD2: I9_IHD, wide definition ischemic heart disease.
