## Supplementary material for "Genetic evidence for T-wave area from 12-lead electrocardiograms to monitor cardiovascular diseases in patients taking diabetes medications": supple_material0818.docx

**Method**

**Selecting European ancestry from UK-Biobank**

European ancestry was selected for using a two-stage approach: first, calculate the top two principal components (PCs) for individuals using HapMap 3 (HM3) of the 1000 Genomes Project as the strategy used in a previous study[1]; second, assign individuals as Europeans if their PC information was optimally matched to the 1000 Genomes Project European reference (compared to other population references, e.g., African, East Asian, South Asian, and Admixed)[2]. The ECG record of the first entry time was used.

**Preprocessing and feature extraction of ECG**

We first corrected the base line by subtracting the average value of the ECG signal for each lead, then dividing the 10-second ECG signal into 1-second ECG signal and removing the noise of each ECG segment. The noise removing criteria were as follows: (1) the ECG signal in one-second is a straight line (the value of the voltage is always the same number), (2) the difference between the maximum and minimum value ECG signals in one second is greater than twice that of the median difference in 10-second ECG. After removing these segments, if the data length was less than 5 seconds, the 10-second ECG data was discarded. After the quality control, each 10-second-long ECG signal was synthesized into one heartbeat for each lead by averaging the beats. Briefly, for each lead, the position of the S wave was first defined. The 230 time points before the S wave, together with the S wave and 230 time points after the S wave, were considered as a continuous heartbeat. Then, the average ECG signal of the beats was calculated. The same procedure was performed on all ECG leads. For the average heartbeat of 12-lead ECG, 168 ECG trace characteristics (ETCs) (14 for each lead) were obtained, which included 36 wave area-related characteristics. A detailed description of these ETCs is given in Supplementary Table 2.

**Gene-based genome-wide association studies**

To identify genes associated with ETC, we performed gene-based genome-wide association analysis using Multimarker Analysis of GenoMic Annotation (MAGMA) [3]. The input of MAGMA were the summary statistics of the GWAS analysis for each of 168 ETCs. A total of 18,764 genes (each harboring at least one SNP in the GWAS) from the National Center for Biotechnology Information (NCBI) in version 37.3 were used for this analysis. Bonferroni correction was applied to correct the gene association test. A gene was associated with a given ETC if the P-value given by MAGAM analysis was < 2.66×10^−6^.

**GWAS data sources**

The GWAS summary statistics for the use of metformin were downloaded from MRC-IEU[4], and involved 462,933 European samples. The GWAS summary statistics pertaining to the use of A10 were obtained from a previous study[5]. Here, A10 is an Anatomical Therapeutic Chemical Classification System (ATC) index obtained from <https://www.whocc.no/atc_ddd_index/>, representing drugs used in diabetes. From the FinnGen database (version: R6, <https://www.finngen.fi>), GWAS summary statistics of cardiovascular diseases (CVDs) were downloaded. The selected CVDs used in this study contained 23 GWAS summary statistics whose case ratios were greater than 1%. Supplementary Table 1 provides comprehensive information on each GWAS summary statistic. For each GWAS summary, SNP effects ($\beta$) and their associated standard errors (*SE*) for medication use and CVDs were converted to a quantitative scale using the approximate approach, $\beta^{'}(or {SE}^{'})=\frac{\beta(or SE)}{\mu(1-\mu)}$, where 𝜇 is the proportion of cases.

**Heritability and genetic correlation**

Single-trait and cross-trait linkage disequilibrium score regression (LDSC) were performed to estimate the liability-scale heritability (*h*^2^) of ETCs, CVDs and medication use as well as their pairwise genetic correlation (*r*_g_)[6, 7]. SNPs were excluded from LDSC analysis if they resided within the major histocompatibility complex (MHC) region (chromosome 6: 28,477,797 – 33,448,354), were strand-ambiguous (i.e., the A/T and G/C SNPs), or had an MAF less than 0.01. The default 1000 Genomes Project European-based LD score reference panels were employed throughout the analyses.

**Gene-set enrichment analysis**

The genome-wide gene-based association studies have provided many candidate genes associated with each ETC and are able to cluster the ETCs according to the genes simultaneously associated with multiple features. The ETCs were first classified into three large categories viz. R wave-related, S wave-related and T wave-related ETCs. The ETCs in each category were further grouped using “hclust” function in R4.0.2. We considered that ETCs formed one specific cluster if they were simultaneously associated with the same genes. Genes associated with the ETCs in one cluster were analyzed in terms of their enriched Gene Ontology (GO) annotation [8, 9] by clusterProfiler[10]. If the FDR was < 0.05, that GO annotation was enriched in the set of genes.

**Mendelian Randomization analysis**

MR evaluates the causal effect of a risk factor (i.e., exposure) on a target trait (i.e., outcome) using genetic variants as instruments, assuming that the genetic variants involved are significantly associated with the exposure and have causal effects on the outcome only through the exposure. This latter assumption may be violated due to the presence of horizontal pleiotropy, which occurs if the genetic variants affect the outcome through non-causal pathways[6]. Horizontal pleiotropy can itself be subclassified into *correlated pleiotropy* which is defined as the genetic variants acting on exposure and outcome via shared factors, and *uncorrelated pleiotropy* that occurs if the genetic variants act on exposure and outcome via other independent pathways.

MR analyses were carried out using R packages “*cause*” (version: 1.2.0), “*gsmr*” (version: 1.0.9), “MRlap” (version 0.0.2), and “*TwoSampleMR*” (version: 0.5.6), respectively. CAUSE selects instrumental SNPs by LD pruning, which are SNPs with *P-value* of exposure GWAS $<$ 5$\times$10^-8^ for taking medications and CVDs, and *P-value* $<$ 1$\times$10^-5^ for ETCs; other MR approaches determined independent instrumental SNPs (with GWAS *P-value* $<$ 1$\times$10^-5^ for ETCs, with GWAS *P-value* $<$ 5$\times$10^-8^ for medication use, and CVDs) by LD clumping (LD *r^2^* $<$ 0.05 within 1,000-kb windows) using PLINK (v1.90)[11]. Instrumental SNPs were further filtered out if they were located within the MHC region[7], had a MAF less than 0.01, or were nominally significantly associated with the outcome (as potential pleiotropic SNPs). In the MR analysis, pleiotropy tests were carried out by “mr_pleiotropy_test”, a function in the package of TwoSampleMR. If the pleiotropy test P-value < 0.05, an outlier test was performed to filter SNPs using the R package (“MR-PRESSO”, <https://github.com/rondolab/MR-PRESSO>). The MRlap was performed using the same instrumental variables as the other MR methods (except CAUSE) after eliminating the outlier SNPs. The complete code is available on GitHub (see web resources).

To reduce the false positive in the MR analysis, seven methods were used in this study, which include IVW, MR-Egger, Weighted mode, weighted median, GSMR, CAUSE, and MRlap. IVW estimates the Wald ratio for each SNP and calculates the causal estimate using a weighted linear regression which does not correct for horizontal pleiotropy[12]. MR-Egger adds an extra intercept to IVW to weigh the possible deviations attributable to uncorrelated pleiotropy[13]. Weighted mode greatly relaxes the assumptions made on correlated and uncorrelated pleiotropy and measures the causal effect only from the most frequent SNP set with consistent effect[14]. The weighted median calculates the causal effect using the weighted median of the SNP ratio under the assumption that most instrumental variants are valid (i.e., more than half the instrumental variants are valid instrumental SNPs)[15]. GSMR is an extension of IVW which applies the heterogeneity in dependent instruments (HEIDI)-test to exclude instrumental SNPs with potentially uncorrelated pleiotropic effects[16, 17]. Bayesian-based CAUSE corrects both correlated and uncorrelated pleiotropy via a multivariate linear model adjusted by a joint distribution of instrumental SNPs, assuming that true causality can be attributed to all instrumental SNPs whereas correlated and uncorrelated pleiotropy only affects partial instrumental SNPs[18]. CAUSE further examines the model fitness using the expected log pointwise posterior density (ELPD) by comparing the causal model (i.e., exposure affects outcome via both causal effect and pleiotropic effects), the shared model (i.e., exposure affects outcome only via pleiotropic effects), and the null model (i.e., no causal effect or pleiotropic effects between exposure and outcome). MRlap is a robust approach for correcting bias introduced by sample overlap, winner’s curse and weak instrumentation[19].

The MR estimates (i.e., $\beta_{xy}$) were converted to liability scale using one of the formulas described by Byrne et al.[20, 21]:

$\beta_{xy(liab\left( y \right):liab(x))}=\frac{Z_{K_{x}}K_{y}(1-K_{y})}{Z_{K_{y}}K_{x}(1-K_{x})}\beta_{xy(logit\left( y \right):logit\left( x \right))}$	(1)

and

$\beta_{xy(liab\left( y \right):liab(x))}=\frac{K_{y}(1-K_{y})}{Z_{K_{y}}}\beta_{xy(logit\left( y \right):liab\left( x \right))}$	(2)

where *x* and *y* represent the exposure and the outcome, *Kx* and *Ky* are the population prevalence of the exposure and the outcome, and *Z_Kx_* and *Z_Ky_* are the values of the standard normal distributions of the exposure and the outcome at such prevalence. *Equation (1)* was used when *y* and *x* are binary traits (e.g., medication-use) and when *y* is a binary trait (e.g., CVD) and *x* is a quantitative trait (e.g., an ETC), *Equation (2)* was used to convert the MR estimates to the liability scale. The population prevalence for CVDs can be found in Supplementary Table 3. Here, the "prevalence" of medication use was the ratio of medication use in GWAS samples.

**Results**

**Genome-wide significant loci associated with areas of R-wave, S-wave and T-wave**

This is the first study to perform GWAS analysis for wave area-related ETCs (ETC) including R-wave area (R-area), S-wave area (S-area) and T-wave area (T-area). Here, the SNPs associated with areas of R wave, S wave and T wave were further analyzed.

In total, 15 independent SNPs were found to be significantly associated (P-value<$5\times{10}^{-8}$/168) with the R-area (Figure 2A-B, Table1). Of these, 2 (rs72694622 and rs75013985), 1 (rs72694603), 5 (rs35596070, rs72840788, rs12227117, rs7132327 and rs133890), 6 (rs4915740, rs35430511, rs55679363, rs80191567, rs7132327, and rs4633690), 3 (rs4915740, rs4633690 and rs8046873), and 2 (rs72694622 and rs7977151) independent SNPs were found to be associated with R-area in lead aVR, I, V1, V2, V3, and V6, respectively (Figure 2A-B, Table1). Among those, rs4915740 is significantly associated with R-area in both leads V2 (P-value = 4.6$\times{10}^{-11}$) and V3 (P-value = 6.7$\times{10}^{-12}$), and is also significantly (P-value = 3.1$\times{10}^{-5}$) associated with the left bundle branch block. The SNPs, rs72694603 (lead I: P-value= 6.5$\times{10}^{-12}$), rs72694622 (lead V6: P-value= 1.2$\times{10}^{-13}$ ; lead aVR: P-value=2.1$\times{10}^{-14}$ ) and rs75013985 (lead aVR: P-value= 1.4$\times{10}^{-15}$ ) are all located in the *KCND3* gene that is known to activate voltage-gated potassium channels [22], and contributes to A-type (transient outward) potassium channel activity affecting ventricular contraction [23]. Another SNP, rs35430511, located in the *CAMK2D* gene, was also found to be significantly associated (P-value= 1.1$\times{10}^{-14}$) with R-area in lead V2. The *CAMK2D* gene is involved in the regulation of Ca^2+^ homeostatis and excitation-contraction coupling in the heart[24, 25].

In all, 23 independent SNPs were found to be significantly associated with S-area (Figure 2A-B, Table1). Five SNPs (rs5016273, rs2207791, rs4915741, rs2207790, and rs11207739) located in the *NF1A* gene were found to be associated with S-area in leads aVF, III and V2~V4. *NFIA* has been previously reported to be associated with QRS duration[26-28], electrocardiographic conduction measures[29, 30], left bundle branch block and congestive heart failure[31]. Four SNPs (rs13165478, rs62379942, rs10054375 and rs10076436) were found to be associated with S-area in lead V1~V4. These SNPs are also eQTLs of *SAP30L* and *GALNT10,* genes which are known to fulfil functions related to ECG trace characteristics[32], autism spectrum disorder[33] and schizophrenia[34]. Another SNP, rs59365541, was found to be associated with S-area in lead aVF (P-value = 8.2$\times{10}^{-12}$) and III (P-value=2.8$\times{10}^{-17}$) in this study. Two SNPs (rs12776791 and rs10885379) in the *VTI1A* gene were significantly associated with S-area in lead V1 (P-value = 3.1$\times{10}^{-11}$) and III (P-value= 9.6$\times{10}^{-12}$) respectively.

Two SNPs, rs7638275 (P-value= 3.5$\times{10}^{-15}$) and rs4963759 (P-value= 4.7$\times{10}^{-12}$), were found to be significantly associated with T-wave area (T-area) in lead aVR (Figure 2A-B, Table1). rs7638275 is located in the *SCN5A* gene that encodes a protein with functions related to voltage-gated sodium channel activity[35]. rs4963759 resides in the *SOX5* gene which is known to harbor many SNPs associated with electrocardiographic features, PR interval[36] and resting heart rate[37].

**Genome-wide significant loci associated with amplitude traits**

As shown in Manhattan plots (Figure 2B), 35 independent SNPs are significantly associated with the amplitude difference between the end of the S wave and the start of the R wave (J_up). Among them, two independent SNPs, rs7132327 and rs3914956 which both localize to 12q24.21, were significantly associated with the J_up in lead aVF, aVL, aVR, I, II, III, V5 and V6, and S wave amplitude and T wave amplitude (Figure 2A and Supplementary Figure 2E-F). In addition, four SNPs (rs2562834, rs546095518, rs11902709 and rs6715901) in the *TTN* gene were found to be associated with J_up in lead V5, V6, aVR, and II, and another SNP, rs16866352, the eQTL of *TTN,* is associated (P-value= 2.3$\times{10}^{-10}$) with J_up in lead aVL. *TTN* gene is known to contribute to the fine balance of forces between the two halves of the sarcomere whilst *TTN* mutations have been reported to be associated with cardiomyopathy[38, 39]. An additional five SNPs were found to be associated with J_up in lead III, II, and aVF are in the *NRP1* gene (rs78656993 and rs1888684) or the eQTLs of *NRP1* (rs1888684, rs10763928, rs10827243 and rs10827246). The *NRP1* gene is known to be involved in the development of the cardiovascular system and angiogenesis[40, 41].

We have found 31 SNPs to be associated with R amplitude (R) (Figure 2A and Supplementary Figure 2D), which include the SNPs (rs12090194, rs72694603, rs72694622 and rs75013985) located in the *KCND3* gene associated with R in lead aVR, I, V5 and V6, and SNPs (rs13031826 and rs35596070) located in the *CCDC141* gene associated with R in lead III, V1 and V2. Another SNP, rs4633690, was found to be significantly associated with R amplitude in lead V1, V2, V3 and V4, and it is the eQTL of *SCAND2P*, *NMB*, *AC103965.1*, *SEC11A*, *ZSCAN2*, *ALPK3*, *WDR73* and *UBE2Q2P1*. *NMB* is associated with hormone activity[42] and neuropeptide hormone activity[22], both of which may affect cardiac function. *ALPK3* is involved in cardiomyocyte differentiation and is known to be associated with cardiomyopathy[43, 44]. A total of 20 independent SNPs (Supplementary Figure 2H) were found to be significantly associated with S amplitude, of which six are located in 12q24.21 and are significantly associated with S amplitude in lead aVF, II, III and V3~V6. Among them, rs4151702, located in the *DINOL* gene, is significantly associated with both S amplitude in lead V4 and V5. In total, 10 SNPs have been found to be associated with T amplitude (Supplementary Figure 2F). A SNP, rs2074238 located in the *KCNQ1* gene that is known to be associated with KCNE beta subunits that modulate current kinetics and induces a voltage-dependent current by rapidly activating and slowly deactivating potassium-selective outward current[45, 46].

**Genome-wide significant loci associated with time-interval traits**

In total, 11 SNPs were identified as being significantly associated with the time intervals of the R-wave and S-wave. No SNPs were identified as being associated with the time interval of the T-wave (Figure 2A). Briefly, three SNPs (rs1997571, rs1896356 and rs7132327) (Supplementary Figure 2A, Supplementary Table 4) were identified as being significantly associated with the time interval of the R-wave (t_R), and eight SNPs (rs72692602, rs72694622, rs75013985, rs6416327, rs4784934, rs28623612, rs28627526 and rs2223036) as being significantly associated with the time interval of S-wave (t_S) in chest lead (V2~V6). Of these, rs72692602 located in the *KCND3* gene, was found to be significantly associated with t_S in lead V3, V4, and V5 (Supplementary Figure 2B). The SNPs, rs4784934, rs28623612, and rs28627526 were found to be significantly associated with t_S in lead V2~V3, V4, and V5, respectively. These SNPs represent eQTLs for the *RP11-481J2.2* and *GINS3* genes. Additionally, nine SNPs (rs11102347, rs2120436, rs72692597, rs7309985, rs1910046, rs9630280, rs7132327, rs7301677 and rs10850409) were found to be associated with the S begin-to-T end interval (t_ST) (Supplementary Figure 2C), of which three are located in the *KCDN3* gene (lead aVR and I related SNPs: rs11102347, rs2120436, rs72692597) and the remaining six SNPs are located in 12q24.21. Finally, six SNPs were found to be associated with the QT interval (t_QT) (Supplementary Figure 2I), including three SNPs (rs4767282, rs7959283, and rs10850409) in 12q24.21 associated with t_QT in lead aVF, aVR and II.

Overall, we identified 124 independent SNPs that were significantly (P-value <$\frac{5\times{10}^{-8}}{168}$) associated with ETCs, of which 62 (Table 1) were reported to be associated with other ETCs or CVDs, and 62 were newly identified as being associated with ETCs in this study. Specifically, this study found SNPs in the *NFIA*, *KCND3* and *CCDC141* genes, and in the 12q24.21 region to be associated with more than 10 ETCs, suggesting the important roles of these genomic region in characterizing ECG. We also performed gene-level genome-wide association analysis as shown in Supplementary Material.

**Genes and gene sets associated with ETCs**

We performed genome-wide gene-based association analysis using Multimarker Analysis of GenoMic Annotation (MAGMA) [3]. In total, 148 genes were identified as being significantly (Bonferroni adjusted *P* < 0.05) associated with at least one ETC (Supplementary Table 5). As shown in Supplementary Figure 3A-C, many genes form clusters that are consistently associated with a group of ETCs. For example, the *KCND3* gene was significantly associated (P-value < 0.05/18176) with 29 ETCs, I_J_up, aVR_J_up, V5_J_up, V6_J_up, V4_ST, V4_RS, V5_S-area, V5_S, V5_RS, V5_R, V5_R-area, V6_R-area, V6_R, V6_RS, aVR_R-area, aVR_R, aVR_RS, aVR_t_QT, aVR_t_ST, aVL_R, I_t_ST, I_R-area, I_R, I_RS, V3_t_S, V4_t_S, V5_t_S, V6_t_R, and V6_S) (Supplementary Table 5).

The S-wave related traits formed several clusters. Two genes, *HAND1* and *SAP30L,* were found to be significantly associated with 10 S-wave related features (Supplementary Figure 3A), namely V2_S, V2_ST, V2_S-area, V3_S-area, V3_RS, V4_RS, V3_S, V3_ST, V4_S, and V4_ST). Six genes (*ZNF592, ALPK3, SEC11A, ZSCAN2, NMB, WDR73*) were found to be associated with RS in lead V3 and V4 (cluster 1 in Supplementary Figure 3A). An additional group of genes (*FNBP4*, *NUP160*, *RAPSN*, *MTCH2*, *NDUFS3*, *CELF1*, *SLC39A13*, *SIPA1L1* and *SPTBN1*) was found to be significantly associated with S-wave related ETCs including V3_S, V3_ST, V4_S and V4_ST (cluster 2 in Supplementary Figure 3A) which are enriched in postsynaptic specialization (GO:0099572, P-value=4.06$\times{10}^{-4}$, FDR=2.31$\times{10}^{-2}$). Cluster 3 (Supplementary Figure 3A) includes 9 genes (*CDKN1A*, *FKBP7*, *PRKRA*, *DFNB59*, *PLEKHA3*, *NRP1*, *KCND3*, *SCN5A* and *TTN*) that are significantly associated with the traits (aVR_J_up, V6_J_up, I_J_up, V5_J_up, II_J_up, V6_t_S, and V6_t_ST). These genes are significantly (FDR < 0.05) enriched in biological processes (BP) such as cardiac muscle contraction, cardiac conduction and action potential. They are also enriched in many different cellular components (CC) and molecular functions (MF) as shown in Supplementary Figure 3D and Supplementary Table 6. The R-wave related features formed one cluster (cluster 4) according to their associated genes (Supplementary Figure 3B), which include R amplitude, R-area in lead V1~V3 and RS in lead V3, V4 (cluster 4 in Supplementary Figure 3B) and are associated with 10 genes (*ALPK3*, *ZNF592*, *WDR73*, *ZSCAN2*, *NMB*, *SEC11A*, *CCDC141*, *HSD17B6*, *ATP5B* and *PTGES3*). These genes are enriched (FDR <0.05) in 12 molecular functions, such as 17-beta-hydroxysteroid dehydrogenase (NADP+) activity and NAD-retinol dehydrogenase activity (Supplementary Figure 3E). In cluster 5 (Supplementary Figure 3C), 15 genes, namely *TTN*, *CDKN1A*, *DFNB59*, *PLEKHA3*, *HAND1*, *SAP30L*, *SIPA1L1*, *SPTBN1*, *FNBP4*, *NUP160*, *RAPSN*, *MTCH2*, *NDUFS3*, *CELF1* and *SLC39A13,* are significantly associated with ST in lead V3 and V4. In summary, the genes that are commonly associated with a group of ETCs represent candidates for involvement in the shared genetic mechanisms that underlie these features. Additional gene function analysis was performed on genes associated with individual ETCs as shown in the Supplementary Material.

**Gene-set enrichment analysis**

For genes associated with individual ETCs, we performed gene-set enrichment analysis on GO annotation and Human Phenotype Ontology (HPO) [47] obtained from the MsigDB version v2022.1 by using MAGMA. A set of genes were considered as being enriched with certain gene functions if the FDR provided by MAGMA was less than 0.05.

In total, 42, 42, 38 and 37 genes were found to be significantly associated (FDR<0.05) with R-area in lead V1~V4, respectively. The genes associated with V1_R-area were significantly enriched in six biological processes (regulation of cardiac muscle contraction [FDR=3.58$\times{10}^{-2}$], cardiac muscle contraction [FDR=4.83$\times{10}^{-2}$], cardiac conduction [FDR=1.97$\times{10}^{-2}$], Purkinje myocytes to ventricular cardiac muscle cell signaling [FDR=2.71$\times{10}^{-2}$], cell communication involved in cardiac conduction [FDR=1.97$\times{10}^{-2}$], regulation of cardiac muscle cell action potential [FDR=2.78$\times{10}^{-2}$]) according to Gene Ontology analysis, and three phenotypes (reduced systolic function [FDR=1.97$\times{10}^{-2}$], premature atrial contractions [FDR=1.02$\times{10}^{-2}$], atrial standstill [FDR=2.78$\times{10}^{-2}$]) from Human Phenotype Ontology (HPO) (Supplementary Figure 4A). The genes that were significantly associated with the V2_R-area are significantly enriched in three phenotypes (abnormal adipose tissue morphology [FDR=1.41$\times{10}^{-3}$], lipodystrophy [FDR=3.51$\times{10}^{-2}$], and abnormality of folate metabolism [FDR=8.57$\times{10}^{-4}$]) from HPO (Supplementary Figure 4B).

For S-wave related amplitude ETCs, aVL_S, V1_S, V3_S, V4_S and V5_S, a total of 18, 14, 36, 50 and 22 genes were found significantly associated with them, respectively. Among them, the genes associated with V4_S were significantly enriched in two biological processes (regulation of atrial cardiac muscle cell membrane repolarization [FDR=1.84$\times{10}^{-2}$], cardiac muscle tissue regeneration [FDR=1.34$\times{10}^{-3}$]), one molecular function (nucleosome binding [FDR=3.99$\times{10}^{-2}$]), and three human phenotypes (congestive heart failure [FDR=2.00$\times{10}^{-2}$], dyspnea [FDR=2.78$\times{10}^{-3}$], premature atrial contractions [FDR=3.17$\times{10}^{-2}$]) in HPO (Supplementary Figure 4E). The genes associated with V5_S were enriched in cardiac muscle tissue regeneration [FDR=1.40$\times{10}^{-2}$] and dyspnea [FDR=3.96$\times{10}^{-2}$] whilst the genes associated with V1_S were enriched in cartilage morphogenesis [FDR=6.50$\times{10}^{-3}$] from GO (Supplementary Figure 4F) and glyoxylate and dicarboxylate metabolism [FDR=4.86$\times{10}^{-2}$] from KEGG. There were 3 genes significantly associated with III_S-area, and the SNPs associated with III_S-area were significantly enriched in three biological processes (response to stimulus involved in regulation of muscle adaptation [FDR=4.01$\times{10}^{-2}$], regulation of growth [FDR=4.01$\times{10}^{-2}$], and regulation of flagellated sperm motility [FDR=4.01$\times{10}^{-2}$]) (Supplementary Figure 4C).

There were 49 genes associated with T-wave amplitude, of which 14, 11, 12 and 13 were associated with aVR_T, I_T, V5_T and V6_T respectively but no significant gene-set was enriched. The genes associated with V3_T were enriched in four biological processes (regulation of heart rate [FDR=4.13$\times{10}^{-2}$], regulation of heart rate by chemical signal [FDR=2.85$\times{10}^{-2}$], positive regulation of potassium ion transport [FDR=2.89$\times{10}^{-2}$], cardiac conduction [FDR=3.69$\times{10}^{-2}$]), two cellular components (junctional sarcoplasmic reticulum membrane [FDR=2.89$\times{10}^{-2}$], sarcoplasmic reticulum membrane [FDR=3.80$\times{10}^{-2}$]) and five human phenotypes (prolonged QT interval [FDR=2.89$\times{10}^{-2}$], torsade de pointes [FDR=2.89$\times{10}^{-2}$], ventricular arrhythmia [FDR=4.13$\times{10}^{-2}$], abnormal atrioventricular conduction [FDR=2.85$\times{10}^{-2}$], abnormal QT interval [FDR=2.89$\times{10}^{-2}$]) in HPO (Supplementary Figure 4G). The genes associated with V4_T are enriched in biological processes as regulation of heart rate by chemical signal [FDR=4.20$\times{10}^{-4}$], cardiac muscle cell action potential involved in contraction [FDR=4.38$\times{10}^{-2}$], cardiac muscle cell contraction [FDR=2.43$\times{10}^{-2}$], ventricular cardiac muscle cell action potential [FDR=2.50$\times{10}^{-2}$], membrane repolarization during cardiac muscle cell action potential [FDR=4.38$\times{10}^{-2}$], and membrane repolarization during ventricular cardiac muscle cell action potential [FDR=3.81$\times{10}^{-4}$] (Supplementary Figure 4H). In total, 29 genes were found to be significantly associated with the ETC, aVR_T-area, but only one gene-set was enriched (long hallux [FDR=3.34$\times{10}^{-2}$], Supplementary Figure 4D).

Fewer genes were identified as being significantly associated with ETCs related to time interval. Even so, ten genes were found to be significantly associated with V4_t_S, and these were significantly enriched in calcium signaling pathway, hypertrophic cardiomyopathy [FDR=1.73$\times{10}^{-2}$], dilated cardiomyopathy [FDR=2.06$\times{10}^{-2}$], and aldosterone regulated sodium reabsorption [FDR=1.73$\times{10}^{-2}$]. Moreover, II_t_ST were enriched in antigen processing and presentation [FDR=2.64$\times{10}^{-2}$], type I diabetes mellitus [FDR=2.64$\times{10}^{-2}$], graft versus host disease [FDR=2.64$\times{10}^{-2}$], dilated cardiomyopathy [FDR=4.96$\times{10}^{-2}$]. Furthermore, SNPs associated with aVL_t_QT were enriched in nine (Supplementary Figure 5C) Gene Ontology (GO)[8, 9] functions, including aortic root aneurysm and elbow ankylosis.

31.	*UKBiobank TOPMed-imputed PheWeb*. Available from: <https://pheweb.org/UKB-TOPMed/>.
