## Supplementary material for "Genetic evidence for T-wave area from 12-lead electrocardiograms to monitor cardiovascular diseases in patients taking diabetes medications": supple_tables0606.docx

Supplementary Table 1. The GWAS resources of the GWAS summaries used in this study.

| **Category** | **GWAS id** | **Abbreviation** | | **Phenotype** | **Ncase** | **Ncontrol** | **Sample size** | **Resource** |
| --- | --- | --- | --- | --- | --- | --- | --- | --- |
| medication use | A10 | A10 | drugs used in diabetes | | 15272 | 290641 | 305913 | [1] |
| medication use | ukb-b-14609 | metformin | Treatment/medication code: metformin | | 11,552 | 451,381 | 462,933 | MRC-IEU[2] |
| CVDs | I9_AF | AF | Atrial fibrillation and flutter | | 28670 | 135821 | 164491 | FinnGen[3] |
| CVDs | I9_AF_REIMB | AFR | Atrial fibrillation and flutter with reimbursement | | 15256 | 135821 | 151077 | FinnGen[3] |
| CVDs | I9_AORTANEUR | AA | Aortic aneurysm | | 3658 | 244907 | 248565 | FinnGen[3] |
| CVDs | I9_ATHSCLE | ATH | Atherosclerosis, excluding cerebral, coronary and PAD | | 8391 | 244907 | 253298 | FinnGen[3] |
| CVDs | I9_CARDMPRI | CM1 | Cardiomyopathies, Primary/intrinsic | | 2685 | 182971 | 185656 | FinnGen[3] |
| CVDs | I9_CARDMYO | CM2 | Cardiomyopathy | | 3772 | 182971 | 186743 | FinnGen[3] |
| CVDs | I9_CHD | CHD1 | Major coronary heart disease event | | 25707 | 234698 | 260405 | FinnGen[3] |
| CVDs | I9_CHD_NOREV | CHD2 | Major coronary heart disease event excluding revascularizations | | 21441 | 234495 | 255936 | FinnGen[3] |
| CVDs | I9_CORATHER | CA | Coronary atherosclerosis | | 28598 | 222551 | 251149 | FinnGen[3] |
| CVDs | I9_CVD | CVD | Cardiovascular diseases | | 135546 | 124859 | 260405 | FinnGen[3] |
| CVDs | I9_HEARTFAIL | HF1 | Heart failure, strict | | 15838 | 229946 | 245784 | FinnGen[3] |
| CVDs | I9_HEARTFAIL_ALLCAUSE | HF2 | All-cause Heart Failure | | 30098 | 229612 | 259710 | FinnGen[3] |
| CVDs | I9_HEARTFAIL_NS | HF3 | Heart failure, not strict | | 30459 | 229946 | 260405 | FinnGen[3] |
| CVDs | I9_IHD | IHD2 | Ischaemic heart disease, wide definition | | 39358 | 221047 | 260405 | FinnGen[3] |
| CVDs | I9_ISCHHEART | IHD1 | Ischemic heart diseases | | 37854 | 222551 | 260405 | FinnGen[3] |
| CVDs | I9_MI | MI1 | Myocardial infarction | | 15787 | 222551 | 238338 | FinnGen[3] |
| CVDs | I9_MI_STRICT | MI2 | Myocardial infarction, strict | | 14305 | 222551 | 236856 | FinnGen[3] |
| CVDs | I9_NONRHEVALV | NRVD | Non-rheumatic valve diseases | | 12375 | 182971 | 195346 | FinnGen[3] |
| CVDs | I9_PAD | PAD | Peripheral artery disease | | 9021 | 244907 | 253928 | FinnGen[3] |
| CVDs | I9_VHD | VHD | Valvular heart disease including rheumatic fever | | 47003 | 182971 | 229974 | FinnGen[3] |
| CVDs | I9_VTE | VTE | Venous thromboembolism | | 11288 | 249117 | 260405 | FinnGen[3] |
| CVDs | I9_ANGINA | ANGINA | Angina pectoris | | 21944 | 222551 | 244495 | FinnGen[3] |

Supplementary Table 2. The description and sketch for the ETCs used in this study.

| ETC | Description | Sketch |
| --- | --- | --- |
| R | R amplitude of combined wave | 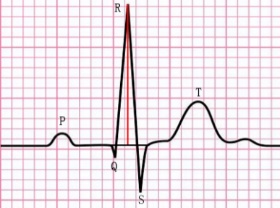 |
| S | S amplitude of combined wave | 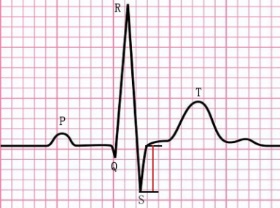 |
| T | T amplitude of combined wave | 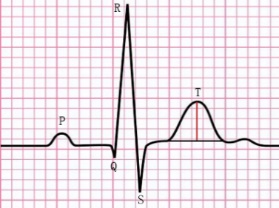 |
| RS | the difference between R and S amplitude | 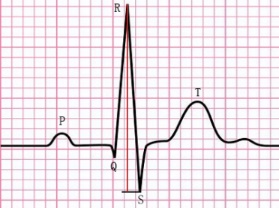 |
| ST | the difference between S and T amplitude | 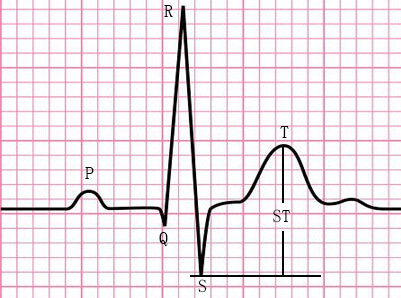 |
| J_up | difference between R start and S end in voltage | 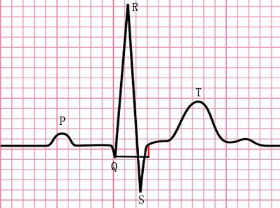 |
| R-area | R peak area | 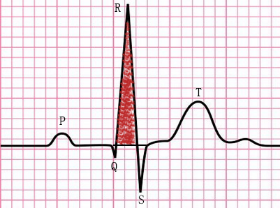 |
| S-area | S peak area | 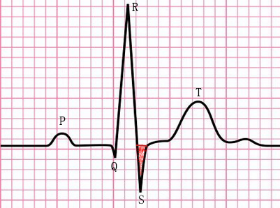 |
| T-area | T peak area | 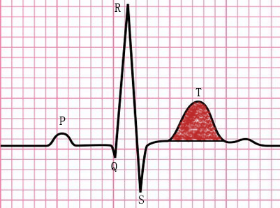 |
| t_T | T begin-to-T end interval | 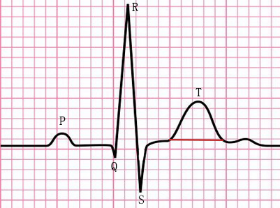 |
| t_S | S begin-to-S end interval | 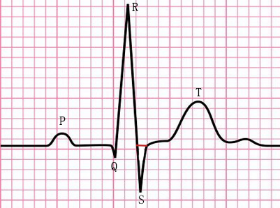 |
| t_R | R begin-to-R end interval | 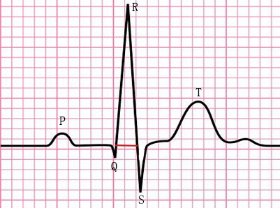 |
| t_ST | S begin-to-T end interval | 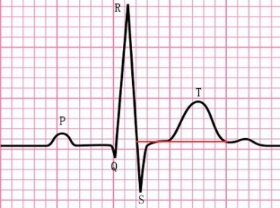 |
| t_QT | R begin-to-T end interval | 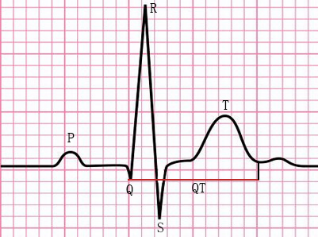 |

Supplementary Table 3. The prevalence of used cardiovascular diseases and the data resource. IHME: the prevalence came from the global burden of disease study of Institute for Health Metrics and Evaluation[4].

| **GWAS id** | **Phenotype** | **Prevalence** | **Reference** |
| --- | --- | --- | --- |
| I9_AF | Atrial fibrillation and flutter | 0.018077 | IHME |
| I9_AF_REIMB | Atrial fibrillation and flutter with reimbursement | 0.018077 | IHME |
| I9_AORTANEUR | Aortic aneurysm | 0.022 | Svensjo et al.[5] |
| I9_ATHSCLE | Atherosclerosis, excluding cerebral, coronary and PAD | 0.019 | Bergström et al.[6] |
| I9_CARDMPRI | Cardiomyopathies, Primary/intrinsic | 0.001464 | IHME |
| I9_CARDMYO | Cardiomyopathy | 0.001464 | IHME |
| I9_CHD | Major coronary heart disease event | 0.0725 | Zhu et al.[7] |
| I9_CHD_NOREV | Major coronary heart disease event excluding revascularizations | 0.0725 | Zhu et al.[7] |
| I9_CORATHER | Coronary atherosclerosis | 0.019 | Bergström et al.[6] |
| I9_CVD | Cardiovascular diseases | 0.117318 | IHME |
| I9_HEARTFAIL | Heart failure,strict | 0.010581 | Bragazzi et al.[8] |
| I9_HEARTFAIL_ALLCAUSE | All-cause Heart Failure | 0.010581 | Bragazzi et al.[8] |
| I9_HEARTFAIL_NS | Heart failure, not strict | 0.010581 | Bragazzi et al.[8] |
| I9_IHD | Ischaemic heart disease, wide definition | 0.046995 | IHME |
| I9_ISCHHEART | Ischemic heart diseases | 0.046995 | IHME |
| I9_MI | Myocardial infarction | 0.016004 | Bhatnagar et al. [9] |
| I9_MI_STRICT | Myocardial infarction, strict | 0.016004 | Bhatnagar et al. [9] |
| I9_NONRHEVALV | Non-rheumatic valve diseases | 0.013608 | IHME |
| I9_PAD | Peripheral artery disease | 0.033513 | IHME |
| I9_VHD | Valvular heart disease including rheumatic fever | 0.025 | Iung et al.[10] |
| I9_VTE | Venous thromboembolism | 0.0026 | Ghanima et al.[11] |
| I9_ANGINA | Angina pectoris | 0.025242 | Bhatnagar et al. [9] |

Supplementary Table 4. Overview of the 124 independent SNPs discovered associated with ETCs by GWAS. The independent (LD r2<0.05) genetic variant with the p-value < 5e-8/168 is shown. Annotations: GWAS: GWAS catalogue; PheWeb: UKBiobank TOPMed-imputed PheWeb: https://pheweb.org/UKB-TOPMed/; eQTLs: eQTLGen Consortium, https://eqtlgen.org/cis-eqtls.html. Chromosomes (CHR), base pair positions (POS, GRCh37) and nearest protein coding gene (Gene).

| ETC | Lead | SNP | CHR | POS | GWAS | PheWeb^a^ | eQTL | Gene |
| --- | --- | --- | --- | --- | --- | --- | --- | --- |
| R-area | V2; V3 | rs4915740 | 1 | 61886744 |  | Left bundle branch block |  | NFIA |
|  | I | rs72694603 | 1 | 112458893 |  | Atrial fibrillation and flutter; Cardiac dysrhythmias; Secondary malignant neoplasm of skin |  | KCND3 |
|  | V6; aVR | rs72694622 | 1 | 112481667 | DNA methylation variation |  |  | KCND3 |
|  | aVR | rs75013985 | 1 | 112530430 | PR interval | Paroxysmal tachycardia, unspecified; Paroxysmal supraventricular tachycardia |  | KCND3 |
|  | V1 | rs35596070 | 2 | 179759692 | Heart rate response to recovery post exercise | Atrioventricular [AV] block | SESTD1 | CCDC141 |
|  | V2 | rs35430511 | 4 | 114387138 |  |  |  | CAMK2D |
|  | V2 | rs55679363 | 8 | 125867834 | Supranormal left ventricular ejection fraction; Hematocrit; Hemoglobin concentration; PR interval |  | MTSS1 |  |
|  | V1 | rs72840788 | 10 | 121415685 | Parkinson's disease or first-degree relation to individual with Parkinson's disease; High-sensitivity cardiac troponin T levels; Left ventricular global radial strain; Left ventricular ejection fraction; Left ventricular end-diastolic volume; Left ventricular end-systolic volume; Left ventricular global circumferential strain; Free cholesterol to total lipids ratio in very large HDL; Hypertrophic cardiomyopathy; Left ventricular end-systolic volume; Left ventricular ejection fraction | Heart failure NOS; stress incontinence, female; Congestive heart failure; non-hypertensive | MCMBP; BAG3 | BAG3 |
|  | V2 | rs80191567 | 12 | 57156053 |  |  |  |  |
|  | V1 | rs12227117 | 12 | 57166413 |  | Atrial fibrillation and flutter; Cardiac dysrhythmias | RBMS2; PRIM1; SPRYD4; BAZ2A; NACA; PTGES3 | HSD17B6 |
|  | V6 | rs7977151 | 12 | 115362972 | Hypertrophic cardiomyopathy |  |  |  |
|  | V1; V2 | rs7132327 | 12 | 115381071 | QRS complex (12-leadsum); Myocardial fractal dimension; Electrocardiogram morphology |  |  |  |
|  | V2; V3 | rs4633690 | 15 | 85361960 |  |  | SCAND2P; NMB; AC103965.1; SEC11A; ZSCAN2; ALPK3; WDR73; UBE2Q2P1 | ALPK3 |
|  | V3 | rs8046873 | 16 | 82751932 |  |  |  | CDH13 |
|  | V1 | rs133890 | 22 | 26160161 |  |  | MYO18B | MYO18B |
| S-area | V4 | rs5016273 | 1 | 61881512 |  | Left bundle branch block; Congestive heart failure (CHF) NOS |  | NFIA |
|  | aVF | rs2207791 | 1 | 61894902 | QRS duration |  |  | NFIA |
|  | III | rs4915741 | 1 | 61897148 |  |  |  | NFIA |
|  | V2 | rs2207790 | 1 | 61897967 | QRS duration; Electrocardiographic conduction measures | Left bundle branch block |  | NFIA |
|  | V3 | rs11207739 | 1 | 61899519 |  | Left bundle branch block |  | NFIA |
|  | V5 | rs12145374 | 1 | 112480536 |  |  |  | KCND3 |
|  | V5 | rs75013985 | 1 | 112530430 | PR interval | Paroxysmal tachycardia, unspecified; Paroxysmal supraventricular tachycardia |  | KCND3 |
|  | V4 | rs116532272 | 1 | 112560237 | TPE interval (resting); PR interval | Paroxysmal tachycardia, unspecified; Paroxysmal supraventricular tachycardia |  |  |
|  | III | rs1873164 | 2 | 179753549 |  |  | SESTD1 | CCDC141 |
|  | V4 | rs6797133 | 3 | 38656033 |  | Diseases of hair and hair follicles; Sebaceous cyst; Diseases of sebaceous glands | ACVR2B; XYLB | SCN5A |
|  | V2 | rs13165478 | 5 | 153869040 | QRS complex; QRS duration; Global electrical heterogeneity phenotypes |  | SAP30L; GALNT10 |  |
|  | V4 | rs62379942 | 5 | 153871070 |  |  | SAP30L; GALNT10 |  |
|  | V3 | rs10054375 | 5 | 153871832 | Electrocardiographic traits; Electrocardiogram morphology |  | SAP30L; GALNT10 |  |
|  | V1 | rs10076436 | 5 | 153871841 | Global electrical heterogeneity phenotypes; QRS duration; Myocardial fractal dimension |  | SAP30L; GALNT10 |  |
|  | V4 | rs4151702 | 6 | 36645988 | Ascending aorta maximum area |  | CDKN1A; RAB44; KCTD20; ETV7; SRSF3 | DINOL |
|  | aVF; III | rs59365541 | 6 | 117471951 | Electrocardiogram morphology | Bundle branch block; Left bundle branch block; Inguinal hernia; Cardiac conduction disorders; Atrial fibrillation and flutter | DSE |  |
|  | V5 | rs1194743 | 10 | 54212597 | Electrocardiographic traits |  |  | LNCAROD |
|  | V1 | rs12776791 | 10 | 114489010 |  |  | RP11-57H14.4 | VTI1A |
|  | III | rs10885379 | 10 | 114494005 |  |  | RP11-57H14.4; VTI1A | VTI1A |
|  | V5 | rs7132593 | 12 | 115360866 |  |  |  |  |
|  | V4 | rs7487962 | 12 | 115361930 |  |  |  |  |
|  | V3 | rs3914956 | 12 | 115363751 | QRS duration |  |  |  |
|  | III | rs7132327 | 12 | 115381071 | QRS complex (12-leadsum); Myocardial fractal dimension; Electrocardiogram morphology |  |  |  |
| T-area | aVR | rs7638275 | 3 | 38665823 | QT dynamics during exercise; Global electrical heterogeneity phenotypes | Atrial fibrillation and flutter; Cardiac dysrhythmias |  | SCN5A |
|  | aVR | rs4963759 | 12 | 24586390 |  |  |  | SOX5 |
| J_up | I | rs2120436 | 1 | 112451447 | Atrial fibrillation/atrial flutter | Atrial fibrillation and flutter; Cardiac dysrhythmias |  | KCND3 |
|  | aVL | rs16866352 | 2 | 179368380 |  | Cardiac dysrhythmias; Atrial fibrillation and flutter | TTN | PLEKHA3 |
|  | I | rs6755784 | 2 | 179389742 | Left ventricular ejection fraction; Left ventricular mass; Left ventricular end-diastolic volume; Left ventricular end-systolic volume; Left ventricular global circumferential strain; Left ventricular global radial strain |  | PRKRA | TTN-AS1 |
|  | V5; V6; aVR | rs2562834 | 2 | 179580583 |  | Cardiac dysrhythmias; Atrial fibrillation and flutter | PRKRA; AC009948.5; TTN | TTN |
|  | V6; aVR | rs546095518 | 2 | 179595589 |  |  |  | TTN |
|  | II | rs11902709 | 2 | 179608207 |  |  | PRKRA; AC009948.5; SESTD1 | TTN |
|  | aVR | rs6715901 | 2 | 179650954 | Type 2 diabetes |  | PLEKHA3; SESTD1 | TTN |
|  | V6 | rs12464215 | 2 | 179690968 |  |  | PLEKHA3 |  |
|  | aVR | rs17362588 | 2 | 179721046 | Heart rate response to recovery post exercise; Diastolic blood pressure; Pulse pressure; QRS duration; Hypertrophic cardiomyopathy; Heart rate |  | DFNB59 | CCDC141 |
|  | aVR | rs1982717 | 2 | 213346989 |  | Benign neoplasm of other parts of digestive system |  | ERBB4 |
|  | aVR | rs6797133 | 3 | 38656033 |  | Diseases of hair and hair follicles; Sebaceous cyst; Diseases of sebaceous glands | ACVR2B; XYLB | SCN5A |
|  | I | rs55824920 | 3 | 38659952 |  | Diseases of hair and hair follicles; Sebaceous cyst; Diseases of sebaceous glands |  | SCN5A |
|  | I | rs6882776 | 5 | 172664163 | TPE interval (resting); Atrial fibrillation; Heart rate | Atrial fibrillation and flutter | BNIP1; CREBRF |  |
|  | aVR | rs6891790 | 5 | 172670745 | Atrial fibrillation; QT interval | Atrial fibrillation and flutter; Cardiac dysrhythmias | BNIP1; CREBRF |  |
|  | V6 | rs72825038 | 6 | 7527269 | PR interval; Electrocardiogram morphology |  |  |  |
|  | V5; V6 | rs12199346 | 6 | 36641546 |  |  | CDKN1A; RAB44; KCTD20; SRSF3 | PANDAR |
|  | aVL | rs12192632 | 6 | 117525985 | Systolic blood pressure | Left bundle branch block; Bundle branch block; Cardiac conduction disorders | DSE |  |
|  | I | rs281724 | 8 | 108622329 |  |  | ANGPT1 |  |
|  | III | rs78656993 | 10 | 33522809 |  |  |  | NRP1 |
|  | II | rs1888684 | 10 | 33622977 |  |  | NRP1 | NRP1 |
|  | II | rs10763928 | 10 | 33632954 |  |  | NRP1 |  |
|  | aVF | rs10827243 | 10 | 33633418 |  |  | NRP1 |  |
|  | III | rs10827246 | 10 | 33642629 |  |  | NRP1 |  |
|  | V6 | rs1194743 | 10 | 54212597 | Electrocardiographic traits |  |  | LNCAROD |
|  | II | rs1733724 | 10 | 54223977 | QRS duration; QRS complex; Electrocardiogram morphology |  |  | LNCAROD |
|  | V5 | rs17831429 | 10 | 112587774 |  |  |  | RBM20 |
|  | I | rs1886600 | 10 | 114490043 |  |  | RP11-57H14.4; VTI1A | VTI1A |
|  | aVL | rs1408817 | 10 | 114498145 |  |  | ACSL5 | VTI1A |
|  | V5; V6 | rs34972471 | 11 | 48050391 |  |  | PTPRJ; MYBPC3; ACP2; C1QTNF4; NR1H3; NUP160; DDB2; PSMC3; AGBL2; RP11-750H9.5; FNBP4; MTCH2 | PTPRJ |
|  | V6 | rs10850366 | 12 | 115144773 |  |  |  |  |
|  | aVR; I | rs7309985 | 12 | 115152129 |  |  |  |  |
|  | V5; II | rs1910046 | 12 | 115152444 |  |  |  |  |
|  | aVF | rs3914956 | 12 | 115363751 | QRS duration |  |  |  |
|  | V5; V6; II; III; aVR; I | rs7132327 | 12 | 115381071 | QRS complex (12-leadsum); Myocardial fractal dimension; Electrocardiogram morphology |  |  |  |
|  | V6 | rs3746435 | 20 | 33587198 | QRS duration |  | TRPC4AP; EDEM2; MYH7B; UQCC; MAP1LC3A; CPNE1; GGT7; EIF2S2; RP4-614O4.12; ITCH; PROCR; MMP24-AS1; CEP250; RBM39; GSS | MYH7B |
| R | V2 | rs112087357 | 1 | 26432370 |  | Hypercholesterolemia | PAFAH2; STMN1; RPS6KA1; CD52; CEP85; DHDDS; SH3BGRL3 |  |
|  | V3 | rs12142600 | 1 | 61886599 | Electrocardiographic traits | Left bundle branch block |  | NFIA |
|  | aVR | rs12090194 | 1 | 112454822 | P wave terminal force | Atrial fibrillation and flutter; Cardiac dysrhythmias; Secondary malignant neoplasm of skin |  | KCND3 |
|  | I | rs72694603 | 1 | 112458893 |  | Atrial fibrillation and flutter; Cardiac dysrhythmias; Secondary malignant neoplasm of skin |  | KCND3 |
|  | V5; V6 | rs72694622 | 1 | 112481667 | DNA methylation variation |  |  | KCND3 |
|  | I; aVR | rs75013985 | 1 | 112530430 | PR interval | Paroxysmal tachycardia, unspecified; Paroxysmal supraventricular tachycardia |  | KCND3 |
|  | V1 | rs6723399 | 2 | 179381715 | Supranormal left ventricular ejection fraction | Cardiac dysrhythmias; Atrial fibrillation and flutter | SESTD1; PRKRA; TTN; TTN-AS1 |  |
|  | III | rs13031826 | 2 | 179756602 |  | Atrioventricular [AV] block |  | CCDC141 |
|  | V1; V2 | rs35596070 | 2 | 179759692 | Heart rate response to recovery post exercise | Atrioventricular [AV] block | SESTD1 | CCDC141 |
|  | V2 | rs55754224 | 4 | 114428714 | Atrial fibrillation |  |  | CAMK2D |
|  | V2 | rs55679363 | 8 | 125867834 | Supranormal left ventricular ejection fraction; Hematocrit; Hemoglobin concentration; PR interval |  | MTSS1 |  |
|  | II; aVR | rs10763928 | 10 | 33632954 |  |  | NRP1 |  |
|  | V1 | rs72840788 | 10 | 121415685 | Parkinson's disease or first-degree relation to individual with Parkinson's disease; High-sensitivity cardiac troponin T levels; Left ventricular global radial strain; Left ventricular ejection fraction; Left ventricular end-diastolic volume; Left ventricular end-systolic volume; Left ventricular global circumferential strain; Free cholesterol to total lipids ratio in very large HDL; Hypertrophic cardiomyopathy; Left ventricular end-systolic volume; Left ventricular ejection fraction | Heart failure NOS; stress incontinence, female; Congestive heart failure; non-hypertensive | MCMBP; BAG3 | BAG3 |
|  | V2 | rs17617337 | 10 | 121426884 | Diastolic blood pressure; Left ventricular ejection fraction; Heart failure | Heart failure NOS; stress incontinence, female; Congestive heart failure; non-hypertensive | MCMBP; BAG3 | BAG3 |
|  | V2 | rs2958149 | 12 | 57109792 |  | Atrial fibrillation and flutter; Cardiac dysrhythmias | RBMS2; PRIM1; SPRYD4; BAZ2A; NACA; PTGES3 | NACA |
|  | V1 | rs12227117 | 12 | 57166413 |  | Atrial fibrillation and flutter; Cardiac dysrhythmias | RBMS2; PRIM1; SPRYD4; BAZ2A; NACA; PTGES3 | HSD17B6 |
|  | I; V6 | rs10850366 | 12 | 115144773 |  |  |  |  |
|  | V6 | rs4122458 | 12 | 115363954 |  |  |  |  |
|  | aVR | rs4767282 | 12 | 115369038 |  |  |  |  |
|  | V1 | rs7301677 | 12 | 115381147 | Global electrical heterogeneity phenotypes (QRS-T angle) |  |  |  |
|  | V1; V2; V3; V4 | rs4633690 | 15 | 85361960 |  |  | SCAND2P; NMB; AC103965.1; SEC11A; ZSCAN2; ALPK3; WDR73; UBE2Q2P1 | ALPK3 |
|  | V3 | rs8046873 | 16 | 82751932 |  |  |  | CDH13 |
|  | aVL | rs11874 | 17 | 45017193 | Electrocardiographic traits; Medication use (agents acting on the renin-angiotensin system); congenital heart disease (anomalies of thoracic arteries and veins) | Essential hypertension; Hypertension; Diverticulosis and diverticulitis; Diverticulosis | LRRC37A; NSFP1; ARL17B; FAM215B; NSF; TBKBP1 | GOSR2 |
|  | V1 | rs133885 | 22 | 26159289 | Mathematical ability in children with dyslexia; Left atrial total emptying fraction; Electrocardiogram morphology |  | MYO18B | MYO18B |
| RS | V3 | rs61775418 | 1 | 26393505 | Electrocardiogram morphology | Hypercholesterolemia | PAFAH2; STMN1; RPS6KA1; CD52; CEP85; SH3BGRL3; CNKSR1 | TRIM63 |
|  | III | rs4915741 | 1 | 61897148 |  |  |  | NFIA |
|  | aVR | rs2120436 | 1 | 112451447 | Atrial fibrillation/atrial flutter | Atrial fibrillation and flutter; Cardiac dysrhythmias |  | KCND3 |
|  | I | rs72694603 | 1 | 112458893 |  | Atrial fibrillation and flutter; Cardiac dysrhythmias; Secondary malignant neoplasm of skin |  | KCND3 |
|  | V4 | rs4839185 | 1 | 112460262 | P wave terminal force | Atrial fibrillation and flutter; Cardiac dysrhythmias; Secondary malignant neoplasm of skin |  | KCND3 |
|  | V5; V6 | rs72694622 | 1 | 112481667 | DNA methylation variation |  |  | KCND3 |
|  | V5; V6; I; aVR | rs75013985 | 1 | 112530430 | PR interval | Paroxysmal tachycardia, unspecified; Paroxysmal supraventricular tachycardia |  | KCND3 |
|  | V4 | rs2840167 | 2 | 36683316 |  | Bundle branch block | FEZ2; CRIM1 | CRIM1 |
|  | III | rs1873164 | 2 | 179753549 |  |  | SESTD1 | CCDC141 |
|  | V2 | rs13185595 | 5 | 153872170 | QRS duration; QRS complex |  | SAP30L; GALNT10 |  |
|  | aVF; III | rs12192632 | 6 | 117525985 | Systolic blood pressure | Left bundle branch block; Bundle branch block; Cardiac conduction disorders | DSE |  |
|  | aVF; II; III; aVR | rs10763928 | 10 | 33632954 |  |  | NRP1 |  |
|  | aVR | rs1194743 | 10 | 54212597 | Electrocardiographic traits |  |  | LNCAROD |
|  | V6; I | rs10850366 | 12 | 115144773 |  |  |  |  |
|  | III | rs7487962 | 12 | 115361930 |  |  |  |  |
|  | V6 | rs4122458 | 12 | 115363954 |  |  |  |  |
|  | V5 | rs7959283 | 12 | 115370538 |  |  |  |  |
|  | aVR | rs7980129 | 12 | 115372552 |  |  |  |  |
|  | aVF; II | rs7132327 | 12 | 115381071 | QRS complex (12-leadsum); Myocardial fractal dimension; Electrocardiogram morphology |  |  |  |
|  | V2; V3 | rs28595395 | 15 | 85334952 |  |  | SCAND2P; NMB; AC103965.1; SEC11A; ZSCAN2; ALPK3; WDR73; UBE2Q2P1 | ZNF592 |
|  | V4 | rs8039472 | 15 | 85361644 | Left ventricle wall thickness; Left ventricular mass to end-diastolic volume ratio; Left ventricular end-systolic volume; Left ventricular global circumferential strain |  | SCAND2P; AC103965.1; NMB; WDR73; ZSCAN2; SEC11A; ALPK3; UBE2Q2P1 | ALPK3 |
|  | aVL | rs11874 | 17 | 45017193 | Electrocardiographic traits; Medication use (agents acting on the renin-angiotensin system); congenital heart disease (anomalies of thoracic arteries and veins) | Essential hypertension; Hypertension; Diverticulosis and diverticulitis; Diverticulosis | LRRC37A; NSFP1; ARL17B; FAM215B; NSF; TBKBP1 | GOSR2 |
| S | III; aVF | rs4915741 | 1 | 61897148 |  |  |  | NFIA |
|  | II | rs2207790 | 1 | 61897967 | QRS duration; Electrocardiographic conduction measures | Left bundle branch block |  | NFIA |
|  | V4 | rs2941584 | 2 | 54881621 | Lumbar spine bone mineral density |  | SPTBN1; CLHC1; RTN4; AC093110.3; EML6 | SPTBN1 |
|  | III; aVF | rs1873164 | 2 | 179753549 |  |  | SESTD1 | CCDC141 |
|  | V1 | rs7700110 | 4 | 114439894 |  |  |  | CAMK2D |
|  | aVL | rs35132791 | 4 | 114456506 | TPE interval (resting) |  |  | CAMK2D |
|  | V2; V4 | rs13165478 | 5 | 153869040 | QRS complex; QRS duration; Global electrical heterogeneity phenotypes |  | SAP30L; GALNT10 |  |
|  | V3 | rs10054375 | 5 | 153871832 | Electrocardiographic traits; Electrocardiogram morphology |  | SAP30L; GALNT10 |  |
|  | V4; V5 | rs4151702 | 6 | 36645988 | Ascending aorta maximum area |  | CDKN1A; RAB44; KCTD20; ETV7; SRSF3 | DINOL |
|  | III; aVF | rs59365541 | 6 | 117471951 | Electrocardiogram morphology | Bundle branch block; Left bundle branch block; Inguinal hernia; Cardiac conduction disorders; Atrial fibrillation and flutter | DSE |  |
|  | V4 | rs13234350 | 7 | 35446584 |  |  |  |  |
|  | V5 | rs1194743 | 10 | 54212597 | Electrocardiographic traits |  |  | LNCAROD |
|  | V5 | rs10850366 | 12 | 115144773 |  |  |  |  |
|  | V4 | rs1910046 | 12 | 115152444 |  |  |  |  |
|  | V3; V4; V5 | rs3914956 | 12 | 115363751 | QRS duration |  |  |  |
|  | V6 | rs2384555 | 12 | 115367657 |  |  |  |  |
|  | II; III; aVF | rs7132327 | 12 | 115381071 | QRS complex (12-leadsum); Myocardial fractal dimension; Electrocardiogram morphology |  |  |  |
|  | V4 | rs806322 | 13 | 50841444 |  |  | DLEU1; DLEU7; RCBTB1; RP11-34F20.5 | DLEU1 |
|  | V4 | rs61989417 | 14 | 72010998 |  |  | SIPA1L1 | SIPA1L1 |
| ST | V1 | rs12566775 | 1 | 3226929 |  |  |  | PRDM16 |
|  | aVR | rs1763604 | 1 | 16339772 | Breast cancer, ovarian cancer or prostate cancer; Electrocardiogram morphology |  | RP11-169K16.9; CASP9; SPEN; RP4-798A10.2; ZBTB17 |  |
|  | aVF | rs2207790 | 1 | 61897967 | QRS duration; Electrocardiographic conduction measures | Left bundle branch block |  | NFIA |
|  | V3; V4 | rs2941584 | 2 | 54881621 | Lumbar spine bone mineral density |  | SPTBN1; CLHC1; RTN4; AC093110.3; EML6 | SPTBN1 |
|  | aVR | rs7573293 | 2 | 179753245 |  |  | SESTD1 | CCDC141 |
|  | III | rs1873164 | 2 | 179753549 |  |  | SESTD1 | CCDC141 |
|  | I | rs1489490 | 2 | 179764484 |  |  | PLEKHA3; CCDC141 | CCDC141 |
|  | aVL | rs11098193 | 4 | 114429407 |  |  |  | CAMK2D |
|  | V1 | rs56087422 | 4 | 114444394 |  |  |  | CAMK2D |
|  | V2; V4; V5 | rs13165478 | 5 | 153869040 | QRS complex; QRS duration; Global electrical heterogeneity phenotypes |  | SAP30L; GALNT10 |  |
|  | V3 | rs10054375 | 5 | 153871832 | Electrocardiographic traits; Electrocardiogram morphology |  | SAP30L; GALNT10 |  |
|  | V4; V5 | rs4151702 | 6 | 36645988 | Ascending aorta maximum area |  | CDKN1A; RAB44; KCTD20; ETV7; SRSF3 | DINOL |
|  | III | rs59365541 | 6 | 117471951 | Electrocardiogram morphology | Bundle branch block; Left bundle branch block; Inguinal hernia; Cardiac conduction disorders; Atrial fibrillation and flutter | DSE |  |
|  | aVF | rs1848404 | 6 | 117475647 |  | Inguinal hernia; Bundle branch block; Left bundle branch block; Cardiac conduction disorders; Atrial fibrillation and flutter | DSE |  |
|  | V5 | rs6416327 | 12 | 115361364 |  |  |  |  |
|  | V4 | rs7487962 | 12 | 115361930 |  |  |  |  |
|  | V3 | rs3914956 | 12 | 115363751 | QRS duration |  |  |  |
|  | aVF; III | rs7132327 | 12 | 115381071 | QRS complex (12-leadsum); Myocardial fractal dimension; Electrocardiogram morphology |  |  |  |
|  | aVL | rs7301677 | 12 | 115381147 | Global electrical heterogeneity phenotypes (QRS-T angle) |  |  |  |
|  | I | rs10850409 | 12 | 115381740 | PR segment duration; QRS complex; QRS duration |  |  |  |
|  | V4 | rs61989382 | 14 | 71983638 | PR interval | Benign neoplasm of other parts of digestive system | SIPA1L1 | SIPA1L1 |
| T | I | rs846109 | 1 | 6289835 |  |  |  | ICMT |
|  | V2 | rs10918571 | 1 | 162015277 |  |  |  |  |
|  | V1; V3; V4 | rs12036340 | 1 | 162015740 | QRS complex (12-leadsum) |  |  |  |
|  | II | rs7638275 | 3 | 38665823 | QT dynamics during exercise; Global electrical heterogeneity phenotypes | Atrial fibrillation and flutter; Cardiac dysrhythmias |  | SCN5A |
|  | V2 | rs3951016 | 6 | 118559658 | Resting heart rate; Atrial fibrillation; QRS duration | Diabetic retinopathy |  | SLC35F1 |
|  | V1; V2; V3; V4 | rs2074238 | 11 | 2484803 | QT interval; T wave morphology restitution during exercise; T wave morphology restitution during recovery from exercise; QT dynamics during exercise; Electrocardiogram morphology |  |  | KCNQ1 |
|  | V5; V6; aVR | rs7307613 | 12 | 24595192 | T wave morphology restitution during exercise; Electrocardiogram morphology |  |  | SOX5 |
|  | I | rs7957437 | 12 | 24616209 |  |  |  | SOX5 |
|  | aVR | rs7132327 | 12 | 115381071 | QRS complex (12-leadsum); Myocardial fractal dimension; Electrocardiogram morphology |  |  |  |
|  | I | rs7301677 | 12 | 115381147 | Global electrical heterogeneity phenotypes (QRS-T angle) |  |  |  |
| t_QT | aVR | rs2120436 | 1 | 112451447 | Atrial fibrillation/atrial flutter | Atrial fibrillation and flutter; Cardiac dysrhythmias |  | KCND3 |
|  | aVR | rs12036340 | 1 | 162015740 | QRS complex (12-leadsum) |  |  |  |
|  | aVR | rs1910046 | 12 | 115152444 |  |  |  |  |
|  | aVR | rs4767282 | 12 | 115369038 |  |  |  |  |
|  | aVF | rs7959283 | 12 | 115370538 |  |  |  |  |
|  | II | rs10850409 | 12 | 115381740 | PR segment duration; QRS complex; QRS duration |  |  |  |
| t_R | V1 | rs1997571 | 7 | 116198621 | Heart rate response to recovery post exercise; QT interval; Electrocardiogram morphology | Atrial fibrillation and flutter | TES | CAV1 |
|  | V6 | rs1896356 | 12 | 115367286 |  |  |  |  |
|  | V5 | rs7132327 | 12 | 115381071 | QRS complex (12-leadsum); Myocardial fractal dimension; Electrocardiogram morphology |  |  |  |
| t_S | V4 | rs72692602 | 1 | 112458833 |  | Atrial fibrillation and flutter; Cardiac dysrhythmias; Secondary malignant neoplasm of skin |  | KCND3 |
|  | V5 | rs72694622 | 1 | 112481667 | DNA methylation variation |  |  | KCND3 |
|  | V3; V4; V5 | rs75013985 | 1 | 112530430 | PR interval | Paroxysmal tachycardia, unspecified; Paroxysmal supraventricular tachycardia |  | KCND3 |
|  | V6 | rs6416327 | 12 | 115361364 |  |  |  |  |
|  | V2; V3 | rs4784934 | 16 | 58459926 | QT interval; Electrocardiographic traits |  | RP11-481J2.2; GINS3 |  |
|  | V5 | rs28623612 | 16 | 58467237 |  |  | GINS3; RP11-481J2.2 |  |
|  | V4 | rs28627526 | 16 | 58467739 |  |  | RP11-481J2.2; GINS3 |  |
|  | V4 | rs2223036 | 21 | 30604782 |  | Congestive heart failure; non-hypertensive; Heart failure NOS; Other forms of chronic heart disease | LINC00189; GAPDHP14; C21orf7; BACH1; GRIK1; N6AMT1 | LINC00189 |
| t_ST | I | rs11102347 | 1 | 112391809 |  |  |  | KCND3 |
|  | aVR | rs2120436 | 1 | 112451447 | Atrial fibrillation/atrial flutter | Atrial fibrillation and flutter; Cardiac dysrhythmias |  | KCND3 |
|  | I | rs72692597 | 1 | 112455442 |  | Atrial fibrillation and flutter; Cardiac dysrhythmias; Secondary malignant neoplasm of skin |  | KCND3 |
|  | I | rs7309985 | 12 | 115152129 |  |  |  |  |
|  | aVR | rs1910046 | 12 | 115152444 |  |  |  |  |
|  | V5; V6 | rs9630280 | 12 | 115361757 |  |  |  |  |
|  | aVR | rs7132327 | 12 | 115381071 | QRS complex (12-leadsum); Myocardial fractal dimension; Electrocardiogram morphology |  |  |  |
|  | aVF; II | rs7301677 | 12 | 115381147 | Global electrical heterogeneity phenotypes (QRS-T angle) |  |  |  |
|  | III | rs10850409 | 12 | 115381740 | PR segment duration; QRS complex; QRS duration |  |  |  |

^a^: (P<0.05/1419[3.5e-5])

Supplementary Table 5. The related genes (Bonferroni corrected P-value < 0.05) for ETCs analyzed by gene-based genome-wide association analysis using MAGMA.

| ETC | GENE symbol | GENE id | Strand | CHR | START | STOP | P value | FDR | Bonferroni corrected P |
| --- | --- | --- | --- | --- | --- | --- | --- | --- | --- |
| I_R | MFHAS1 | 9258 | - | 8 | 8640864 | 8751131 | 2.46E-06 | 9.25E-03 | 4.62E-02 |
| I_R | XKR6 | 286046 | - | 8 | 10753654 | 11058875 | 1.65E-07 | 1.03E-03 | 3.09E-03 |
| I_R | KCND3 | 3752 | - | 1 | 112318444 | 112532147 | 6.45E-18 | 1.21E-13 | 1.21E-13 |
| I_R | MSRA | 4482 | + | 8 | 9911830 | 10286401 | 1.77E-06 | 8.29E-03 | 3.32E-02 |
| I_R | LOC643355 | 643355 | + | 1 | 112532593 | 112541464 | 1.02E-07 | 9.61E-04 | 1.92E-03 |
| aVF_RS | NRP1 | 8829 | - | 10 | 33466419 | 33623833 | 6.84E-10 | 1.28E-05 | 1.28E-05 |
| aVF_RS | ADK | 132 | + | 10 | 75910943 | 76469061 | 1.63E-09 | 1.53E-05 | 3.06E-05 |
| aVF_RS | CRIM1 | 51232 | + | 2 | 36583370 | 36778278 | 3.68E-07 | 2.30E-03 | 6.90E-03 |
| aVF_RS | AP3M1 | 26985 | - | 10 | 75880015 | 75910826 | 4.91E-07 | 2.30E-03 | 9.22E-03 |
| aVF_ST | NRP1 | 8829 | - | 10 | 33466419 | 33623833 | 9.98E-08 | 1.87E-03 | 1.87E-03 |
| aVF_ST | SPTBN1 | 6711 | + | 2 | 54683454 | 54898583 | 2.16E-06 | 1.05E-02 | 4.05E-02 |
| aVF_ST | SIPA1L1 | 26037 | + | 14 | 71788108 | 72207761 | 1.63E-06 | 1.05E-02 | 3.05E-02 |
| aVF_ST | FBN2 | 2201 | - | 5 | 127593601 | 127873735 | 2.23E-06 | 1.05E-02 | 4.19E-02 |
| aVF_J_up | CCDC141 | 285025 | - | 2 | 179694484 | 179914841 | 1.08E-07 | 6.77E-04 | 2.03E-03 |
| aVF_J_up | PRDM6 | 93166 | + | 5 | 122424841 | 122529960 | 2.80E-08 | 2.62E-04 | 5.25E-04 |
| aVF_J_up | CERS3 | 204219 | - | 15 | 100940600 | 101084925 | 1.74E-07 | 8.16E-04 | 3.26E-03 |
| aVF_J_up | TTN | 7273 | - | 2 | 179390716 | 179672150 | 8.35E-07 | 3.13E-03 | 1.57E-02 |
| aVF_J_up | NRP1 | 8829 | - | 10 | 33466419 | 33623833 | 3.33E-13 | 6.26E-09 | 6.26E-09 |
| aVF_R-area | ADK | 132 | + | 10 | 75910943 | 76469061 | 2.55E-06 | 4.78E-02 | 4.78E-02 |
| aVF_S-area | NRP1 | 8829 | - | 10 | 33466419 | 33623833 | 5.60E-07 | 1.05E-02 | 1.05E-02 |
| aVF_t_R | ISLR2 | 57611 | + | 15 | 74420544 | 74429143 | 2.51E-06 | 4.71E-02 | 4.71E-02 |
| aVF_t_ST | TKT | 7086 | - | 3 | 53258723 | 53290130 | 1.49E-06 | 1.40E-02 | 2.80E-02 |
| aVF_t_ST | NRP1 | 8829 | - | 10 | 33466419 | 33623833 | 4.48E-09 | 8.40E-05 | 8.40E-05 |
| aVF_t_QT | TKT | 7086 | - | 3 | 53258723 | 53290130 | 2.57E-07 | 4.82E-03 | 4.82E-03 |
| V1_R | NMB | 4828 | - | 15 | 85198360 | 85201802 | 4.26E-12 | 1.14E-08 | 7.99E-08 |
| V1_R | PRKRA | 8575 | - | 2 | 179296141 | 179315958 | 8.40E-07 | 7.17E-04 | 1.58E-02 |
| V1_R | SIPA1L1 | 26037 | + | 14 | 71788108 | 72207761 | 6.12E-08 | 6.76E-05 | 1.15E-03 |
| V1_R | PTGES3 | 10728 | - | 12 | 57057125 | 57082138 | 6.09E-11 | 9.52E-08 | 1.14E-06 |
| V1_R | 6-Sep | 23157 | - | X | 118749687 | 118827333 | 1.45E-06 | 1.19E-03 | 2.73E-02 |
| V1_R | PRIM1 | 5557 | - | 12 | 57125364 | 57146146 | 5.60E-09 | 7.00E-06 | 1.05E-04 |
| V1_R | ALPK3 | 57538 | + | 15 | 85359911 | 85416713 | 4.21E-13 | 1.32E-09 | 7.91E-09 |
| V1_R | ZNF592 | 9640 | + | 15 | 85291818 | 85349663 | 9.15E-12 | 2.00E-08 | 1.72E-07 |
| V1_R | SH3PXD2A | 9644 | - | 10 | 105353784 | 105615195 | 4.02E-07 | 3.59E-04 | 7.54E-03 |
| V1_R | ATP5B | 506 | - | 12 | 57031959 | 57039852 | 7.74E-10 | 1.12E-06 | 1.45E-05 |
| V1_R | PLEKHA3 | 65977 | + | 2 | 179345199 | 179369783 | 4.73E-09 | 6.33E-06 | 8.87E-05 |
| V1_R | BAG3 | 9531 | + | 10 | 121410859 | 121437331 | 2.39E-13 | 8.98E-10 | 4.49E-09 |
| V1_R | CCDC141 | 285025 | - | 2 | 179694484 | 179914841 | 2.46E-17 | 4.62E-13 | 4.62E-13 |
| V1_R | SOX8 | 30812 | + | 16 | 1031808 | 1036979 | 3.87E-07 | 3.59E-04 | 7.26E-03 |
| V1_R | ZSCAN2 | 54993 | + | 15 | 85144249 | 85166947 | 9.60E-12 | 2.00E-08 | 1.80E-07 |
| V1_R | NACA | 4666 | - | 12 | 57106211 | 57119326 | 1.29E-11 | 2.43E-08 | 2.43E-07 |
| V1_R | FKBP7 | 51661 | - | 2 | 179328391 | 179343646 | 2.05E-07 | 2.14E-04 | 3.85E-03 |
| V1_R | WDR73 | 84942 | - | 15 | 85185607 | 85197521 | 1.74E-11 | 2.98E-08 | 3.27E-07 |
| V1_R | SEC11A | 23478 | - | 15 | 85212768 | 85259691 | 4.22E-14 | 1.98E-10 | 7.92E-10 |
| V1_R | LMF1 | 64788 | - | 16 | 903634 | 1031318 | 2.49E-08 | 2.92E-05 | 4.67E-04 |
| V1_R | HSD17B6 | 8630 | + | 12 | 57146237 | 57181574 | 3.43E-14 | 1.98E-10 | 6.43E-10 |
| V1_R | DFNB59 | 494513 | + | 2 | 179316163 | 179326149 | 2.22E-07 | 2.19E-04 | 4.16E-03 |
| V1_R | TTN | 7273 | - | 2 | 179390716 | 179672150 | 4.85E-15 | 4.55E-11 | 9.10E-11 |
| V1_S | CAMK2D | 817 | - | 4 | 114372188 | 114683669 | 4.77E-07 | 2.29E-03 | 8.95E-03 |
| V1_S | NRP1 | 8829 | - | 10 | 33466419 | 33623833 | 9.76E-07 | 2.29E-03 | 1.83E-02 |
| V1_S | NUP160 | 23279 | - | 11 | 47799670 | 47870057 | 9.18E-07 | 2.29E-03 | 1.72E-02 |
| V1_S | VTI1A | 143187 | + | 10 | 114206756 | 114578503 | 1.36E-07 | 2.29E-03 | 2.55E-03 |
| V1_S | WDPCP | 51057 | - | 2 | 63348518 | 63815867 | 9.16E-07 | 2.29E-03 | 1.72E-02 |
| V1_S | FNBP4 | 23360 | - | 11 | 47738055 | 47789012 | 1.97E-06 | 4.11E-03 | 3.70E-02 |
| V1_S | PRDM16 | 63976 | + | 1 | 2985565 | 3355185 | 6.33E-07 | 2.29E-03 | 1.19E-02 |
| V1_S | BNIP1 | 662 | + | 5 | 172571445 | 172591390 | 3.98E-07 | 2.29E-03 | 7.46E-03 |
| V1_S | CREBRF | 153222 | + | 5 | 172483355 | 172566291 | 9.78E-07 | 2.29E-03 | 1.84E-02 |
| V1_T | NOS1AP | 9722 | + | 1 | 162039581 | 162339813 | 1.09E-12 | 2.04E-08 | 2.04E-08 |
| V1_RS | TTN | 7273 | - | 2 | 179390716 | 179672150 | 1.51E-08 | 1.42E-04 | 2.83E-04 |
| V1_RS | NRP1 | 8829 | - | 10 | 33466419 | 33623833 | 1.05E-08 | 1.42E-04 | 1.98E-04 |
| V1_RS | BNIP1 | 662 | + | 5 | 172571445 | 172591390 | 2.45E-06 | 1.53E-02 | 4.60E-02 |
| V1_ST | VTI1A | 143187 | + | 10 | 114206756 | 114578503 | 2.48E-06 | 1.55E-02 | 4.65E-02 |
| V1_ST | CAMK2D | 817 | - | 4 | 114372188 | 114683669 | 1.05E-07 | 1.97E-03 | 1.97E-03 |
| V1_ST | PRDM16 | 63976 | + | 1 | 2985565 | 3355185 | 2.37E-06 | 1.55E-02 | 4.44E-02 |
| V1_R-area | PRIM1 | 5557 | - | 12 | 57125364 | 57146146 | 6.44E-08 | 1.21E-04 | 1.21E-03 |
| V1_R-area | WDR73 | 84942 | - | 15 | 85185607 | 85197521 | 1.11E-07 | 1.90E-04 | 2.09E-03 |
| V1_R-area | NACA | 4666 | - | 12 | 57106211 | 57119326 | 2.45E-10 | 2.30E-06 | 4.60E-06 |
| V1_R-area | ATP5B | 506 | - | 12 | 57031959 | 57039852 | 3.73E-09 | 1.00E-05 | 7.00E-05 |
| V1_R-area | SEC11A | 23478 | - | 15 | 85212768 | 85259691 | 3.75E-09 | 1.00E-05 | 7.03E-05 |
| V1_R-area | ZSCAN2 | 54993 | + | 15 | 85144249 | 85166947 | 1.47E-07 | 2.20E-04 | 2.76E-03 |
| V1_R-area | HSD17B6 | 8630 | + | 12 | 57146237 | 57181574 | 6.19E-13 | 1.16E-08 | 1.16E-08 |
| V1_R-area | CCDC141 | 285025 | - | 2 | 179694484 | 179914841 | 3.00E-09 | 1.00E-05 | 5.62E-05 |
| V1_R-area | TTN | 7273 | - | 2 | 179390716 | 179672150 | 2.58E-06 | 3.42E-03 | 4.84E-02 |
| V1_R-area | PTGES3 | 10728 | - | 12 | 57057125 | 57082138 | 1.18E-09 | 7.40E-06 | 2.22E-05 |
| V1_R-area | BAG3 | 9531 | + | 10 | 121410859 | 121437331 | 5.60E-08 | 1.17E-04 | 1.05E-03 |
| V1_R-area | ALPK3 | 57538 | + | 15 | 85359911 | 85416713 | 1.53E-07 | 2.20E-04 | 2.86E-03 |
| V1_R-area | ZNF592 | 9640 | + | 15 | 85291818 | 85349663 | 1.74E-09 | 8.17E-06 | 3.27E-05 |
| V1_R-area | NMB | 4828 | - | 15 | 85198360 | 85201802 | 5.63E-08 | 1.17E-04 | 1.06E-03 |
| V1_S-area | VTI1A | 143187 | + | 10 | 114206756 | 114578503 | 8.80E-10 | 1.65E-05 | 1.65E-05 |
| V1_S-area | HAND1 | 9421 | - | 5 | 153854532 | 153857824 | 1.78E-06 | 1.67E-02 | 3.34E-02 |
| V1_t_T | PNRC2 | 55629 | + | 1 | 24286301 | 24289952 | 2.97E-07 | 5.56E-03 | 5.56E-03 |
| V1_t_R | SCN10A | 6336 | - | 3 | 38738837 | 38835501 | 2.95E-09 | 5.53E-05 | 5.53E-05 |
| V1_t_R | SCN5A | 6331 | - | 3 | 38589553 | 38691164 | 6.02E-08 | 3.76E-04 | 1.13E-03 |
| V1_t_R | CAV1 | 857 | + | 7 | 116164839 | 116201239 | 4.82E-08 | 3.76E-04 | 9.04E-04 |
| V1_t_QT | CAV1 | 857 | + | 7 | 116164839 | 116201239 | 4.75E-07 | 8.92E-03 | 8.92E-03 |
| I_t_R | LPCAT2 | 54947 | + | 16 | 55542913 | 55620582 | 2.66E-06 | 4.99E-02 | 4.99E-02 |
| V2_R | ZSCAN2 | 54993 | + | 15 | 85144249 | 85166947 | 1.72E-13 | 5.39E-10 | 3.23E-09 |
| V2_R | CAMK2D | 817 | - | 4 | 114372188 | 114683669 | 2.58E-07 | 2.69E-04 | 4.84E-03 |
| V2_R | LMF1 | 64788 | - | 16 | 903634 | 1031318 | 1.08E-08 | 1.69E-05 | 2.03E-04 |
| V2_R | PTGES3 | 10728 | - | 12 | 57057125 | 57082138 | 1.68E-09 | 3.51E-06 | 3.16E-05 |
| V2_R | NMB | 4828 | - | 15 | 85198360 | 85201802 | 3.22E-15 | 2.01E-11 | 6.04E-11 |
| V2_R | PDIK1L | 149420 | + | 1 | 26437656 | 26452039 | 1.53E-07 | 1.69E-04 | 2.87E-03 |
| V2_R | VTI1A | 143187 | + | 10 | 114206756 | 114578503 | 1.62E-06 | 1.38E-03 | 3.04E-02 |
| V2_R | MEF2D | 4209 | - | 1 | 156433513 | 156470634 | 6.85E-08 | 8.57E-05 | 1.29E-03 |
| V2_R | HSD17B6 | 8630 | + | 12 | 57146237 | 57181574 | 7.68E-13 | 2.06E-09 | 1.44E-08 |
| V2_R | NACA | 4666 | - | 12 | 57106211 | 57119326 | 2.38E-09 | 4.47E-06 | 4.47E-05 |
| V2_R | PRIM1 | 5557 | - | 12 | 57125364 | 57146146 | 4.46E-07 | 4.19E-04 | 8.37E-03 |
| V2_R | TRIM63 | 84676 | - | 1 | 26377792 | 26394142 | 8.39E-08 | 9.84E-05 | 1.57E-03 |
| V2_R | SOX8 | 30812 | + | 16 | 1031808 | 1036979 | 5.85E-08 | 7.84E-05 | 1.10E-03 |
| V2_R | ALPK3 | 57538 | + | 15 | 85359911 | 85416713 | 2.22E-16 | 4.17E-12 | 4.17E-12 |
| V2_R | WDR73 | 84942 | - | 15 | 85185607 | 85197521 | 1.44E-14 | 6.77E-11 | 2.71E-10 |
| V2_R | BAG3 | 9531 | + | 10 | 121410859 | 121437331 | 4.38E-08 | 6.32E-05 | 8.22E-04 |
| V2_R | TTN | 7273 | - | 2 | 179390716 | 179672150 | 1.49E-06 | 1.33E-03 | 2.79E-02 |
| V2_R | SH3BGRL3 | 83442 | + | 1 | 26606213 | 26608013 | 3.50E-07 | 3.46E-04 | 6.57E-03 |
| V2_R | SEC11A | 23478 | - | 15 | 85212768 | 85259691 | 2.75E-14 | 1.03E-10 | 5.17E-10 |
| V2_R | ZNF592 | 9640 | + | 15 | 85291818 | 85349663 | 2.05E-15 | 1.93E-11 | 3.85E-11 |
| V2_R | CCDC141 | 285025 | - | 2 | 179694484 | 179914841 | 2.90E-09 | 4.95E-06 | 5.45E-05 |
| V2_R | ATP5B | 506 | - | 12 | 57031959 | 57039852 | 4.70E-11 | 1.10E-07 | 8.81E-07 |
| V2_S | HAND1 | 9421 | - | 5 | 153854532 | 153857824 | 3.01E-13 | 5.64E-09 | 5.64E-09 |
| V2_S | NRP1 | 8829 | - | 10 | 33466419 | 33623833 | 1.71E-07 | 1.07E-03 | 3.20E-03 |
| V2_S | SAP30L | 79685 | + | 5 | 153825517 | 153840613 | 2.30E-11 | 2.16E-07 | 4.31E-07 |
| V2_S | SPTBN1 | 6711 | + | 2 | 54683454 | 54898583 | 5.33E-07 | 2.50E-03 | 1.00E-02 |
| V2_T | SLC35F1 | 222553 | + | 6 | 118228689 | 118638839 | 4.35E-07 | 2.04E-03 | 8.16E-03 |
| V2_T | CEP85L | 387119 | - | 6 | 118781935 | 119031238 | 1.13E-07 | 1.06E-03 | 2.11E-03 |
| V2_T | NOS1AP | 9722 | + | 1 | 162039581 | 162339813 | 2.15E-18 | 4.04E-14 | 4.04E-14 |
| V2_T | CNOT1 | 23019 | - | 16 | 58553850 | 58663790 | 3.15E-07 | 1.97E-03 | 5.91E-03 |
| V2_RS | ATP5B | 506 | - | 12 | 57031959 | 57039852 | 1.04E-06 | 2.43E-03 | 1.95E-02 |
| V2_RS | WDR73 | 84942 | - | 15 | 85185607 | 85197521 | 8.98E-07 | 2.41E-03 | 1.68E-02 |
| V2_RS | NRP1 | 8829 | - | 10 | 33466419 | 33623833 | 2.10E-08 | 1.97E-04 | 3.94E-04 |
| V2_RS | HSD17B6 | 8630 | + | 12 | 57146237 | 57181574 | 7.21E-07 | 2.25E-03 | 1.35E-02 |
| V2_RS | NMB | 4828 | - | 15 | 85198360 | 85201802 | 3.69E-07 | 1.39E-03 | 6.93E-03 |
| V2_RS | SEC11A | 23478 | - | 15 | 85212768 | 85259691 | 5.91E-08 | 3.70E-04 | 1.11E-03 |
| V2_RS | ALPK3 | 57538 | + | 15 | 85359911 | 85416713 | 7.89E-08 | 3.70E-04 | 1.48E-03 |
| V2_RS | ZSCAN2 | 54993 | + | 15 | 85144249 | 85166947 | 1.22E-06 | 2.54E-03 | 2.29E-02 |
| V2_RS | ZNF592 | 9640 | + | 15 | 85291818 | 85349663 | 2.02E-09 | 3.79E-05 | 3.79E-05 |
| V2_ST | HAND1 | 9421 | - | 5 | 153854532 | 153857824 | 3.17E-12 | 5.96E-08 | 5.96E-08 |
| V2_ST | SAP30L | 79685 | + | 5 | 153825517 | 153840613 | 7.17E-10 | 6.73E-06 | 1.35E-05 |
| V2_ST | SPTBN1 | 6711 | + | 2 | 54683454 | 54898583 | 2.85E-07 | 1.78E-03 | 5.35E-03 |
| I_t_ST | MSRA | 4482 | + | 8 | 9911830 | 10286401 | 1.75E-07 | 1.64E-03 | 3.28E-03 |
| I_t_ST | KCND3 | 3752 | - | 1 | 112318444 | 112532147 | 7.88E-15 | 1.48E-10 | 1.48E-10 |
| I_t_ST | CASQ2 | 845 | - | 1 | 116242624 | 116311426 | 1.50E-06 | 9.40E-03 | 2.82E-02 |
| V2_R-area | ZSCAN2 | 54993 | + | 15 | 85144249 | 85166947 | 4.58E-10 | 1.43E-06 | 8.60E-06 |
| V2_R-area | SPTBN1 | 6711 | + | 2 | 54683454 | 54898583 | 1.21E-08 | 2.53E-05 | 2.28E-04 |
| V2_R-area | PFKFB2 | 5208 | + | 1 | 207207761 | 207254368 | 1.85E-06 | 2.05E-03 | 3.47E-02 |
| V2_R-area | WDR73 | 84942 | - | 15 | 85185607 | 85197521 | 5.59E-11 | 2.10E-07 | 1.05E-06 |
| V2_R-area | NACA | 4666 | - | 12 | 57106211 | 57119326 | 7.25E-07 | 1.13E-03 | 1.36E-02 |
| V2_R-area | TRIM63 | 84676 | - | 1 | 26377792 | 26394142 | 1.86E-06 | 2.05E-03 | 3.49E-02 |
| V2_R-area | SEC11A | 23478 | - | 15 | 85212768 | 85259691 | 1.71E-11 | 1.04E-07 | 3.20E-07 |
| V2_R-area | CCDC141 | 285025 | - | 2 | 179694484 | 179914841 | 9.33E-07 | 1.25E-03 | 1.75E-02 |
| V2_R-area | CD34 | 947 | - | 1 | 208059883 | 208084742 | 8.40E-07 | 1.21E-03 | 1.58E-02 |
| V2_R-area | CAMK2D | 817 | - | 4 | 114372188 | 114683669 | 1.09E-09 | 2.56E-06 | 2.05E-05 |
| V2_R-area | ALPK3 | 57538 | + | 15 | 85359911 | 85416713 | 2.23E-11 | 1.04E-07 | 4.18E-07 |
| V2_R-area | ZNF592 | 9640 | + | 15 | 85291818 | 85349663 | 5.73E-12 | 5.38E-08 | 1.08E-07 |
| V2_R-area | LMF1 | 64788 | - | 16 | 903634 | 1031318 | 1.46E-06 | 1.82E-03 | 2.73E-02 |
| V2_R-area | NMB | 4828 | - | 15 | 85198360 | 85201802 | 1.72E-12 | 3.23E-08 | 3.23E-08 |
| V2_R-area | ATP5B | 506 | - | 12 | 57031959 | 57039852 | 1.11E-07 | 2.09E-04 | 2.09E-03 |
| V2_R-area | PDIK1L | 149420 | + | 1 | 26437656 | 26452039 | 2.08E-06 | 2.17E-03 | 3.91E-02 |
| V2_R-area | PTGES3 | 10728 | - | 12 | 57057125 | 57082138 | 5.52E-07 | 9.41E-04 | 1.04E-02 |
| V2_R-area | HSD17B6 | 8630 | + | 12 | 57146237 | 57181574 | 5.95E-10 | 1.59E-06 | 1.12E-05 |
| V2_S-area | SAP30L | 79685 | + | 5 | 153825517 | 153840613 | 2.86E-09 | 1.84E-05 | 5.37E-05 |
| V2_S-area | HAND1 | 9421 | - | 5 | 153854532 | 153857824 | 2.02E-11 | 3.79E-07 | 3.79E-07 |
| V2_S-area | NFIA | 4774 | + | 1 | 61542946 | 61928460 | 8.45E-07 | 3.97E-03 | 1.59E-02 |
| V2_S-area | SCN5A | 6331 | - | 3 | 38589553 | 38691164 | 2.94E-09 | 1.84E-05 | 5.52E-05 |
| V2_t_S | GALNT8 | 26290 | + | 12 | 4829752 | 4881892 | 2.13E-07 | 4.00E-03 | 4.00E-03 |
| I_t_QT | CASQ2 | 845 | - | 1 | 116242624 | 116311426 | 1.52E-08 | 2.85E-04 | 2.85E-04 |
| V3_R | WDPCP | 51057 | - | 2 | 63348518 | 63815867 | 1.68E-06 | 2.10E-03 | 3.15E-02 |
| V3_R | MEF2D | 4209 | - | 1 | 156433513 | 156470634 | 2.15E-07 | 4.47E-04 | 4.02E-03 |
| V3_R | ALPK3 | 57538 | + | 15 | 85359911 | 85416713 | 5.00E-10 | 1.56E-06 | 9.38E-06 |
| V3_R | ATP5B | 506 | - | 12 | 57031959 | 57039852 | 8.20E-07 | 1.18E-03 | 1.54E-02 |
| V3_R | PTGES3 | 10728 | - | 12 | 57057125 | 57082138 | 2.30E-06 | 2.69E-03 | 4.31E-02 |
| V3_R | NFIA | 4774 | + | 1 | 61542946 | 61928460 | 4.52E-08 | 1.21E-04 | 8.48E-04 |
| V3_R | CATSPER4 | 378807 | + | 1 | 26517119 | 26529033 | 7.62E-07 | 1.18E-03 | 1.43E-02 |
| V3_R | TRIM63 | 84676 | - | 1 | 26377792 | 26394142 | 3.03E-07 | 5.68E-04 | 5.68E-03 |
| V3_R | CCDC141 | 285025 | - | 2 | 179694484 | 179914841 | 1.06E-06 | 1.42E-03 | 1.99E-02 |
| V3_R | ZNF592 | 9640 | + | 15 | 85291818 | 85349663 | 5.00E-10 | 1.56E-06 | 9.38E-06 |
| V3_R | HSD17B6 | 8630 | + | 12 | 57146237 | 57181574 | 5.86E-08 | 1.37E-04 | 1.10E-03 |
| V3_R | ZSCAN2 | 54993 | + | 15 | 85144249 | 85166947 | 1.09E-12 | 5.10E-09 | 2.04E-08 |
| V3_R | WDR73 | 84942 | - | 15 | 85185607 | 85197521 | 1.55E-13 | 1.25E-09 | 2.90E-09 |
| V3_R | NMB | 4828 | - | 15 | 85198360 | 85201802 | 7.27E-15 | 1.36E-10 | 1.36E-10 |
| V3_R | PDIK1L | 149420 | + | 1 | 26437656 | 26452039 | 3.43E-07 | 5.85E-04 | 6.43E-03 |
| V3_R | SEC11A | 23478 | - | 15 | 85212768 | 85259691 | 2.00E-13 | 1.25E-09 | 3.74E-09 |
| V3_S | NDUFS3 | 4722 | + | 11 | 47600562 | 47606115 | 6.52E-09 | 3.27E-05 | 1.22E-04 |
| V3_S | SIPA1L1 | 26037 | + | 14 | 71788108 | 72207761 | 6.88E-08 | 1.43E-04 | 1.29E-03 |
| V3_S | FNBP4 | 23360 | - | 11 | 47738055 | 47789012 | 1.38E-08 | 4.32E-05 | 2.59E-04 |
| V3_S | RAPSN | 5913 | - | 11 | 47459308 | 47470730 | 1.43E-07 | 2.68E-04 | 2.68E-03 |
| V3_S | NR1H3 | 10062 | + | 11 | 47269851 | 47290584 | 6.95E-07 | 1.09E-03 | 1.30E-02 |
| V3_S | NUP160 | 23279 | - | 11 | 47799670 | 47870057 | 1.37E-08 | 4.32E-05 | 2.57E-04 |
| V3_S | SAP30L | 79685 | + | 5 | 153825517 | 153840613 | 4.44E-15 | 4.17E-11 | 8.33E-11 |
| V3_S | MTCH2 | 23788 | - | 11 | 47638858 | 47664206 | 3.66E-08 | 8.58E-05 | 6.86E-04 |
| V3_S | AGBL2 | 79841 | - | 11 | 47681143 | 47736928 | 3.55E-08 | 8.58E-05 | 6.67E-04 |
| V3_S | SLC39A13 | 91252 | + | 11 | 47428683 | 47438051 | 2.65E-06 | 3.55E-03 | 4.97E-02 |
| V3_S | CELF1 | 10658 | - | 11 | 47487489 | 47574792 | 4.44E-07 | 7.57E-04 | 8.33E-03 |
| V3_S | MADD | 8567 | + | 11 | 47290927 | 47351582 | 8.62E-07 | 1.24E-03 | 1.62E-02 |
| V3_S | SPTBN1 | 6711 | + | 2 | 54683454 | 54898583 | 6.98E-09 | 3.27E-05 | 1.31E-04 |
| V3_S | HAND1 | 9421 | - | 5 | 153854532 | 153857824 | 4.16E-15 | 4.17E-11 | 7.81E-11 |
| V3_T | NOS1AP | 9722 | + | 1 | 162039581 | 162339813 | 3.52E-18 | 6.61E-14 | 6.61E-14 |
| V3_T | CNOT1 | 23019 | - | 16 | 58553850 | 58663790 | 1.02E-06 | 6.39E-03 | 1.92E-02 |
| V3_T | SLC35F1 | 222553 | + | 6 | 118228689 | 118638839 | 1.76E-06 | 8.23E-03 | 3.29E-02 |
| V3_T | CEP85L | 387119 | - | 6 | 118781935 | 119031238 | 4.77E-08 | 4.47E-04 | 8.94E-04 |
| V3_RS | FNBP4 | 23360 | - | 11 | 47738055 | 47789012 | 1.44E-06 | 2.25E-03 | 2.70E-02 |
| V3_RS | ZNF592 | 9640 | + | 15 | 85291818 | 85349663 | 1.85E-13 | 3.47E-09 | 3.47E-09 |
| V3_RS | WDR73 | 84942 | - | 15 | 85185607 | 85197521 | 5.51E-11 | 2.07E-07 | 1.03E-06 |
| V3_RS | NMB | 4828 | - | 15 | 85198360 | 85201802 | 1.46E-11 | 1.08E-07 | 2.75E-07 |
| V3_RS | SAP30L | 79685 | + | 5 | 153825517 | 153840613 | 2.29E-07 | 6.13E-04 | 4.29E-03 |
| V3_RS | NDUFS3 | 4722 | + | 11 | 47600562 | 47606115 | 1.44E-06 | 2.25E-03 | 2.70E-02 |
| V3_RS | ALPK3 | 57538 | + | 15 | 85359911 | 85416713 | 1.73E-11 | 1.08E-07 | 3.24E-07 |
| V3_RS | SENP2 | 59343 | + | 3 | 185300284 | 185348889 | 2.56E-06 | 3.69E-03 | 4.80E-02 |
| V3_RS | NUP160 | 23279 | - | 11 | 47799670 | 47870057 | 1.27E-06 | 2.25E-03 | 2.38E-02 |
| V3_RS | ZSCAN2 | 54993 | + | 15 | 85144249 | 85166947 | 1.10E-10 | 3.45E-07 | 2.07E-06 |
| V3_RS | MEF2D | 4209 | - | 1 | 156433513 | 156470634 | 2.85E-07 | 6.69E-04 | 5.35E-03 |
| V3_RS | SEC11A | 23478 | - | 15 | 85212768 | 85259691 | 2.31E-11 | 1.08E-07 | 4.33E-07 |
| V3_RS | ADK | 132 | + | 10 | 75910943 | 76469061 | 9.57E-07 | 2.00E-03 | 1.80E-02 |
| V3_ST | SPTBN1 | 6711 | + | 2 | 54683454 | 54898583 | 9.35E-10 | 5.85E-06 | 1.76E-05 |
| V3_ST | NUP160 | 23279 | - | 11 | 47799670 | 47870057 | 1.23E-08 | 5.14E-05 | 2.31E-04 |
| V3_ST | SAP30L | 79685 | + | 5 | 153825517 | 153840613 | 1.01E-13 | 9.44E-10 | 1.89E-09 |
| V3_ST | LPPR2 | 64748 | + | 19 | 11466062 | 11476374 | 2.33E-06 | 3.37E-03 | 4.37E-02 |
| V3_ST | AGBL2 | 79841 | - | 11 | 47681143 | 47736928 | 5.12E-08 | 1.37E-04 | 9.60E-04 |
| V3_ST | BMP8A | 353500 | + | 1 | 39957318 | 39995541 | 1.52E-06 | 2.38E-03 | 2.85E-02 |
| V3_ST | SIPA1L1 | 26037 | + | 14 | 71788108 | 72207761 | 1.84E-08 | 5.76E-05 | 3.46E-04 |
| V3_ST | RAPSN | 5913 | - | 11 | 47459308 | 47470730 | 1.16E-06 | 2.17E-03 | 2.17E-02 |
| V3_ST | CELF1 | 10658 | - | 11 | 47487489 | 47574792 | 1.50E-06 | 2.38E-03 | 2.81E-02 |
| V3_ST | NDUFS3 | 4722 | + | 11 | 47600562 | 47606115 | 6.81E-08 | 1.60E-04 | 1.28E-03 |
| V3_ST | MTCH2 | 23788 | - | 11 | 47638858 | 47664206 | 2.06E-07 | 4.29E-04 | 3.86E-03 |
| V3_ST | HAND1 | 9421 | - | 5 | 153854532 | 153857824 | 3.33E-14 | 6.24E-10 | 6.24E-10 |
| V3_ST | FNBP4 | 23360 | - | 11 | 47738055 | 47789012 | 1.37E-08 | 5.14E-05 | 2.57E-04 |
| V3_R-area | CCDC141 | 285025 | - | 2 | 179694484 | 179914841 | 1.05E-06 | 1.51E-03 | 1.96E-02 |
| V3_R-area | ALPK3 | 57538 | + | 15 | 85359911 | 85416713 | 3.22E-12 | 2.62E-08 | 6.04E-08 |
| V3_R-area | TRIM63 | 84676 | - | 1 | 26377792 | 26394142 | 1.59E-06 | 2.13E-03 | 2.99E-02 |
| V3_R-area | CAMK2D | 817 | - | 4 | 114372188 | 114683669 | 2.00E-07 | 3.76E-04 | 3.75E-03 |
| V3_R-area | NMB | 4828 | - | 15 | 85198360 | 85201802 | 3.00E-12 | 2.62E-08 | 5.64E-08 |
| V3_R-area | ZSCAN2 | 54993 | + | 15 | 85144249 | 85166947 | 1.06E-09 | 3.30E-06 | 1.98E-05 |
| V3_R-area | NFIA | 4774 | + | 1 | 61542946 | 61928460 | 2.00E-07 | 3.76E-04 | 3.76E-03 |
| V3_R-area | WDR73 | 84942 | - | 15 | 85185607 | 85197521 | 1.61E-10 | 6.04E-07 | 3.02E-06 |
| V3_R-area | PDIK1L | 149420 | + | 1 | 26437656 | 26452039 | 1.05E-06 | 1.51E-03 | 1.96E-02 |
| V3_R-area | SPTBN1 | 6711 | + | 2 | 54683454 | 54898583 | 1.45E-07 | 3.41E-04 | 2.73E-03 |
| V3_R-area | ZNF592 | 9640 | + | 15 | 85291818 | 85349663 | 4.20E-12 | 2.62E-08 | 7.87E-08 |
| V3_R-area | ATP5B | 506 | - | 12 | 57031959 | 57039852 | 9.11E-07 | 1.51E-03 | 1.71E-02 |
| V3_R-area | SEC11A | 23478 | - | 15 | 85212768 | 85259691 | 4.22E-11 | 1.98E-07 | 7.92E-07 |
| V3_R-area | HSD17B6 | 8630 | + | 12 | 57146237 | 57181574 | 1.17E-07 | 3.14E-04 | 2.20E-03 |
| V3_S-area | SCN5A | 6331 | - | 3 | 38589553 | 38691164 | 1.89E-09 | 1.18E-05 | 3.54E-05 |
| V3_S-area | NFIA | 4774 | + | 1 | 61542946 | 61928460 | 9.40E-07 | 4.41E-03 | 1.76E-02 |
| V3_S-area | SAP30L | 79685 | + | 5 | 153825517 | 153840613 | 2.33E-11 | 4.37E-07 | 4.37E-07 |
| V3_S-area | HAND1 | 9421 | - | 5 | 153854532 | 153857824 | 1.91E-10 | 1.79E-06 | 3.58E-06 |
| V3_t_S | GALNT8 | 26290 | + | 12 | 4829752 | 4881892 | 2.09E-06 | 7.82E-03 | 3.91E-02 |
| V3_t_S | CEP85L | 387119 | - | 6 | 118781935 | 119031238 | 3.33E-07 | 3.12E-03 | 6.24E-03 |
| V3_t_S | SLC35F1 | 222553 | + | 6 | 118228689 | 118638839 | 9.28E-08 | 1.74E-03 | 1.74E-03 |
| V3_t_S | KCND3 | 3752 | - | 1 | 112318444 | 112532147 | 5.07E-07 | 3.17E-03 | 9.51E-03 |
| V3_t_S | CDKN1A | 1026 | + | 6 | 36644237 | 36655116 | 1.29E-06 | 6.07E-03 | 2.43E-02 |
| V4_R | C11orf16 | 56673 | - | 11 | 8941623 | 8954553 | 1.35E-06 | 2.65E-03 | 2.54E-02 |
| V4_R | AKIP1 | 56672 | + | 11 | 8932701 | 8941626 | 1.41E-06 | 2.65E-03 | 2.65E-02 |
| V4_R | NMB | 4828 | - | 15 | 85198360 | 85201802 | 2.05E-08 | 1.28E-04 | 3.84E-04 |
| V4_R | ZSCAN2 | 54993 | + | 15 | 85144249 | 85166947 | 4.40E-07 | 1.18E-03 | 8.25E-03 |
| V4_R | SEC11A | 23478 | - | 15 | 85212768 | 85259691 | 3.52E-08 | 1.65E-04 | 6.61E-04 |
| V4_R | NRIP3 | 56675 | - | 11 | 9002123 | 9025596 | 1.17E-07 | 4.38E-04 | 2.19E-03 |
| V4_R | ZNF592 | 9640 | + | 15 | 85291818 | 85349663 | 2.67E-09 | 5.02E-05 | 5.02E-05 |
| V4_R | ASCL3 | 56676 | - | 11 | 8959119 | 8964580 | 7.31E-07 | 1.71E-03 | 1.37E-02 |
| V4_R | WDR73 | 84942 | - | 15 | 85185607 | 85197521 | 2.02E-07 | 6.33E-04 | 3.80E-03 |
| V4_R | ALPK3 | 57538 | + | 15 | 85359911 | 85416713 | 7.82E-09 | 7.34E-05 | 1.47E-04 |
| V4_S | DFNB59 | 494513 | + | 2 | 179316163 | 179326149 | 7.02E-08 | 1.01E-04 | 1.32E-03 |
| V4_S | CYP2S1 | 29785 | + | 19 | 41699115 | 41713444 | 1.79E-07 | 2.40E-04 | 3.36E-03 |
| V4_S | SIPA1L1 | 26037 | + | 14 | 71788108 | 72207761 | 5.83E-10 | 1.56E-06 | 1.09E-05 |
| V4_S | SPI1 | 6688 | - | 11 | 47376409 | 47400127 | 1.69E-06 | 1.58E-03 | 3.17E-02 |
| V4_S | PLEKHA3 | 65977 | + | 2 | 179345199 | 179369783 | 1.26E-08 | 2.37E-05 | 2.37E-04 |
| V4_S | SAP30L | 79685 | + | 5 | 153825517 | 153840613 | 6.61E-15 | 1.24E-10 | 1.24E-10 |
| V4_S | FKBP7 | 51661 | - | 2 | 179328391 | 179343646 | 2.29E-06 | 1.95E-03 | 4.29E-02 |
| V4_S | SLC39A13 | 91252 | + | 11 | 47428683 | 47438051 | 3.66E-07 | 4.58E-04 | 6.86E-03 |
| V4_S | MTCH2 | 23788 | - | 11 | 47638858 | 47664206 | 5.98E-08 | 9.35E-05 | 1.12E-03 |
| V4_S | RAPSN | 5913 | - | 11 | 47459308 | 47470730 | 2.13E-09 | 5.00E-06 | 4.00E-05 |
| V4_S | ADK | 132 | + | 10 | 75910943 | 76469061 | 5.39E-08 | 9.20E-05 | 1.01E-03 |
| V4_S | FNBP4 | 23360 | - | 11 | 47738055 | 47789012 | 2.06E-06 | 1.84E-03 | 3.86E-02 |
| V4_S | NR1H3 | 10062 | + | 11 | 47269851 | 47290584 | 8.61E-07 | 9.50E-04 | 1.62E-02 |
| V4_S | CDKN1A | 1026 | + | 6 | 36644237 | 36655116 | 4.91E-10 | 1.54E-06 | 9.21E-06 |
| V4_S | CELF1 | 10658 | - | 11 | 47487489 | 47574792 | 9.38E-07 | 9.62E-04 | 1.76E-02 |
| V4_S | NDUFS3 | 4722 | + | 11 | 47600562 | 47606115 | 4.49E-09 | 9.37E-06 | 8.43E-05 |
| V4_S | SPTBN1 | 6711 | + | 2 | 54683454 | 54898583 | 4.62E-10 | 1.54E-06 | 8.66E-06 |
| V4_S | MADD | 8567 | + | 11 | 47290927 | 47351582 | 6.57E-07 | 7.70E-04 | 1.23E-02 |
| V4_S | MYBPC3 | 4607 | - | 11 | 47352957 | 47374253 | 9.74E-07 | 9.62E-04 | 1.83E-02 |
| V4_S | TTN | 7273 | - | 2 | 179390716 | 179672150 | 3.52E-11 | 2.20E-07 | 6.60E-07 |
| V4_S | SCN5A | 6331 | - | 3 | 38589553 | 38691164 | 2.85E-10 | 1.34E-06 | 5.35E-06 |
| V4_S | HAND1 | 9421 | - | 5 | 153854532 | 153857824 | 2.46E-13 | 2.31E-09 | 4.61E-09 |
| V4_T | NOS1AP | 9722 | + | 1 | 162039581 | 162339813 | 4.21E-14 | 7.89E-10 | 7.89E-10 |
| V4_RS | NUP160 | 23279 | - | 11 | 47799670 | 47870057 | 2.80E-07 | 3.50E-04 | 5.26E-03 |
| V4_RS | ZSCAN2 | 54993 | + | 15 | 85144249 | 85166947 | 4.54E-09 | 1.22E-05 | 8.51E-05 |
| V4_RS | MYBPC3 | 4607 | - | 11 | 47352957 | 47374253 | 2.33E-06 | 1.90E-03 | 4.38E-02 |
| V4_RS | SLC39A13 | 91252 | + | 11 | 47428683 | 47438051 | 2.32E-06 | 1.90E-03 | 4.35E-02 |
| V4_RS | SENP2 | 59343 | + | 3 | 185300284 | 185348889 | 1.31E-06 | 1.23E-03 | 2.47E-02 |
| V4_RS | NR1H3 | 10062 | + | 11 | 47269851 | 47290584 | 5.98E-07 | 6.23E-04 | 1.12E-02 |
| V4_RS | CRIM1 | 51232 | + | 2 | 36583370 | 36778278 | 2.22E-08 | 4.46E-05 | 4.16E-04 |
| V4_RS | ARFGAP2 | 84364 | - | 11 | 47185849 | 47199054 | 1.82E-06 | 1.62E-03 | 3.41E-02 |
| V4_RS | WDR73 | 84942 | - | 15 | 85185607 | 85197521 | 4.40E-09 | 1.22E-05 | 8.25E-05 |
| V4_RS | RAPSN | 5913 | - | 11 | 47459308 | 47470730 | 1.67E-07 | 2.41E-04 | 3.13E-03 |
| V4_RS | CELF1 | 10658 | - | 11 | 47487489 | 47574792 | 8.16E-07 | 8.06E-04 | 1.53E-02 |
| V4_RS | ZNF592 | 9640 | + | 15 | 85291818 | 85349663 | 8.44E-12 | 1.58E-07 | 1.58E-07 |
| V4_RS | KCND3 | 3752 | - | 1 | 112318444 | 112532147 | 6.68E-10 | 2.50E-06 | 1.25E-05 |
| V4_RS | SIPA1L1 | 26037 | + | 14 | 71788108 | 72207761 | 7.22E-08 | 1.23E-04 | 1.36E-03 |
| V4_RS | NMB | 4828 | - | 15 | 85198360 | 85201802 | 4.26E-10 | 2.00E-06 | 7.99E-06 |
| V4_RS | MADD | 8567 | + | 11 | 47290927 | 47351582 | 4.81E-07 | 5.31E-04 | 9.03E-03 |
| V4_RS | ALPK3 | 57538 | + | 15 | 85359911 | 85416713 | 3.43E-10 | 2.00E-06 | 6.44E-06 |
| V4_RS | FNBP4 | 23360 | - | 11 | 47738055 | 47789012 | 2.00E-07 | 2.68E-04 | 3.76E-03 |
| V4_RS | NDUFS3 | 4722 | + | 11 | 47600562 | 47606115 | 2.13E-08 | 4.46E-05 | 3.99E-04 |
| V4_RS | MTCH2 | 23788 | - | 11 | 47638858 | 47664206 | 1.27E-07 | 1.98E-04 | 2.38E-03 |
| V4_RS | AGBL2 | 79841 | - | 11 | 47681143 | 47736928 | 4.02E-07 | 4.71E-04 | 7.54E-03 |
| V4_RS | SEC11A | 23478 | - | 15 | 85212768 | 85259691 | 2.94E-10 | 2.00E-06 | 5.51E-06 |
| V4_RS | ADK | 132 | + | 10 | 75910943 | 76469061 | 2.37E-08 | 4.46E-05 | 4.46E-04 |
| V4_ST | RAPSN | 5913 | - | 11 | 47459308 | 47470730 | 3.58E-09 | 1.12E-05 | 6.72E-05 |
| V4_ST | CELF1 | 10658 | - | 11 | 47487489 | 47574792 | 1.41E-06 | 1.55E-03 | 2.64E-02 |
| V4_ST | MTCH2 | 23788 | - | 11 | 47638858 | 47664206 | 2.48E-07 | 4.66E-04 | 4.66E-03 |
| V4_ST | NR1H3 | 10062 | + | 11 | 47269851 | 47290584 | 1.86E-06 | 1.74E-03 | 3.48E-02 |
| V4_ST | SPTBN1 | 6711 | + | 2 | 54683454 | 54898583 | 2.93E-11 | 1.37E-07 | 5.50E-07 |
| V4_ST | MADD | 8567 | + | 11 | 47290927 | 47351582 | 2.04E-06 | 1.80E-03 | 3.83E-02 |
| V4_ST | SLC39A13 | 91252 | + | 11 | 47428683 | 47438051 | 5.63E-07 | 9.60E-04 | 1.06E-02 |
| V4_ST | FNBP4 | 23360 | - | 11 | 47738055 | 47789012 | 9.00E-07 | 1.21E-03 | 1.69E-02 |
| V4_ST | HAND1 | 9421 | - | 5 | 153854532 | 153857824 | 9.68E-14 | 1.82E-09 | 1.82E-09 |
| V4_ST | ACTL7A | 10881 | + | 9 | 111624603 | 111626035 | 2.11E-06 | 1.80E-03 | 3.95E-02 |
| V4_ST | BMP8A | 353500 | + | 1 | 39957318 | 39995541 | 1.37E-06 | 1.55E-03 | 2.58E-02 |
| V4_ST | SAP30L | 79685 | + | 5 | 153825517 | 153840613 | 4.90E-13 | 4.59E-09 | 9.18E-09 |
| V4_ST | KCND3 | 3752 | - | 1 | 112318444 | 112532147 | 1.50E-06 | 1.56E-03 | 2.82E-02 |
| V4_ST | PLEKHA3 | 65977 | + | 2 | 179345199 | 179369783 | 1.02E-08 | 2.74E-05 | 1.92E-04 |
| V4_ST | SIPA1L1 | 26037 | + | 14 | 71788108 | 72207761 | 2.83E-10 | 1.06E-06 | 5.32E-06 |
| V4_ST | TTN | 7273 | - | 2 | 179390716 | 179672150 | 1.70E-11 | 1.06E-07 | 3.19E-07 |
| V4_ST | CDKN1A | 1026 | + | 6 | 36644237 | 36655116 | 8.70E-07 | 1.21E-03 | 1.63E-02 |
| V4_ST | NDUFS3 | 4722 | + | 11 | 47600562 | 47606115 | 3.70E-08 | 8.68E-05 | 6.94E-04 |
| V4_ST | MYBPC3 | 4607 | - | 11 | 47352957 | 47374253 | 1.05E-06 | 1.32E-03 | 1.97E-02 |
| V4_ST | DFNB59 | 494513 | + | 2 | 179316163 | 179326149 | 5.13E-08 | 1.07E-04 | 9.63E-04 |
| V4_ST | NUP160 | 23279 | - | 11 | 47799670 | 47870057 | 1.64E-06 | 1.62E-03 | 3.08E-02 |
| V4_ST | CYP2S1 | 29785 | + | 19 | 41699115 | 41713444 | 6.65E-07 | 1.04E-03 | 1.25E-02 |
| V4_J_up | PDZRN4 | 29951 | + | 12 | 41582250 | 41968392 | 5.45E-08 | 1.02E-03 | 1.02E-03 |
| V4_R-area | FKBP7 | 51661 | - | 2 | 179328391 | 179343646 | 2.16E-07 | 6.74E-04 | 4.05E-03 |
| V4_R-area | NRIP3 | 56675 | - | 11 | 9002123 | 9025596 | 5.29E-09 | 9.93E-05 | 9.93E-05 |
| V4_R-area | ASCL3 | 56676 | - | 11 | 8959119 | 8964580 | 5.03E-08 | 3.98E-04 | 9.45E-04 |
| V4_R-area | SCUBE2 | 57758 | - | 11 | 9041047 | 9113150 | 7.54E-07 | 1.77E-03 | 1.41E-02 |
| V4_R-area | C11orf16 | 56673 | - | 11 | 8941623 | 8954553 | 1.06E-07 | 3.98E-04 | 1.99E-03 |
| V4_R-area | OSBPL6 | 114880 | + | 2 | 179059208 | 179264160 | 1.83E-06 | 3.44E-03 | 3.44E-02 |
| V4_R-area | CRIM1 | 51232 | + | 2 | 36583370 | 36778278 | 1.28E-06 | 2.66E-03 | 2.40E-02 |
| V4_R-area | ST5 | 6764 | - | 11 | 8714899 | 8932498 | 8.51E-08 | 3.98E-04 | 1.60E-03 |
| V4_R-area | TMEM9B | 56674 | - | 11 | 8968840 | 8986553 | 3.36E-07 | 9.02E-04 | 6.31E-03 |
| V4_R-area | AKIP1 | 56672 | + | 11 | 8932701 | 8941626 | 9.41E-08 | 3.98E-04 | 1.77E-03 |
| V4_S-area | CDKN1A | 1026 | + | 6 | 36644237 | 36655116 | 4.26E-09 | 2.66E-05 | 7.99E-05 |
| V4_S-area | HAND1 | 9421 | - | 5 | 153854532 | 153857824 | 7.82E-08 | 2.45E-04 | 1.47E-03 |
| V4_S-area | ADK | 132 | + | 10 | 75910943 | 76469061 | 3.90E-08 | 1.83E-04 | 7.31E-04 |
| V4_S-area | SCN5A | 6331 | - | 3 | 38589553 | 38691164 | 8.49E-12 | 1.59E-07 | 1.59E-07 |
| V4_S-area | TTN | 7273 | - | 2 | 179390716 | 179672150 | 1.09E-07 | 2.91E-04 | 2.04E-03 |
| V4_S-area | NFIA | 4774 | + | 1 | 61542946 | 61928460 | 6.81E-08 | 2.45E-04 | 1.28E-03 |
| V4_S-area | SAP30L | 79685 | + | 5 | 153825517 | 153840613 | 1.86E-09 | 1.75E-05 | 3.50E-05 |
| V4_t_S | BACH1 | 571 | + | 21 | 30671151 | 30734257 | 1.49E-06 | 2.79E-03 | 2.79E-02 |
| V4_t_S | PITX3 | 5309 | - | 10 | 103989946 | 104001231 | 7.99E-07 | 2.14E-03 | 1.50E-02 |
| V4_t_S | GBF1 | 8729 | + | 10 | 104005255 | 104142656 | 2.62E-07 | 1.11E-03 | 4.91E-03 |
| V4_t_S | USP16 | 10600 | + | 21 | 30396938 | 30426809 | 7.96E-07 | 2.14E-03 | 1.49E-02 |
| V4_t_S | TMEM44 | 93109 | - | 3 | 194308402 | 194354150 | 1.30E-06 | 2.71E-03 | 2.44E-02 |
| V4_t_S | KCND3 | 3752 | - | 1 | 112318444 | 112532147 | 1.39E-12 | 2.61E-08 | 2.61E-08 |
| V4_t_S | MAP3K7CL | 56911 | + | 21 | 30449659 | 30548210 | 1.21E-07 | 7.59E-04 | 2.28E-03 |
| V4_t_S | FBXL15 | 79176 | + | 10 | 104178979 | 104182894 | 1.09E-06 | 2.56E-03 | 2.05E-02 |
| V4_t_S | DPT | 1805 | - | 1 | 168664695 | 168698442 | 9.89E-09 | 9.28E-05 | 1.86E-04 |
| V4_t_S | GALNT8 | 26290 | + | 12 | 4829752 | 4881892 | 2.95E-07 | 1.11E-03 | 5.54E-03 |
| V4_t_S | NFKB2 | 4791 | + | 10 | 104153867 | 104162286 | 2.39E-06 | 4.08E-03 | 4.48E-02 |
| V4_t_ST | CCR4 | 1233 | + | 3 | 32993066 | 32996403 | 1.04E-06 | 1.96E-02 | 1.96E-02 |
| V5_R | ADK | 132 | + | 10 | 75910943 | 76469061 | 1.34E-06 | 5.05E-03 | 2.52E-02 |
| V5_R | NDUFS3 | 4722 | + | 11 | 47600562 | 47606115 | 4.64E-07 | 4.00E-03 | 8.71E-03 |
| V5_R | NCOA2 | 10499 | - | 8 | 71021997 | 71316062 | 2.24E-06 | 7.01E-03 | 4.21E-02 |
| V5_R | MTCH2 | 23788 | - | 11 | 47638858 | 47664206 | 6.40E-07 | 4.00E-03 | 1.20E-02 |
| V5_R | CRIM1 | 51232 | + | 2 | 36583370 | 36778278 | 1.06E-06 | 4.99E-03 | 2.00E-02 |
| V5_R | KCND3 | 3752 | - | 1 | 112318444 | 112532147 | 3.84E-10 | 7.21E-06 | 7.21E-06 |
| V5_S | TTN | 7273 | - | 2 | 179390716 | 179672150 | 2.89E-09 | 1.81E-05 | 5.43E-05 |
| V5_S | SCN5A | 6331 | - | 3 | 38589553 | 38691164 | 7.20E-10 | 6.76E-06 | 1.35E-05 |
| V5_S | SIPA1L1 | 26037 | + | 14 | 71788108 | 72207761 | 8.84E-08 | 3.32E-04 | 1.66E-03 |
| V5_S | AFAP1 | 60312 | - | 4 | 7760440 | 7941653 | 4.56E-07 | 1.43E-03 | 8.55E-03 |
| V5_S | RAPSN | 5913 | - | 11 | 47459308 | 47470730 | 1.40E-06 | 2.92E-03 | 2.63E-02 |
| V5_S | ADK | 132 | + | 10 | 75910943 | 76469061 | 6.68E-07 | 1.79E-03 | 1.25E-02 |
| V5_S | NDUFS3 | 4722 | + | 11 | 47600562 | 47606115 | 2.28E-06 | 4.28E-03 | 4.28E-02 |
| V5_S | PLEKHA3 | 65977 | + | 2 | 179345199 | 179369783 | 1.24E-06 | 2.92E-03 | 2.33E-02 |
| V5_S | CDKN1A | 1026 | + | 6 | 36644237 | 36655116 | 2.48E-11 | 4.66E-07 | 4.66E-07 |
| V5_S | KCND3 | 3752 | - | 1 | 112318444 | 112532147 | 2.82E-08 | 1.32E-04 | 5.29E-04 |
| V5_T | HSPB7 | 27129 | - | 1 | 16340523 | 16346084 | 2.04E-06 | 7.65E-03 | 3.82E-02 |
| V5_T | ZBTB17 | 7709 | - | 1 | 16268364 | 16302627 | 1.87E-06 | 7.65E-03 | 3.50E-02 |
| V5_T | PDZK1IP1 | 10158 | - | 1 | 47649261 | 47655771 | 9.90E-07 | 7.65E-03 | 1.86E-02 |
| V5_T | NOS1AP | 9722 | + | 1 | 162039581 | 162339813 | 4.68E-07 | 7.65E-03 | 8.79E-03 |
| V5_T | GLYR1 | 84656 | - | 16 | 4853204 | 4897383 | 1.79E-06 | 7.65E-03 | 3.37E-02 |
| V5_T | PLCL1 | 5334 | + | 2 | 198669426 | 199014608 | 2.65E-06 | 7.83E-03 | 4.97E-02 |
| V5_RS | C1QTNF4 | 114900 | - | 11 | 47611216 | 47615961 | 3.03E-07 | 5.69E-04 | 5.69E-03 |
| V5_RS | SPI1 | 6688 | - | 11 | 47376409 | 47400127 | 1.53E-07 | 3.18E-04 | 2.86E-03 |
| V5_RS | FNBP4 | 23360 | - | 11 | 47738055 | 47789012 | 1.88E-06 | 2.71E-03 | 3.53E-02 |
| V5_RS | ADK | 132 | + | 10 | 75910943 | 76469061 | 3.33E-09 | 1.93E-05 | 6.26E-05 |
| V5_RS | CELF1 | 10658 | - | 11 | 47487489 | 47574792 | 9.95E-07 | 1.56E-03 | 1.87E-02 |
| V5_RS | MTCH2 | 23788 | - | 11 | 47638858 | 47664206 | 4.12E-09 | 1.93E-05 | 7.74E-05 |
| V5_RS | PSMC3 | 5702 | - | 11 | 47440320 | 47448024 | 3.77E-08 | 1.18E-04 | 7.07E-04 |
| V5_RS | RAPSN | 5913 | - | 11 | 47459308 | 47470730 | 1.61E-08 | 6.03E-05 | 3.01E-04 |
| V5_RS | CRIM1 | 51232 | + | 2 | 36583370 | 36778278 | 7.87E-08 | 1.85E-04 | 1.48E-03 |
| V5_RS | KCND3 | 3752 | - | 1 | 112318444 | 112532147 | 4.30E-13 | 8.07E-09 | 8.07E-09 |
| V5_RS | SLC39A13 | 91252 | + | 11 | 47428683 | 47438051 | 7.77E-08 | 1.85E-04 | 1.46E-03 |
| V5_RS | NCOA2 | 10499 | - | 8 | 71021997 | 71316062 | 4.79E-07 | 8.17E-04 | 8.99E-03 |
| V5_RS | NDUFS3 | 4722 | + | 11 | 47600562 | 47606115 | 1.85E-09 | 1.74E-05 | 3.47E-05 |
| V5_ST | SIPA1L1 | 26037 | + | 14 | 71788108 | 72207761 | 3.31E-07 | 2.07E-03 | 6.20E-03 |
| V5_ST | TTN | 7273 | - | 2 | 179390716 | 179672150 | 5.24E-09 | 9.84E-05 | 9.84E-05 |
| V5_ST | PLEKHA3 | 65977 | + | 2 | 179345199 | 179369783 | 7.16E-07 | 3.36E-03 | 1.34E-02 |
| V5_ST | CDKN1A | 1026 | + | 6 | 36644237 | 36655116 | 9.28E-07 | 3.48E-03 | 1.74E-02 |
| V5_ST | RAPSN | 5913 | - | 11 | 47459308 | 47470730 | 2.44E-07 | 2.07E-03 | 4.57E-03 |
| V5_ST | SPTBN1 | 6711 | + | 2 | 54683454 | 54898583 | 1.64E-06 | 5.14E-03 | 3.09E-02 |
| V5_J_up | NSRP1 | 84081 | + | 17 | 28443821 | 28513493 | 8.54E-07 | 1.78E-03 | 1.60E-02 |
| V5_J_up | PLEKHA3 | 65977 | + | 2 | 179345199 | 179369783 | 2.78E-09 | 2.61E-05 | 5.22E-05 |
| V5_J_up | TTN | 7273 | - | 2 | 179390716 | 179672150 | 6.12E-16 | 1.15E-11 | 1.15E-11 |
| V5_J_up | SCN5A | 6331 | - | 3 | 38589553 | 38691164 | 7.36E-07 | 1.73E-03 | 1.38E-02 |
| V5_J_up | PRKRA | 8575 | - | 2 | 179296141 | 179315958 | 1.36E-06 | 2.32E-03 | 2.55E-02 |
| V5_J_up | PTPRJ | 5795 | + | 11 | 48002101 | 48192394 | 5.50E-08 | 2.06E-04 | 1.03E-03 |
| V5_J_up | DFNB59 | 494513 | + | 2 | 179316163 | 179326149 | 1.11E-08 | 6.91E-05 | 2.07E-04 |
| V5_J_up | FKBP7 | 51661 | - | 2 | 179328391 | 179343646 | 1.31E-07 | 4.08E-04 | 2.45E-03 |
| V5_J_up | KCND3 | 3752 | - | 1 | 112318444 | 112532147 | 4.41E-07 | 1.18E-03 | 8.28E-03 |
| V5_J_up | LOC388780 | 388780 | + | 20 | 2187574 | 2193797 | 1.04E-06 | 1.95E-03 | 1.95E-02 |
| V5_J_up | CDKN1A | 1026 | + | 6 | 36644237 | 36655116 | 2.16E-08 | 1.01E-04 | 4.05E-04 |
| V5_R-area | MTCH2 | 23788 | - | 11 | 47638858 | 47664206 | 6.66E-07 | 3.12E-03 | 1.25E-02 |
| V5_R-area | KCND3 | 3752 | - | 1 | 112318444 | 112532147 | 1.26E-09 | 2.36E-05 | 2.36E-05 |
| V5_R-area | NDUFS3 | 4722 | + | 11 | 47600562 | 47606115 | 2.00E-06 | 7.52E-03 | 3.76E-02 |
| V5_R-area | CRIM1 | 51232 | + | 2 | 36583370 | 36778278 | 8.44E-09 | 7.92E-05 | 1.58E-04 |
| V5_R-area | NCOA2 | 10499 | - | 8 | 71021997 | 71316062 | 4.91E-07 | 3.07E-03 | 9.21E-03 |
| V5_S-area | KCND3 | 3752 | - | 1 | 112318444 | 112532147 | 3.48E-11 | 6.52E-07 | 6.52E-07 |
| V5_S-area | AK5 | 26289 | + | 1 | 77747662 | 78025654 | 2.27E-06 | 4.20E-03 | 4.26E-02 |
| V5_S-area | ACTL7B | 10880 | - | 9 | 111616868 | 111618275 | 3.01E-07 | 1.41E-03 | 5.65E-03 |
| V5_S-area | NDUFS3 | 4722 | + | 11 | 47600562 | 47606115 | 1.34E-06 | 3.60E-03 | 2.52E-02 |
| V5_S-area | CDKN1A | 1026 | + | 6 | 36644237 | 36655116 | 8.79E-08 | 5.50E-04 | 1.65E-03 |
| V5_S-area | SCN5A | 6331 | - | 3 | 38589553 | 38691164 | 1.28E-10 | 1.20E-06 | 2.40E-06 |
| V5_S-area | PSMC3 | 5702 | - | 11 | 47440320 | 47448024 | 1.70E-06 | 3.98E-03 | 3.18E-02 |
| V5_S-area | ADK | 132 | + | 10 | 75910943 | 76469061 | 5.46E-07 | 1.71E-03 | 1.03E-02 |
| V5_S-area | RAPSN | 5913 | - | 11 | 47459308 | 47470730 | 2.12E-06 | 4.20E-03 | 3.97E-02 |
| V5_S-area | TTN | 7273 | - | 2 | 179390716 | 179672150 | 4.39E-07 | 1.65E-03 | 8.24E-03 |
| V5_S-area | MTCH2 | 23788 | - | 11 | 47638858 | 47664206 | 2.46E-06 | 4.20E-03 | 4.62E-02 |
| I_S | ZSCAN2 | 54993 | + | 15 | 85144249 | 85166947 | 6.85E-07 | 1.29E-02 | 1.29E-02 |
| II_R | NRP1 | 8829 | - | 10 | 33466419 | 33623833 | 3.53E-07 | 3.31E-03 | 6.62E-03 |
| II_R | ADK | 132 | + | 10 | 75910943 | 76469061 | 7.81E-08 | 1.46E-03 | 1.46E-03 |
| V5_T-area | SPANXN1 | 494118 | + | X | 144329107 | 144337728 | 1.98E-06 | 2.99E-02 | 3.71E-02 |
| V5_t_S | KCND3 | 3752 | - | 1 | 112318444 | 112532147 | 2.08E-10 | 3.90E-06 | 3.90E-06 |
| V5_t_S | TTN | 7273 | - | 2 | 179390716 | 179672150 | 1.93E-06 | 1.81E-02 | 3.63E-02 |
| V5_t_R | CEP85L | 387119 | - | 6 | 118781935 | 119031238 | 2.32E-06 | 2.18E-02 | 4.35E-02 |
| V5_t_R | TTN | 7273 | - | 2 | 179390716 | 179672150 | 2.52E-07 | 4.73E-03 | 4.73E-03 |
| II_S | DUS4L | 11062 | + | 7 | 107204402 | 107218968 | 9.92E-07 | 1.76E-02 | 1.86E-02 |
| II_S | HEATR5B | 54497 | - | 2 | 37208153 | 37311485 | 1.88E-06 | 1.76E-02 | 3.52E-02 |
| V6_R | KCND3 | 3752 | - | 1 | 112318444 | 112532147 | 5.11E-13 | 9.58E-09 | 9.58E-09 |
| V6_R | TTN | 7273 | - | 2 | 179390716 | 179672150 | 2.15E-08 | 2.02E-04 | 4.04E-04 |
| V6_R | CRIM1 | 51232 | + | 2 | 36583370 | 36778278 | 1.07E-06 | 4.01E-03 | 2.00E-02 |
| V6_R | ADK | 132 | + | 10 | 75910943 | 76469061 | 4.83E-07 | 2.26E-03 | 9.06E-03 |
| V6_R | NRP1 | 8829 | - | 10 | 33466419 | 33623833 | 4.19E-08 | 2.62E-04 | 7.86E-04 |
| V6_S | KCND3 | 3752 | - | 1 | 112318444 | 112532147 | 1.28E-06 | 1.73E-02 | 2.40E-02 |
| V6_T | SOX5 | 6660 | - | 12 | 23682438 | 24715383 | 3.27E-08 | 6.14E-04 | 6.14E-04 |
| V6_T | SSBP3 | 23648 | - | 1 | 54691104 | 54872068 | 2.21E-06 | 8.31E-03 | 4.15E-02 |
| V6_T | LMF1 | 64788 | - | 16 | 903634 | 1031318 | 1.76E-06 | 8.26E-03 | 3.30E-02 |
| V6_T | DPT | 1805 | - | 1 | 168664695 | 168698442 | 7.66E-07 | 4.79E-03 | 1.44E-02 |
| V6_T | SREBF1 | 6720 | - | 17 | 17714663 | 17740331 | 7.58E-07 | 4.79E-03 | 1.42E-02 |
| V6_RS | KCND3 | 3752 | - | 1 | 112318444 | 112532147 | 6.71E-14 | 1.26E-09 | 1.26E-09 |
| V6_RS | CRIM1 | 51232 | + | 2 | 36583370 | 36778278 | 1.03E-06 | 3.86E-03 | 1.93E-02 |
| V6_RS | ADK | 132 | + | 10 | 75910943 | 76469061 | 5.31E-08 | 2.49E-04 | 9.96E-04 |
| V6_RS | TTN | 7273 | - | 2 | 179390716 | 179672150 | 1.20E-08 | 7.51E-05 | 2.25E-04 |
| V6_RS | NRP1 | 8829 | - | 10 | 33466419 | 33623833 | 2.49E-09 | 2.34E-05 | 4.67E-05 |
| V6_J_up | TBX3 | 6926 | - | 12 | 115108059 | 115121969 | 1.50E-08 | 2.56E-05 | 2.82E-04 |
| V6_J_up | RBM20 | 282996 | + | 10 | 112404155 | 112599227 | 5.19E-08 | 8.12E-05 | 9.74E-04 |
| V6_J_up | KCND3 | 3752 | - | 1 | 112318444 | 112532147 | 7.01E-10 | 1.46E-06 | 1.31E-05 |
| V6_J_up | PLEKHA3 | 65977 | + | 2 | 179345199 | 179369783 | 7.97E-14 | 4.98E-10 | 1.49E-09 |
| V6_J_up | FKBP7 | 51661 | - | 2 | 179328391 | 179343646 | 3.40E-14 | 3.19E-10 | 6.37E-10 |
| V6_J_up | NRP1 | 8829 | - | 10 | 33466419 | 33623833 | 4.49E-09 | 8.42E-06 | 8.42E-05 |
| V6_J_up | SIPA1L1 | 26037 | + | 14 | 71788108 | 72207761 | 7.13E-07 | 8.92E-04 | 1.34E-02 |
| V6_J_up | PRKRA | 8575 | - | 2 | 179296141 | 179315958 | 8.43E-13 | 2.64E-09 | 1.58E-08 |
| V6_J_up | CCDC141 | 285025 | - | 2 | 179694484 | 179914841 | 2.77E-11 | 6.49E-08 | 5.19E-07 |
| V6_J_up | FOLH1 | 2346 | - | 11 | 49168187 | 49230222 | 3.79E-07 | 5.47E-04 | 7.11E-03 |
| V6_J_up | PTPRJ | 5795 | + | 11 | 48002101 | 48192394 | 5.59E-07 | 7.49E-04 | 1.05E-02 |
| V6_J_up | TTN | 7273 | - | 2 | 179390716 | 179672150 | 4.74E-15 | 8.89E-11 | 8.89E-11 |
| V6_J_up | CDKN1A | 1026 | + | 6 | 36644237 | 36655116 | 3.31E-13 | 1.24E-09 | 6.20E-09 |
| V6_J_up | OSBPL6 | 114880 | + | 2 | 179059208 | 179264160 | 2.33E-06 | 2.73E-03 | 4.37E-02 |
| V6_J_up | SCN5A | 6331 | - | 3 | 38589553 | 38691164 | 1.45E-11 | 3.88E-08 | 2.72E-07 |
| V6_J_up | DFNB59 | 494513 | + | 2 | 179316163 | 179326149 | 3.13E-13 | 1.24E-09 | 5.88E-09 |
| V6_R-area | CRIM1 | 51232 | + | 2 | 36583370 | 36778278 | 3.04E-08 | 2.85E-04 | 5.70E-04 |
| V6_R-area | KCND3 | 3752 | - | 1 | 112318444 | 112532147 | 2.01E-12 | 3.78E-08 | 3.78E-08 |
| V6_R-area | TTN | 7273 | - | 2 | 179390716 | 179672150 | 2.27E-07 | 1.42E-03 | 4.26E-03 |
| II_T | LMF1 | 64788 | - | 16 | 903634 | 1031318 | 6.29E-07 | 1.18E-02 | 1.18E-02 |
| V6_t_S | FKBP7 | 51661 | - | 2 | 179328391 | 179343646 | 1.91E-06 | 7.15E-03 | 3.58E-02 |
| V6_t_S | NRP1 | 8829 | - | 10 | 33466419 | 33623833 | 4.07E-07 | 2.55E-03 | 7.65E-03 |
| V6_t_S | DFNB59 | 494513 | + | 2 | 179316163 | 179326149 | 7.82E-07 | 3.67E-03 | 1.47E-02 |
| V6_t_S | TTN | 7273 | - | 2 | 179390716 | 179672150 | 1.54E-10 | 2.89E-06 | 2.89E-06 |
| V6_t_S | PLEKHA3 | 65977 | + | 2 | 179345199 | 179369783 | 3.98E-07 | 2.55E-03 | 7.46E-03 |
| V6_t_R | TTN | 7273 | - | 2 | 179390716 | 179672150 | 4.74E-10 | 8.89E-06 | 8.89E-06 |
| V6_t_R | KCND3 | 3752 | - | 1 | 112318444 | 112532147 | 2.61E-09 | 2.45E-05 | 4.90E-05 |
| V6_t_R | PLEKHA3 | 65977 | + | 2 | 179345199 | 179369783 | 2.36E-06 | 1.48E-02 | 4.44E-02 |
| V6_t_ST | DFNB59 | 494513 | + | 2 | 179316163 | 179326149 | 1.59E-06 | 9.93E-03 | 2.98E-02 |
| V6_t_ST | TTN | 7273 | - | 2 | 179390716 | 179672150 | 6.65E-10 | 1.25E-05 | 1.25E-05 |
| V6_t_ST | NRP1 | 8829 | - | 10 | 33466419 | 33623833 | 1.09E-08 | 1.02E-04 | 2.04E-04 |
| V6_t_QT | NRP1 | 8829 | - | 10 | 33466419 | 33623833 | 2.80E-07 | 5.25E-03 | 5.25E-03 |
| II_RS | CRIM1 | 51232 | + | 2 | 36583370 | 36778278 | 2.09E-06 | 1.31E-02 | 3.92E-02 |
| II_RS | ADK | 132 | + | 10 | 75910943 | 76469061 | 9.80E-09 | 9.20E-05 | 1.84E-04 |
| II_RS | NRP1 | 8829 | - | 10 | 33466419 | 33623833 | 6.11E-09 | 9.20E-05 | 1.15E-04 |
| II_ST | HEATR5B | 54497 | - | 2 | 37208153 | 37311485 | 2.05E-07 | 2.74E-03 | 3.85E-03 |
| II_ST | LMF1 | 64788 | - | 16 | 903634 | 1031318 | 2.24E-06 | 1.05E-02 | 4.21E-02 |
| II_ST | DUS4L | 11062 | + | 7 | 107204402 | 107218968 | 5.73E-07 | 3.58E-03 | 1.08E-02 |
| II_ST | HSPB7 | 27129 | - | 1 | 16340523 | 16346084 | 2.92E-07 | 2.74E-03 | 5.49E-03 |
| II_J_up | TBX3 | 6926 | - | 12 | 115108059 | 115121969 | 2.82E-08 | 1.32E-04 | 5.29E-04 |
| II_J_up | FKBP7 | 51661 | - | 2 | 179328391 | 179343646 | 1.09E-06 | 2.91E-03 | 2.04E-02 |
| II_J_up | CCDC141 | 285025 | - | 2 | 179694484 | 179914841 | 2.51E-10 | 1.57E-06 | 4.71E-06 |
| II_J_up | CERS3 | 204219 | - | 15 | 100940600 | 101084925 | 1.50E-07 | 5.61E-04 | 2.81E-03 |
| II_J_up | PRDM6 | 93166 | + | 5 | 122424841 | 122529960 | 5.05E-07 | 1.58E-03 | 9.47E-03 |
| II_J_up | TTN | 7273 | - | 2 | 179390716 | 179672150 | 1.46E-14 | 2.74E-10 | 2.74E-10 |
| II_J_up | NRP1 | 8829 | - | 10 | 33466419 | 33623833 | 1.66E-11 | 1.56E-07 | 3.12E-07 |
| II_R-area | ADK | 132 | + | 10 | 75910943 | 76469061 | 1.62E-07 | 3.05E-03 | 3.05E-03 |
| I_T | DEFB136 | 613210 | - | 8 | 11831446 | 11832108 | 2.19E-08 | 1.39E-04 | 4.12E-04 |
| I_T | BLK | 640 | + | 8 | 11351521 | 11422108 | 4.23E-07 | 1.59E-03 | 7.93E-03 |
| I_T | DEFB134 | 613211 | - | 8 | 11851489 | 11853760 | 2.23E-08 | 1.39E-04 | 4.18E-04 |
| I_T | SOX5 | 6660 | - | 12 | 23682438 | 24715383 | 3.25E-09 | 6.10E-05 | 6.10E-05 |
| I_T | LMF1 | 64788 | - | 16 | 903634 | 1031318 | 2.44E-07 | 1.15E-03 | 4.58E-03 |
| II_t_S | NRP1 | 8829 | - | 10 | 33466419 | 33623833 | 9.13E-07 | 1.71E-02 | 1.71E-02 |
| II_t_ST | NRP1 | 8829 | - | 10 | 33466419 | 33623833 | 1.53E-08 | 2.86E-04 | 2.86E-04 |
| II_t_QT | NRP1 | 8829 | - | 10 | 33466419 | 33623833 | 9.80E-08 | 1.84E-03 | 1.84E-03 |
| III_R | ADK | 132 | + | 10 | 75910943 | 76469061 | 1.18E-06 | 1.05E-02 | 2.22E-02 |
| III_R | ZNF592 | 9640 | + | 15 | 85291818 | 85349663 | 1.68E-06 | 1.05E-02 | 3.15E-02 |
| III_R | CCDC141 | 285025 | - | 2 | 179694484 | 179914841 | 1.66E-07 | 3.11E-03 | 3.11E-03 |
| III_S | NRP1 | 8829 | - | 10 | 33466419 | 33623833 | 1.42E-12 | 2.67E-08 | 2.67E-08 |
| III_S | CCDC141 | 285025 | - | 2 | 179694484 | 179914841 | 6.50E-08 | 6.09E-04 | 1.22E-03 |
| III_S | VTI1A | 143187 | + | 10 | 114206756 | 114578503 | 1.95E-06 | 7.32E-03 | 3.66E-02 |
| III_S | SIPA1L1 | 26037 | + | 14 | 71788108 | 72207761 | 8.34E-07 | 5.22E-03 | 1.57E-02 |
| III_S | CASQ2 | 845 | - | 1 | 116242624 | 116311426 | 1.38E-06 | 6.47E-03 | 2.59E-02 |
| III_T | CASP7 | 840 | + | 10 | 115438921 | 115490668 | 2.23E-06 | 2.39E-02 | 4.19E-02 |
| III_T | MYLK | 4638 | - | 3 | 123331143 | 123603149 | 2.55E-06 | 2.39E-02 | 4.78E-02 |
| III_RS | CWC27 | 10283 | + | 5 | 64064755 | 64314590 | 7.68E-07 | 4.55E-03 | 1.44E-02 |
| III_RS | CCDC141 | 285025 | - | 2 | 179694484 | 179914841 | 2.55E-10 | 2.39E-06 | 4.78E-06 |
| III_RS | NRP1 | 8829 | - | 10 | 33466419 | 33623833 | 4.48E-13 | 8.40E-09 | 8.40E-09 |
| III_RS | ADK | 132 | + | 10 | 75910943 | 76469061 | 9.71E-07 | 4.55E-03 | 1.82E-02 |
| III_ST | CASQ2 | 845 | - | 1 | 116242624 | 116311426 | 4.93E-07 | 1.85E-03 | 9.25E-03 |
| III_ST | NRP1 | 8829 | - | 10 | 33466419 | 33623833 | 2.54E-11 | 4.76E-07 | 4.76E-07 |
| III_ST | VTI1A | 143187 | + | 10 | 114206756 | 114578503 | 5.74E-08 | 3.80E-04 | 1.08E-03 |
| III_ST | CCDC141 | 285025 | - | 2 | 179694484 | 179914841 | 6.07E-08 | 3.80E-04 | 1.14E-03 |
| III_ST | SIPA1L1 | 26037 | + | 14 | 71788108 | 72207761 | 1.50E-07 | 7.04E-04 | 2.81E-03 |
| III_J_up | PRDM6 | 93166 | + | 5 | 122424841 | 122529960 | 1.82E-06 | 1.14E-02 | 3.42E-02 |
| III_J_up | CCDC141 | 285025 | - | 2 | 179694484 | 179914841 | 7.44E-07 | 6.98E-03 | 1.40E-02 |
| III_J_up | NRP1 | 8829 | - | 10 | 33466419 | 33623833 | 1.26E-10 | 2.36E-06 | 2.36E-06 |
| III_S-area | NRP1 | 8829 | - | 10 | 33466419 | 33623833 | 4.88E-10 | 4.58E-06 | 9.16E-06 |
| III_S-area | VTI1A | 143187 | + | 10 | 114206756 | 114578503 | 2.36E-10 | 4.43E-06 | 4.43E-06 |
| III_S-area | CCDC141 | 285025 | - | 2 | 179694484 | 179914841 | 4.64E-07 | 2.90E-03 | 8.70E-03 |
| I_RS | CAMK2D | 817 | - | 4 | 114372188 | 114683669 | 1.04E-07 | 9.79E-04 | 1.96E-03 |
| I_RS | KCND3 | 3752 | - | 1 | 112318444 | 112532147 | 1.47E-16 | 2.76E-12 | 2.76E-12 |
| I_RS | LOC643355 | 643355 | + | 1 | 112532593 | 112541464 | 2.72E-07 | 1.70E-03 | 5.11E-03 |
| III_t_R | CFAP36 | 112942 | + | 2 | 55746740 | 55772216 | 2.35E-06 | 4.40E-02 | 4.40E-02 |
| aVR_R | ADK | 132 | + | 10 | 75910943 | 76469061 | 6.55E-08 | 6.15E-04 | 1.23E-03 |
| aVR_R | KCND3 | 3752 | - | 1 | 112318444 | 112532147 | 2.46E-20 | 4.62E-16 | 4.62E-16 |
| aVR_R | LOC643355 | 643355 | + | 1 | 112532593 | 112541464 | 8.86E-07 | 4.15E-03 | 1.66E-02 |
| aVR_R | NRP1 | 8829 | - | 10 | 33466419 | 33623833 | 2.23E-07 | 1.40E-03 | 4.19E-03 |
| I_ST | LMF1 | 64788 | - | 16 | 903634 | 1031318 | 5.34E-08 | 5.01E-04 | 1.00E-03 |
| I_ST | SOX8 | 30812 | + | 16 | 1031808 | 1036979 | 2.57E-06 | 9.63E-03 | 4.81E-02 |
| I_ST | CCDC141 | 285025 | - | 2 | 179694484 | 179914841 | 1.63E-10 | 3.06E-06 | 3.06E-06 |
| I_ST | ATP5B | 506 | - | 12 | 57031959 | 57039852 | 1.41E-06 | 6.62E-03 | 2.65E-02 |
| I_ST | HSD17B6 | 8630 | + | 12 | 57146237 | 57181574 | 1.15E-06 | 6.62E-03 | 2.16E-02 |
| aVR_T | DEFB136 | 613210 | - | 8 | 11831446 | 11832108 | 1.53E-06 | 3.58E-03 | 2.87E-02 |
| aVR_T | SOX8 | 30812 | + | 16 | 1031808 | 1036979 | 2.68E-08 | 1.68E-04 | 5.03E-04 |
| aVR_T | CCDC141 | 285025 | - | 2 | 179694484 | 179914841 | 5.70E-07 | 1.78E-03 | 1.07E-02 |
| aVR_T | SOX5 | 6660 | - | 12 | 23682438 | 24715383 | 1.54E-09 | 1.44E-05 | 2.89E-05 |
| aVR_T | HSPB7 | 27129 | - | 1 | 16340523 | 16346084 | 4.26E-07 | 1.69E-03 | 8.00E-03 |
| aVR_T | PRKCA | 5578 | + | 17 | 64298760 | 64806862 | 4.49E-07 | 1.69E-03 | 8.43E-03 |
| aVR_T | DEFB134 | 613211 | - | 8 | 11851489 | 11853760 | 9.13E-07 | 2.45E-03 | 1.71E-02 |
| aVR_T | LMF1 | 64788 | - | 16 | 903634 | 1031318 | 5.37E-10 | 1.01E-05 | 1.01E-05 |
| aVR_RS | ADK | 132 | + | 10 | 75910943 | 76469061 | 1.27E-07 | 1.19E-03 | 2.37E-03 |
| aVR_RS | KCND3 | 3752 | - | 1 | 112318444 | 112532147 | 3.19E-19 | 5.98E-15 | 5.98E-15 |
| aVR_RS | LOC643355 | 643355 | + | 1 | 112532593 | 112541464 | 2.10E-06 | 9.83E-03 | 3.93E-02 |
| aVR_RS | NRP1 | 8829 | - | 10 | 33466419 | 33623833 | 1.31E-06 | 8.20E-03 | 2.46E-02 |
| aVR_ST | DEFB136 | 613210 | - | 8 | 11831446 | 11832108 | 6.86E-10 | 4.29E-06 | 1.29E-05 |
| aVR_ST | TBC1D3P6 | 1.02E+08 | + | 1 | 16302626 | 16323701 | 1.03E-06 | 1.93E-03 | 1.93E-02 |
| aVR_ST | CCDC141 | 285025 | - | 2 | 179694484 | 179914841 | 6.95E-11 | 6.52E-07 | 1.30E-06 |
| aVR_ST | DEFB134 | 613211 | - | 8 | 11851489 | 11853760 | 2.51E-08 | 6.22E-05 | 4.70E-04 |
| aVR_ST | HSPB7 | 27129 | - | 1 | 16340523 | 16346084 | 1.83E-11 | 3.44E-07 | 3.44E-07 |
| aVR_ST | XKR6 | 286046 | - | 8 | 10753654 | 11058875 | 2.65E-08 | 6.22E-05 | 4.97E-04 |
| aVR_ST | SOX8 | 30812 | + | 16 | 1031808 | 1036979 | 2.39E-07 | 4.97E-04 | 4.48E-03 |
| aVR_ST | CLCNKA | 1187 | + | 1 | 16348486 | 16360548 | 7.52E-09 | 2.35E-05 | 1.41E-04 |
| aVR_ST | ZBTB17 | 7709 | - | 1 | 16268364 | 16302627 | 1.43E-09 | 6.71E-06 | 2.68E-05 |
| aVR_ST | LMF1 | 64788 | - | 16 | 903634 | 1031318 | 1.83E-09 | 6.87E-06 | 3.43E-05 |
| aVR_J_up | FKBP7 | 51661 | - | 2 | 179328391 | 179343646 | 3.84E-11 | 1.20E-07 | 7.20E-07 |
| aVR_J_up | SPTBN1 | 6711 | + | 2 | 54683454 | 54898583 | 6.51E-07 | 7.38E-04 | 1.22E-02 |
| aVR_J_up | ERBB4 | 2066 | - | 2 | 212240442 | 213403879 | 4.45E-10 | 1.04E-06 | 8.34E-06 |
| aVR_J_up | DFNB59 | 494513 | + | 2 | 179316163 | 179326149 | 4.32E-10 | 1.04E-06 | 8.11E-06 |
| aVR_J_up | VTI1A | 143187 | + | 10 | 114206756 | 114578503 | 6.69E-07 | 7.38E-04 | 1.26E-02 |
| aVR_J_up | KCND3 | 3752 | - | 1 | 112318444 | 112532147 | 7.46E-09 | 1.17E-05 | 1.40E-04 |
| aVR_J_up | FOLH1 | 2346 | - | 11 | 49168187 | 49230222 | 1.31E-06 | 1.31E-03 | 2.45E-02 |
| aVR_J_up | LRCH1 | 23143 | + | 13 | 47127296 | 47327176 | 6.60E-10 | 1.24E-06 | 1.24E-05 |
| aVR_J_up | SCN5A | 6331 | - | 3 | 38589553 | 38691164 | 3.79E-16 | 7.12E-12 | 7.12E-12 |
| aVR_J_up | PRKRA | 8575 | - | 2 | 179296141 | 179315958 | 9.17E-12 | 4.30E-08 | 1.72E-07 |
| aVR_J_up | TTN | 7273 | - | 2 | 179390716 | 179672150 | 1.65E-14 | 1.55E-10 | 3.10E-10 |
| aVR_J_up | CDKN1A | 1026 | + | 6 | 36644237 | 36655116 | 2.43E-07 | 3.04E-04 | 4.56E-03 |
| aVR_J_up | PLEKHA3 | 65977 | + | 2 | 179345199 | 179369783 | 2.05E-11 | 7.70E-08 | 3.85E-07 |
| aVR_J_up | NKX2-5 | 1482 | - | 5 | 172659107 | 172662315 | 4.08E-09 | 6.97E-06 | 7.66E-05 |
| aVR_J_up | TBX3 | 6926 | - | 12 | 115108059 | 115121969 | 5.59E-10 | 1.17E-06 | 1.05E-05 |
| aVR_J_up | ADCY10 | 55811 | - | 1 | 167778357 | 167883608 | 1.33E-06 | 1.31E-03 | 2.50E-02 |
| aVR_J_up | SCN10A | 6336 | - | 3 | 38738837 | 38835501 | 1.97E-07 | 2.64E-04 | 3.69E-03 |
| aVR_J_up | TRDN | 10345 | - | 6 | 123537484 | 123958428 | 1.04E-08 | 1.50E-05 | 1.94E-04 |
| aVR_J_up | CCDC141 | 285025 | - | 2 | 179694484 | 179914841 | 4.11E-14 | 2.57E-10 | 7.72E-10 |
| aVR_R-area | ADK | 132 | + | 10 | 75910943 | 76469061 | 4.93E-07 | 3.09E-03 | 9.26E-03 |
| aVR_R-area | KCND3 | 3752 | - | 1 | 112318444 | 112532147 | 1.48E-17 | 2.77E-13 | 2.77E-13 |
| aVR_R-area | ARHGAP11A | 9824 | + | 15 | 32906720 | 32931868 | 2.50E-06 | 1.17E-02 | 4.68E-02 |
| aVR_R-area | LOC643355 | 643355 | + | 1 | 112532593 | 112541464 | 6.02E-08 | 5.64E-04 | 1.13E-03 |
| aVR_S-area | SCN10A | 6336 | - | 3 | 38738837 | 38835501 | 1.25E-07 | 2.35E-03 | 2.35E-03 |
| aVR_T-area | ZBTB17 | 7709 | - | 1 | 16268364 | 16302627 | 4.36E-07 | 9.32E-04 | 8.18E-03 |
| aVR_T-area | DPT | 1805 | - | 1 | 168664695 | 168698442 | 1.34E-08 | 8.38E-05 | 2.51E-04 |
| aVR_T-area | DEFB136 | 613210 | - | 8 | 11831446 | 11832108 | 4.47E-07 | 9.32E-04 | 8.39E-03 |
| aVR_T-area | HSPB7 | 27129 | - | 1 | 16340523 | 16346084 | 1.24E-07 | 3.87E-04 | 2.32E-03 |
| aVR_T-area | SSBP3 | 23648 | - | 1 | 54691104 | 54872068 | 2.20E-06 | 4.13E-03 | 4.13E-02 |
| aVR_T-area | LMF1 | 64788 | - | 16 | 903634 | 1031318 | 5.40E-09 | 5.07E-05 | 1.01E-04 |
| aVR_T-area | SOX8 | 30812 | + | 16 | 1031808 | 1036979 | 2.65E-07 | 7.09E-04 | 4.97E-03 |
| aVR_T-area | SOX5 | 6660 | - | 12 | 23682438 | 24715383 | 4.77E-08 | 2.24E-04 | 8.95E-04 |
| aVR_T-area | LITAF | 9516 | - | 16 | 11641578 | 11681322 | 5.97E-08 | 2.24E-04 | 1.12E-03 |
| aVR_T-area | PLCL1 | 5334 | + | 2 | 198669426 | 199014608 | 1.77E-09 | 3.32E-05 | 3.32E-05 |
| I_J_up | PRKRA | 8575 | - | 2 | 179296141 | 179315958 | 1.43E-07 | 3.83E-04 | 2.68E-03 |
| I_J_up | PLEKHA3 | 65977 | + | 2 | 179345199 | 179369783 | 1.45E-08 | 5.44E-05 | 2.72E-04 |
| I_J_up | PRDM6 | 93166 | + | 5 | 122424841 | 122529960 | 2.23E-08 | 6.98E-05 | 4.19E-04 |
| I_J_up | LOC100130370 | 1E+08 | - | 17 | 79349699 | 79359160 | 2.19E-06 | 3.40E-03 | 4.11E-02 |
| I_J_up | VTI1A | 143187 | + | 10 | 114206756 | 114578503 | 1.04E-13 | 9.76E-10 | 1.95E-09 |
| I_J_up | FKBP7 | 51661 | - | 2 | 179328391 | 179343646 | 7.58E-07 | 1.29E-03 | 1.42E-02 |
| I_J_up | TBX3 | 6926 | - | 12 | 115108059 | 115121969 | 2.35E-06 | 3.40E-03 | 4.42E-02 |
| I_J_up | IQSEC1 | 9922 | - | 3 | 12938542 | 13114652 | 2.77E-07 | 5.82E-04 | 5.20E-03 |
| I_J_up | TTN | 7273 | - | 2 | 179390716 | 179672150 | 2.37E-12 | 1.48E-08 | 4.45E-08 |
| I_J_up | SCN10A | 6336 | - | 3 | 38738837 | 38835501 | 5.86E-07 | 1.10E-03 | 1.10E-02 |
| I_J_up | KCND3 | 3752 | - | 1 | 112318444 | 112532147 | 7.24E-10 | 3.40E-06 | 1.36E-05 |
| I_J_up | LMF1 | 64788 | - | 16 | 903634 | 1031318 | 2.79E-07 | 5.82E-04 | 5.24E-03 |
| I_J_up | SCN5A | 6331 | - | 3 | 38589553 | 38691164 | 2.93E-14 | 5.50E-10 | 5.50E-10 |
| aVR_t_ST | KCND3 | 3752 | - | 1 | 112318444 | 112532147 | 2.62E-13 | 4.92E-09 | 4.92E-09 |
| aVR_t_ST | SCN5A | 6331 | - | 3 | 38589553 | 38691164 | 4.93E-08 | 4.63E-04 | 9.26E-04 |
| aVR_t_ST | NOS1AP | 9722 | + | 1 | 162039581 | 162339813 | 5.45E-07 | 3.41E-03 | 1.02E-02 |
| aVR_t_QT | NOS1AP | 9722 | + | 1 | 162039581 | 162339813 | 2.44E-08 | 2.29E-04 | 4.58E-04 |
| aVR_t_QT | SCN5A | 6331 | - | 3 | 38589553 | 38691164 | 1.74E-07 | 1.09E-03 | 3.27E-03 |
| aVR_t_QT | KCND3 | 3752 | - | 1 | 112318444 | 112532147 | 1.56E-10 | 2.93E-06 | 2.93E-06 |
| aVL_R | KCND3 | 3752 | - | 1 | 112318444 | 112532147 | 5.84E-07 | 1.10E-02 | 1.10E-02 |
| aVL_R | HAAO | 23498 | - | 2 | 42994229 | 43019753 | 1.28E-06 | 1.20E-02 | 2.41E-02 |
| aVL_S | CAMK2D | 817 | - | 4 | 114372188 | 114683669 | 2.79E-08 | 5.24E-04 | 5.24E-04 |
| aVL_S | AKR1A1 | 10327 | + | 1 | 46016455 | 46035723 | 2.40E-06 | 1.30E-02 | 4.51E-02 |
| aVL_S | CCDC141 | 285025 | - | 2 | 179694484 | 179914841 | 1.03E-06 | 9.63E-03 | 1.93E-02 |
| I_R-area | KCND3 | 3752 | - | 1 | 112318444 | 112532147 | 1.70E-14 | 3.18E-10 | 3.18E-10 |
| I_R-area | LOC643355 | 643355 | + | 1 | 112532593 | 112541464 | 3.47E-08 | 3.25E-04 | 6.51E-04 |
| aVL_RS | CAMK2D | 817 | - | 4 | 114372188 | 114683669 | 1.54E-06 | 2.88E-02 | 2.88E-02 |
| aVL_ST | HSD17B6 | 8630 | + | 12 | 57146237 | 57181574 | 1.86E-06 | 9.62E-03 | 3.48E-02 |
| aVL_ST | NMB | 4828 | - | 15 | 85198360 | 85201802 | 2.05E-06 | 9.62E-03 | 3.85E-02 |
| aVL_ST | CCDC141 | 285025 | - | 2 | 179694484 | 179914841 | 7.24E-08 | 6.79E-04 | 1.36E-03 |
| aVL_ST | CAMK2D | 817 | - | 4 | 114372188 | 114683669 | 1.48E-09 | 2.77E-05 | 2.77E-05 |
| aVL_J_up | LMF1 | 64788 | - | 16 | 903634 | 1031318 | 3.15E-07 | 1.48E-03 | 5.92E-03 |
| aVL_J_up | VTI1A | 143187 | + | 10 | 114206756 | 114578503 | 2.17E-08 | 1.36E-04 | 4.07E-04 |
| aVL_J_up | NRP1 | 8829 | - | 10 | 33466419 | 33623833 | 1.06E-08 | 9.98E-05 | 2.00E-04 |
| aVL_J_up | PRDM6 | 93166 | + | 5 | 122424841 | 122529960 | 1.29E-09 | 2.43E-05 | 2.43E-05 |
| aVL_T-area | CASQ2 | 845 | - | 1 | 116242624 | 116311426 | 1.84E-06 | 3.45E-02 | 3.45E-02 |
| aVL_t_S | G6PC3 | 92579 | + | 17 | 42148098 | 42153712 | 2.70E-07 | 5.07E-03 | 5.07E-03 |
| aVL_t_ST | CASQ2 | 845 | - | 1 | 116242624 | 116311426 | 7.36E-07 | 6.91E-03 | 1.38E-02 |
| aVL_t_ST | VTI1A | 143187 | + | 10 | 114206756 | 114578503 | 1.39E-07 | 2.60E-03 | 2.60E-03 |
| aVF_R | ADK | 132 | + | 10 | 75910943 | 76469061 | 2.27E-06 | 4.26E-02 | 4.26E-02 |
| aVF_S | SPTBN1 | 6711 | + | 2 | 54683454 | 54898583 | 2.65E-06 | 1.05E-02 | 4.97E-02 |
| aVF_S | CCDC141 | 285025 | - | 2 | 179694484 | 179914841 | 2.00E-07 | 1.88E-03 | 3.76E-03 |
| aVF_S | NRP1 | 8829 | - | 10 | 33466419 | 33623833 | 2.59E-10 | 4.87E-06 | 4.87E-06 |
| aVF_S | NFIA | 4774 | + | 1 | 61542946 | 61928460 | 9.70E-07 | 6.07E-03 | 1.82E-02 |
| aVF_T | CASP7 | 840 | + | 10 | 115438921 | 115490668 | 5.17E-07 | 9.70E-03 | 9.70E-03 |

Supplementary Table 6. Enriched gene ontologies (FDR < 0.05) for cluster3 in Figure S2A.

| ONTOLOGY | ID | Description | P value | Gene ID | Count | FDR | Bonferroni corrected P |
| --- | --- | --- | --- | --- | --- | --- | --- |
| BP | GO:0060048 | cardiac muscle contraction | 2.11E-05 | KCND3/SCN5A/TTN | 3 | 9.71E-03 | 1.22E-02 |
| BP | GO:0006941 | striated muscle contraction | 4.51E-05 | KCND3/SCN5A/TTN | 3 | 9.71E-03 | 2.61E-02 |
| BP | GO:0099625 | ventricular cardiac muscle cell membrane repolarization | 6.40E-05 | KCND3/SCN5A | 2 | 9.71E-03 | 3.70E-02 |
| BP | GO:0086005 | ventricular cardiac muscle cell action potential | 9.36E-05 | KCND3/SCN5A | 2 | 9.71E-03 | 5.41E-02 |
| BP | GO:0086091 | regulation of heart rate by cardiac conduction | 1.16E-04 | KCND3/SCN5A | 2 | 9.71E-03 | 6.73E-02 |
| BP | GO:0060047 | heart contraction | 1.19E-04 | KCND3/SCN5A/TTN | 3 | 9.71E-03 | 6.91E-02 |
| BP | GO:0099622 | cardiac muscle cell membrane repolarization | 1.23E-04 | KCND3/SCN5A | 2 | 9.71E-03 | 7.09E-02 |
| BP | GO:0003015 | heart process | 1.34E-04 | KCND3/SCN5A/TTN | 3 | 9.71E-03 | 7.77E-02 |
| BP | GO:0086009 | membrane repolarization | 2.08E-04 | KCND3/SCN5A | 2 | 1.30E-02 | 1.20E-01 |
| BP | GO:0086002 | cardiac muscle cell action potential involved in contraction | 2.24E-04 | KCND3/SCN5A | 2 | 1.30E-02 | 1.30E-01 |
| BP | GO:0006936 | muscle contraction | 3.31E-04 | KCND3/SCN5A/TTN | 3 | 1.74E-02 | 1.92E-01 |
| BP | GO:0086003 | cardiac muscle cell contraction | 4.33E-04 | KCND3/SCN5A | 2 | 2.03E-02 | 2.50E-01 |
| BP | GO:0086001 | cardiac muscle cell action potential | 4.56E-04 | KCND3/SCN5A | 2 | 2.03E-02 | 2.64E-01 |
| BP | GO:0003012 | muscle system process | 6.93E-04 | KCND3/SCN5A/TTN | 3 | 2.84E-02 | 4.01E-01 |
| BP | GO:0061337 | cardiac conduction | 7.38E-04 | KCND3/SCN5A | 2 | 2.84E-02 | 4.26E-01 |
| BP | GO:0070252 | actin-mediated cell contraction | 8.15E-04 | KCND3/SCN5A | 2 | 2.88E-02 | 4.71E-01 |
| BP | GO:0002027 | regulation of heart rate | 8.46E-04 | KCND3/SCN5A | 2 | 2.88E-02 | 4.89E-01 |
| BP | GO:0048640 | negative regulation of developmental growth | 9.45E-04 | CDKN1A/NRP1 | 2 | 3.03E-02 | 5.46E-01 |
| BP | GO:0030048 | actin filament-based movement | 1.35E-03 | KCND3/SCN5A | 2 | 4.11E-02 | 7.82E-01 |
| BP | GO:0001508 | action potential | 1.56E-03 | KCND3/SCN5A | 2 | 4.36E-02 | 9.02E-01 |
| BP | GO:0051592 | response to calcium ion | 1.58E-03 | SCN5A/TTN | 2 | 4.36E-02 | 9.15E-01 |
| CC | GO:0099055 | integral component of postsynaptic membrane | 9.83E-04 | NRP1/KCND3 | 2 | 1.44E-02 | 5.51E-02 |
| CC | GO:0098936 | intrinsic component of postsynaptic membrane | 1.07E-03 | NRP1/KCND3 | 2 | 1.44E-02 | 5.98E-02 |
| CC | GO:0030018 | Z disc | 1.14E-03 | SCN5A/TTN | 2 | 1.44E-02 | 6.37E-02 |
| CC | GO:0042383 | sarcolemma | 1.21E-03 | KCND3/SCN5A | 2 | 1.44E-02 | 6.77E-02 |
| CC | GO:0031674 | I band | 1.36E-03 | SCN5A/TTN | 2 | 1.44E-02 | 7.62E-02 |
| CC | GO:0099699 | integral component of synaptic membrane | 1.56E-03 | NRP1/KCND3 | 2 | 1.44E-02 | 8.74E-02 |
| CC | GO:0099240 | intrinsic component of synaptic membrane | 1.80E-03 | NRP1/KCND3 | 2 | 1.44E-02 | 1.01E-01 |
| CC | GO:0030017 | sarcomere | 3.04E-03 | SCN5A/TTN | 2 | 2.00E-02 | 1.70E-01 |
| CC | GO:0034703 | cation channel complex | 3.39E-03 | KCND3/SCN5A | 2 | 2.00E-02 | 1.90E-01 |
| CC | GO:0030016 | myofibril | 3.60E-03 | SCN5A/TTN | 2 | 2.00E-02 | 2.02E-01 |
| CC | GO:0043292 | contractile fiber | 3.92E-03 | SCN5A/TTN | 2 | 2.00E-02 | 2.20E-01 |
| CC | GO:0002116 | semaphorin receptor complex | 4.48E-03 | NRP1 | 1 | 2.09E-02 | 2.51E-01 |
| CC | GO:0045211 | postsynaptic membrane | 5.05E-03 | NRP1/KCND3 | 2 | 2.18E-02 | 2.83E-01 |
| CC | GO:0034702 | ion channel complex | 5.92E-03 | KCND3/SCN5A | 2 | 2.37E-02 | 3.31E-01 |
| CC | GO:0001518 | voltage-gated sodium channel complex | 6.92E-03 | SCN5A | 1 | 2.58E-02 | 3.88E-01 |
| CC | GO:0097060 | synaptic membrane | 9.38E-03 | NRP1/KCND3 | 2 | 3.15E-02 | 5.25E-01 |
| CC | GO:1902495 | transmembrane transporter complex | 9.57E-03 | KCND3/SCN5A | 2 | 3.15E-02 | 5.36E-01 |
| CC | GO:0034706 | sodium channel complex | 1.06E-02 | SCN5A | 1 | 3.15E-02 | 5.92E-01 |
| CC | GO:1990351 | transporter complex | 1.07E-02 | KCND3/SCN5A | 2 | 3.15E-02 | 5.98E-01 |
| CC | GO:1902555 | endoribonuclease complex | 1.38E-02 | PRKRA | 1 | 3.86E-02 | 7.73E-01 |
| MF | GO:0044325 | transmembrane transporter binding | 1.29E-03 | KCND3/SCN5A | 2 | 3.07E-02 | 1.04E-01 |
| MF | GO:0004713 | protein tyrosine kinase activity | 1.45E-03 | NRP1/TTN | 2 | 3.07E-02 | 1.18E-01 |
| MF | GO:0019838 | growth factor binding | 1.54E-03 | NRP1/SCN5A | 2 | 3.07E-02 | 1.25E-01 |
| MF | GO:0005516 | calmodulin binding | 3.15E-03 | SCN5A/TTN | 2 | 3.07E-02 | 2.55E-01 |
| MF | GO:0005244 | voltage-gated ion channel activity | 3.18E-03 | KCND3/SCN5A | 2 | 3.07E-02 | 2.58E-01 |
| MF | GO:0022832 | voltage-gated channel activity | 3.18E-03 | KCND3/SCN5A | 2 | 3.07E-02 | 2.58E-01 |
| MF | GO:0005528 | FK506 binding | 4.34E-03 | FKBP7 | 1 | 3.07E-02 | 3.51E-01 |
| MF | GO:0120016 | sphingolipid transfer activity | 4.34E-03 | PLEKHA3 | 1 | 3.07E-02 | 3.51E-01 |
| MF | GO:0097493 | structural molecule activity conferring elasticity | 4.77E-03 | TTN | 1 | 3.07E-02 | 3.86E-01 |
| MF | GO:0004861 | cyclin-dependent protein serine/threonine kinase inhibitor activity | 5.20E-03 | CDKN1A | 1 | 3.07E-02 | 4.21E-01 |
| MF | GO:0005527 | macrolide binding | 5.20E-03 | FKBP7 | 1 | 3.07E-02 | 4.21E-01 |
| MF | GO:0017154 | semaphorin receptor activity | 5.20E-03 | NRP1 | 1 | 3.07E-02 | 4.21E-01 |
| MF | GO:1902282 | voltage-gated potassium channel activity involved in ventricular cardiac muscle cell action potential repolarization | 5.20E-03 | KCND3 | 1 | 3.07E-02 | 4.21E-01 |
| MF | GO:0050998 | nitric-oxide synthase binding | 5.64E-03 | SCN5A | 1 | 3.07E-02 | 4.57E-01 |
| MF | GO:0015271 | outward rectifier potassium channel activity | 6.07E-03 | KCND3 | 1 | 3.07E-02 | 4.92E-01 |
| MF | GO:0051371 | muscle alpha-actinin binding | 6.50E-03 | TTN | 1 | 3.07E-02 | 5.27E-01 |
| MF | GO:0086008 | voltage-gated potassium channel activity involved in cardiac muscle cell action potential repolarization | 6.50E-03 | KCND3 | 1 | 3.07E-02 | 5.27E-01 |
| MF | GO:0031625 | ubiquitin protein ligase binding | 6.86E-03 | CDKN1A/SCN5A | 2 | 3.07E-02 | 5.55E-01 |
| MF | GO:0044389 | ubiquitin-like protein ligase binding | 7.73E-03 | CDKN1A/SCN5A | 2 | 3.07E-02 | 6.26E-01 |
| MF | GO:0046624 | sphingolipid transporter activity | 7.80E-03 | PLEKHA3 | 1 | 3.07E-02 | 6.32E-01 |
| MF | GO:0030506 | ankyrin binding | 8.23E-03 | SCN5A | 1 | 3.07E-02 | 6.66E-01 |
| MF | GO:0097001 | ceramide binding | 8.66E-03 | PLEKHA3 | 1 | 3.07E-02 | 7.01E-01 |
| MF | GO:0022836 | gated channel activity | 8.85E-03 | KCND3/SCN5A | 2 | 3.07E-02 | 7.17E-01 |
| MF | GO:0005261 | cation channel activity | 9.10E-03 | KCND3/SCN5A | 2 | 3.07E-02 | 7.37E-01 |
| MF | GO:0017134 | fibroblast growth factor binding | 9.95E-03 | SCN5A | 1 | 3.12E-02 | 8.06E-01 |
| MF | GO:0005248 | voltage-gated sodium channel activity | 1.04E-02 | SCN5A | 1 | 3.12E-02 | 8.41E-01 |
| MF | GO:0120014 | phospholipid transfer activity | 1.04E-02 | PLEKHA3 | 1 | 3.12E-02 | 8.41E-01 |
| MF | GO:0051393 | alpha-actinin binding | 1.21E-02 | TTN | 1 | 3.28E-02 | 9.81E-01 |
| MF | GO:0033218 | amide binding | 1.22E-02 | FKBP7/PLEKHA3 | 2 | 3.28E-02 | 9.89E-01 |
| MF | GO:0046625 | sphingolipid binding | 1.25E-02 | PLEKHA3 | 1 | 3.28E-02 | 1.00E+00 |
| MF | GO:0070273 | phosphatidylinositol-4-phosphate binding | 1.25E-02 | PLEKHA3 | 1 | 3.28E-02 | 1.00E+00 |
| MF | GO:0046873 | metal ion transmembrane transporter activity | 1.38E-02 | KCND3/SCN5A | 2 | 3.40E-02 | 1.00E+00 |
| MF | GO:0030291 | protein serine/threonine kinase inhibitor activity | 1.43E-02 | CDKN1A | 1 | 3.40E-02 | 1.00E+00 |
| MF | GO:0005216 | ion channel activity | 1.46E-02 | KCND3/SCN5A | 2 | 3.40E-02 | 1.00E+00 |
| MF | GO:0030332 | cyclin binding | 1.47E-02 | CDKN1A | 1 | 3.40E-02 | 1.00E+00 |
| MF | GO:0042805 | actinin binding | 1.55E-02 | TTN | 1 | 3.50E-02 | 1.00E+00 |
| MF | GO:0003755 | peptidyl-prolyl cis-trans isomerase activity | 1.64E-02 | FKBP7 | 1 | 3.58E-02 | 1.00E+00 |
| MF | GO:0016859 | cis-trans isomerase activity | 1.77E-02 | FKBP7 | 1 | 3.58E-02 | 1.00E+00 |
| MF | GO:0015267 | channel activity | 1.77E-02 | KCND3/SCN5A | 2 | 3.58E-02 | 1.00E+00 |
| MF | GO:0022803 | passive transmembrane transporter activity | 1.78E-02 | KCND3/SCN5A | 2 | 3.58E-02 | 1.00E+00 |
| MF | GO:0008307 | structural constituent of muscle | 1.81E-02 | TTN | 1 | 3.58E-02 | 1.00E+00 |
| MF | GO:0005272 | sodium channel activity | 1.98E-02 | SCN5A | 1 | 3.81E-02 | 1.00E+00 |
| MF | GO:0015026 | coreceptor activity | 2.02E-02 | NRP1 | 1 | 3.81E-02 | 1.00E+00 |
| MF | GO:0120013 | lipid transfer activity | 2.11E-02 | PLEKHA3 | 1 | 3.87E-02 | 1.00E+00 |
| MF | GO:0016538 | cyclin-dependent protein serine/threonine kinase regulator activity | 2.15E-02 | CDKN1A | 1 | 3.87E-02 | 1.00E+00 |
| MF | GO:0004714 | transmembrane receptor protein tyrosine kinase activity | 2.58E-02 | NRP1 | 1 | 4.54E-02 | 1.00E+00 |
| MF | GO:0005548 | phospholipid transporter activity | 2.66E-02 | PLEKHA3 | 1 | 4.57E-02 | 1.00E+00 |
| MF | GO:0043621 | protein self-association | 2.71E-02 | TTN | 1 | 4.57E-02 | 1.00E+00 |
| MF | GO:0097110 | scaffold protein binding | 2.88E-02 | SCN5A | 1 | 4.75E-02 | 1.00E+00 |
| MF | GO:0004860 | protein kinase inhibitor activity | 3.00E-02 | CDKN1A | 1 | 4.86E-02 | 1.00E+00 |

**Supplementary Table 7**. The sample sizes used for all the phenotypical analysis.

| **Phenotype** | **Abbreviation** | **Case** | **Control** | **Total** |
| --- | --- | --- | --- | --- |
| Electrocardiographic trace characteristics | ETCs | - | - | 38953 |
| A10 | drugs used in diabetes | 1421 | 37532 | 38953 |
| metformin | - | 1250 | 37703 | 38953 |
| diabetes | - | 1134 | 37819 | 38953 |
| Aortic aneurysm | AA | 36 | 38917 | 38953 |
| Atrial fibrillation and flutter | AF | 818 | 38135 | 38953 |
| Atherosclerosis | ATH | 33 | 38920 | 38953 |
| Cardiomyopathy | CM | 65 | 38888 | 38953 |
| Major coronary heart disease event | CHD | 1457 | 37496 | 38953 |
| Coronary atherosclerosis | CA | 1603 | 37350 | 38953 |
| Cardiovascular diseases | CVD | 9995 | 28958 | 38953 |
| Heart failure | HF | 177 | 38776 | 38953 |
| Ischemic heart disease | IHD | 1956 | 36997 | 38953 |
| Myocardial infarction | MI | 546 | 38407 | 38953 |
| Non-rheumatic valve diseases | NRVD | 223 | 38730 | 38953 |
| Peripheral artery disease | PAD | 67 | 38886 | 38953 |
| Venous thromboembolism | VTE | 370 | 38583 | 38953 |
| Angina pectoris | ANGINA | 1119 | 37834 | 38953 |

Supplementary Table 8. Phenotypic relationship between ETCs and medication use.

| **ETC** | **A10** | | **metformin** | |
| --- | --- | --- | --- | --- |
|  | **Beta** | **P-value** | **Beta** | **P-value** |
| I_R | 2.805 | 1.09E-01 | 5.227 | 3.08E-03 |
| I_T-area | 0.052 | 3.09E-02 | 0.064 | 8.66E-03 |
| aVF_RS | -4.066 | 3.83E-02 | -4.928 | 1.29E-02 |
| aVF_ST | -0.361 | 7.76E-01 | -0.778 | 5.43E-01 |
| aVF_J_up | -4.569 | 3.21E-02 | -5.793 | 7.12E-03 |
| aVF_R-area | -0.124 | 9.90E-06 | -0.138 | 9.64E-07 |
| aVF_S-area | 0.042 | 3.34E-01 | 0.034 | 4.45E-01 |
| aVF_T-area | -0.026 | 8.59E-02 | -0.015 | 3.07E-01 |
| aVF_t_T | 0.000 | 8.05E-01 | 0.002 | 1.82E-01 |
| aVF_t_S | -0.002 | 4.20E-01 | -0.003 | 1.24E-01 |
| aVF_t_R | -0.001 | 2.18E-01 | -0.001 | 5.41E-01 |
| aVF_t_ST | 0.003 | 4.11E-02 | 0.003 | 1.74E-02 |
| I_t_T | 0.008 | 4.60E-05 | 0.009 | 1.31E-05 |
| aVF_t_QT | 0.002 | 2.44E-01 | 0.003 | 6.19E-02 |
| V1_R | -5.174 | 1.24E-06 | -4.863 | 6.31E-06 |
| V1_S | 1.891 | 4.40E-01 | -0.104 | 9.66E-01 |
| V1_T | -2.496 | 2.33E-06 | -2.952 | 3.17E-08 |
| V1_RS | -3.283 | 2.58E-01 | -4.967 | 9.01E-02 |
| I_t_S | -0.002 | 1.48E-01 | -0.002 | 3.08E-01 |
| V1_ST | 3.018 | 2.76E-01 | 1.727 | 5.37E-01 |
| V1_J_up | -0.239 | 8.36E-01 | -0.864 | 4.58E-01 |
| V1_R-area | -0.086 | 1.38E-04 | -0.092 | 5.29E-05 |
| V1_S-area | 0.171 | 8.28E-02 | 0.134 | 1.78E-01 |
| V1_T-area | -0.171 | 5.64E-08 | -0.215 | 1.18E-11 |
| V1_t_T | -0.014 | 2.90E-10 | -0.017 | 3.51E-14 |
| V1_t_S | 0.001 | 9.43E-02 | 0.002 | 1.78E-02 |
| V1_t_R | 0.000 | 9.95E-01 | -0.002 | 2.92E-01 |
| V1_t_ST | -0.006 | 4.81E-07 | -0.008 | 9.99E-12 |
| V1_t_QT | -0.006 | 4.50E-03 | -0.010 | 6.08E-06 |
| I_t_R | 0.002 | 4.41E-02 | 0.002 | 4.51E-02 |
| V2_R | -3.313 | 9.24E-02 | -0.851 | 6.68E-01 |
| V2_S | 2.148 | 5.23E-01 | -0.708 | 8.35E-01 |
| V2_T | -7.212 | 5.80E-12 | -7.722 | 2.82E-13 |
| V2_RS | -1.166 | 7.78E-01 | -1.560 | 7.09E-01 |
| V2_ST | -5.606 | 1.54E-01 | -9.051 | 2.27E-02 |
| I_t_ST | 0.005 | 4.67E-04 | 0.005 | 1.90E-04 |
| V2_J_up | 1.882 | 1.42E-01 | 1.656 | 2.01E-01 |
| V2_R-area | -0.065 | 9.60E-02 | -0.019 | 6.26E-01 |
| V2_S-area | 0.038 | 7.78E-01 | -0.041 | 7.60E-01 |
| V2_T-area | 0.030 | 4.37E-02 | 0.032 | 3.54E-02 |
| V2_t_T | 0.004 | 3.97E-03 | 0.005 | 2.05E-03 |
| V2_t_S | 0.000 | 6.71E-01 | 0.002 | 1.37E-01 |
| V2_t_R | -0.002 | 7.39E-02 | -0.002 | 3.57E-02 |
| V2_t_ST | 0.004 | 7.37E-06 | 0.004 | 1.08E-05 |
| V2_t_QT | 0.003 | 9.90E-02 | 0.002 | 1.58E-01 |
| I_t_QT | 0.007 | 2.32E-05 | 0.008 | 8.26E-06 |
| V3_R | -7.049 | 8.41E-03 | -4.176 | 1.22E-01 |
| V3_S | -3.392 | 3.01E-01 | -7.814 | 1.81E-02 |
| V3_T | -9.938 | 1.11E-18 | -11.395 | 1.20E-23 |
| V3_RS | -10.441 | 1.03E-02 | -11.990 | 3.50E-03 |
| V3_ST | -14.996 | 1.01E-04 | -21.161 | 5.48E-08 |
| V3_J_up | 1.310 | 2.58E-01 | 0.601 | 6.07E-01 |
| V3_R-area | -0.143 | 1.19E-02 | -0.076 | 1.86E-01 |
| V3_S-area | -0.069 | 5.87E-01 | -0.225 | 8.00E-02 |
| V3_T-area | 0.080 | 5.67E-06 | 0.094 | 1.54E-07 |
| V3_t_T | 0.009 | 2.73E-12 | 0.010 | 7.33E-16 |
| V3_t_S | 0.001 | 4.32E-01 | 0.002 | 1.95E-01 |
| V3_t_R | 0.000 | 5.43E-01 | 0.000 | 9.35E-01 |
| V3_t_ST | 0.008 | 1.01E-16 | 0.008 | 1.02E-18 |
| V3_t_QT | 0.007 | 2.42E-09 | 0.008 | 1.16E-11 |
| V4_R | -26.171 | 8.24E-14 | -25.447 | 6.38E-13 |
| V4_S | 2.200 | 3.55E-01 | 0.155 | 9.49E-01 |
| V4_T | -10.189 | 2.63E-23 | -12.151 | 6.86E-32 |
| V4_RS | -23.971 | 1.09E-08 | -25.292 | 2.30E-09 |
| V4_ST | -9.222 | 9.90E-04 | -13.483 | 1.84E-06 |
| V4_J_up | -3.972 | 5.86E-02 | -4.516 | 3.31E-02 |
| V4_R-area | -0.562 | 7.95E-15 | -0.535 | 2.30E-13 |
| V4_S-area | 0.008 | 9.25E-01 | -0.042 | 6.13E-01 |
| V4_T-area | 0.042 | 4.00E-02 | 0.049 | 1.71E-02 |
| V4_t_T | 0.010 | 4.04E-12 | 0.010 | 1.27E-13 |
| V4_t_S | -0.005 | 2.91E-03 | -0.005 | 9.13E-03 |
| V4_t_R | 0.000 | 5.83E-01 | 0.000 | 7.97E-01 |
| V4_t_ST | 0.008 | 1.68E-12 | 0.008 | 1.81E-13 |
| V4_t_QT | 0.007 | 4.25E-10 | 0.008 | 3.86E-11 |
| V5_R | -20.524 | 1.10E-07 | -22.060 | 1.56E-08 |
| V5_S | 3.191 | 5.09E-02 | 2.591 | 1.16E-01 |
| V5_T | -8.429 | 1.05E-22 | -10.170 | 9.52E-32 |
| V5_RS | -17.333 | 1.75E-04 | -19.469 | 2.98E-05 |
| V5_ST | -6.410 | 9.44E-04 | -9.073 | 3.54E-06 |
| V5_J_up | -14.085 | 5.55E-05 | -15.192 | 1.65E-05 |
| V5_R-area | -0.412 | 1.17E-07 | -0.434 | 3.19E-08 |
| V5_S-area | -0.005 | 9.34E-01 | -0.031 | 6.01E-01 |
| I_S | -0.237 | 7.23E-01 | 0.811 | 2.30E-01 |
| II_R | -7.183 | 1.74E-04 | -8.386 | 1.42E-05 |
| V5_T-area | 0.042 | 8.08E-02 | 0.059 | 1.50E-02 |
| V5_t_T | 0.010 | 1.79E-09 | 0.012 | 1.02E-12 |
| V5_t_S | -0.005 | 3.22E-02 | -0.005 | 1.29E-02 |
| V5_t_R | 0.001 | 3.62E-01 | 0.001 | 1.68E-01 |
| V5_t_ST | 0.006 | 3.63E-06 | 0.008 | 6.11E-09 |
| V5_t_QT | 0.007 | 8.85E-07 | 0.008 | 3.68E-10 |
| II_S | -0.508 | 5.47E-01 | -0.439 | 6.06E-01 |
| V6_R | -8.399 | 1.66E-02 | -9.642 | 6.45E-03 |
| V6_S | 1.313 | 1.88E-01 | 1.709 | 8.95E-02 |
| V6_T | -6.346 | 1.27E-20 | -7.398 | 5.71E-27 |
| V6_RS | -7.086 | 8.00E-02 | -7.933 | 5.21E-02 |
| V6_ST | -6.100 | 1.68E-06 | -6.985 | 5.56E-08 |
| V6_J_up | -20.650 | 7.68E-08 | -21.737 | 2.09E-08 |
| V6_R-area | -0.149 | 3.46E-02 | -0.165 | 2.02E-02 |
| V6_S-area | -0.036 | 4.24E-01 | -0.027 | 5.54E-01 |
| V6_T-area | 0.050 | 4.41E-02 | 0.068 | 6.30E-03 |
| II_T | -4.411 | 2.85E-17 | -5.012 | 1.81E-21 |
| V6_t_T | 0.011 | 4.74E-09 | 0.013 | 1.06E-11 |
| V6_t_S | -0.003 | 2.11E-01 | -0.002 | 2.83E-01 |
| V6_t_R | 0.002 | 6.15E-02 | 0.002 | 1.36E-02 |
| V6_t_ST | 0.006 | 1.68E-04 | 0.007 | 1.30E-05 |
| V6_t_QT | 0.007 | 3.93E-06 | 0.009 | 3.67E-08 |
| II_RS | -7.691 | 1.05E-03 | -8.825 | 1.96E-04 |
| II_ST | -5.620 | 1.71E-07 | -6.362 | 4.54E-09 |
| II_J_up | -7.902 | 2.47E-03 | -9.001 | 6.38E-04 |
| II_R-area | -0.120 | 2.27E-03 | -0.130 | 9.72E-04 |
| II_S-area | -0.053 | 1.69E-01 | -0.069 | 7.58E-02 |
| II_T-area | -0.002 | 9.19E-01 | 0.007 | 6.62E-01 |
| I_T | -2.911 | 7.28E-11 | -3.050 | 1.36E-11 |
| II_t_T | 0.004 | 5.07E-03 | 0.005 | 3.75E-04 |
| II_t_S | -0.001 | 6.05E-01 | -0.002 | 2.77E-01 |
| II_t_R | 0.002 | 1.08E-01 | 0.003 | 1.09E-02 |
| II_t_ST | 0.005 | 2.92E-04 | 0.005 | 2.52E-04 |
| II_t_QT | 0.006 | 1.38E-04 | 0.007 | 8.72E-06 |
| III_R | -4.980 | 1.91E-07 | -5.559 | 8.46E-09 |
| III_S | 10.227 | 9.91E-08 | 12.455 | 1.29E-10 |
| III_T | -1.237 | 7.11E-05 | -1.338 | 2.07E-05 |
| III_RS | 5.247 | 2.55E-02 | 6.896 | 3.63E-03 |
| III_ST | 10.017 | 2.31E-07 | 11.548 | 3.48E-09 |
| III_J_up | -1.667 | 3.86E-01 | -1.331 | 4.93E-01 |
| III_R-area | -0.063 | 5.38E-03 | -0.064 | 5.13E-03 |
| III_S-area | 0.266 | 2.53E-05 | 0.301 | 2.29E-06 |
| III_T-area | -0.106 | 3.63E-04 | -0.072 | 1.69E-02 |
| III_t_T | -0.009 | 1.11E-03 | -0.006 | 3.97E-02 |
| I_RS | 2.567 | 2.08E-01 | 6.039 | 3.33E-03 |
| III_t_S | -0.002 | 2.16E-01 | -0.004 | 2.13E-02 |
| III_t_R | 0.002 | 1.54E-01 | 0.003 | 1.97E-02 |
| III_t_ST | -0.001 | 4.16E-01 | 0.000 | 7.10E-01 |
| III_t_QT | 0.001 | 6.49E-01 | 0.003 | 1.52E-01 |
| aVR_R | 4.576 | 5.65E-03 | 4.309 | 9.84E-03 |
| aVR_S | -1.315 | 6.28E-03 | -0.875 | 7.18E-02 |
| I_ST | -3.897 | 1.81E-06 | -3.134 | 1.43E-04 |
| aVR_T | -3.644 | 1.04E-16 | -3.784 | 1.36E-17 |
| aVR_RS | 5.892 | 1.66E-03 | 5.184 | 6.13E-03 |
| aVR_ST | 5.668 | 1.79E-16 | 5.481 | 3.04E-15 |
| aVR_J_up | 5.194 | 1.47E-02 | 4.562 | 3.38E-02 |
| aVR_R-area | 0.052 | 1.41E-01 | 0.040 | 2.65E-01 |
| aVR_S-area | -0.071 | 6.09E-03 | -0.057 | 3.01E-02 |
| aVR_T-area | -0.607 | 1.74E-27 | -0.667 | 2.60E-32 |
| aVR_t_T | -0.023 | 4.15E-17 | -0.026 | 7.04E-20 |
| aVR_t_S | -0.005 | 6.38E-03 | -0.005 | 6.28E-03 |
| I_J_up | -2.687 | 2.97E-01 | -1.638 | 5.29E-01 |
| aVR_t_R | 0.003 | 1.24E-02 | 0.003 | 7.56E-03 |
| aVR_t_ST | -0.008 | 1.41E-04 | -0.010 | 2.85E-06 |
| aVR_t_QT | -0.006 | 1.19E-02 | -0.008 | 6.49E-04 |
| aVL_R | 7.515 | 1.27E-09 | 9.488 | 3.08E-14 |
| aVL_S | -0.661 | 4.45E-01 | -0.077 | 9.29E-01 |
| aVL_T | -1.331 | 4.70E-06 | -1.185 | 5.46E-05 |
| I_R-area | 0.123 | 2.78E-03 | 0.179 | 1.58E-05 |
| aVL_RS | 6.855 | 4.25E-05 | 9.411 | 2.58E-08 |
| aVL_ST | -2.673 | 3.26E-03 | -1.963 | 3.23E-02 |
| aVL_J_up | -0.454 | 8.36E-01 | 0.452 | 8.38E-01 |
| aVL_R-area | 0.193 | 1.22E-11 | 0.239 | 8.77E-17 |
| aVL_S-area | 0.010 | 7.05E-01 | 0.049 | 7.63E-02 |
| aVL_T-area | 0.069 | 3.88E-03 | 0.066 | 6.34E-03 |
| aVL_t_T | 0.012 | 8.06E-06 | 0.011 | 3.68E-05 |
| aVL_t_S | 0.003 | 5.49E-02 | 0.005 | 1.95E-03 |
| aVL_t_R | 0.000 | 9.14E-01 | 0.000 | 8.20E-01 |
| I_S-area | -0.006 | 8.29E-01 | 0.035 | 2.24E-01 |
| aVL_t_ST | 0.003 | 4.68E-02 | 0.004 | 7.30E-03 |
| aVL_t_QT | 0.003 | 1.42E-01 | 0.003 | 1.13E-01 |
| aVF_R | -6.630 | 7.79E-07 | -7.904 | 5.36E-09 |
| aVF_S | 2.563 | 2.69E-02 | 2.975 | 1.09E-02 |
| aVF_T | -2.803 | 8.36E-14 | -3.349 | 9.95E-19 |

Supplementary Table 9. The heritability ($h^{2}$) of ETCs calculated by single trait LDSC.

| **ETC** | ***h^2^*** | ***h^2^ se*** | **Lambda gc** | **mean chi^2^** | **Intercept** | **Intercept *se*** |
| --- | --- | --- | --- | --- | --- | --- |
| I_R | 0.119 | 0.031 | 1.047 | 1.071 | 1.003 | 0.009 |
| I_T-area | 0.042 | 0.016 | 0.999 | 1.013 | 0.990 | 0.006 |
| aVF_RS | 0.138 | 0.020 | 1.047 | 1.087 | 1.011 | 0.007 |
| aVF_ST | 0.130 | 0.025 | 1.047 | 1.075 | 1.002 | 0.008 |
| aVF_J_up | 0.105 | 0.020 | 1.047 | 1.072 | 1.015 | 0.007 |
| aVF_R-area | 0.081 | 0.018 | 1.047 | 1.053 | 1.009 | 0.007 |
| aVF_S-area | 0.106 | 0.023 | 1.047 | 1.062 | 1.004 | 0.008 |
| aVF_T-area | 0.023 | 0.016 | 0.999 | 1.011 | 0.998 | 0.007 |
| aVF_t_T | 0.025 | 0.016 | 0.999 | 1.008 | 0.994 | 0.006 |
| aVF_t_S | 0.053 | 0.020 | 0.999 | 1.025 | 0.996 | 0.006 |
| aVF_t_R | 0.047 | 0.019 | 1.047 | 1.027 | 1.002 | 0.007 |
| aVF_t_ST | 0.047 | 0.017 | 0.999 | 1.026 | 1.001 | 0.007 |
| I_t_T | 0.037 | 0.016 | 0.999 | 1.014 | 0.994 | 0.006 |
| aVF_t_QT | 0.068 | 0.019 | 1.047 | 1.031 | 0.993 | 0.007 |
| V1_R | 0.115 | 0.021 | 1.047 | 1.083 | 1.018 | 0.006 |
| V1_S | 0.103 | 0.019 | 1.047 | 1.069 | 1.011 | 0.007 |
| V1_T | 0.052 | 0.017 | 0.999 | 1.020 | 0.991 | 0.006 |
| V1_RS | 0.096 | 0.018 | 1.047 | 1.067 | 1.014 | 0.007 |
| I_t_S | 0.037 | 0.019 | 0.999 | 1.017 | 0.996 | 0.007 |
| V1_ST | 0.087 | 0.018 | 1.047 | 1.057 | 1.009 | 0.007 |
| V1_J_up | 0.035 | 0.016 | 1.047 | 1.021 | 1.002 | 0.007 |
| V1_R-area | 0.102 | 0.020 | 1.047 | 1.072 | 1.015 | 0.007 |
| V1_S-area | 0.073 | 0.018 | 1.047 | 1.042 | 1.002 | 0.007 |
| V1_T-area | 0.076 | 0.018 | 1.047 | 1.041 | 1.000 | 0.007 |
| V1_t_T | 0.075 | 0.017 | 1.047 | 1.042 | 1.001 | 0.007 |
| V1_t_S | 0.025 | 0.016 | 0.999 | 1.017 | 1.003 | 0.006 |
| V1_t_R | 0.043 | 0.015 | 1.047 | 1.035 | 1.012 | 0.006 |
| V1_t_ST | 0.041 | 0.017 | 0.999 | 1.022 | 1.000 | 0.007 |
| V1_t_QT | 0.035 | 0.015 | 0.999 | 1.027 | 1.008 | 0.006 |
| I_t_R | 0.085 | 0.018 | 1.047 | 1.037 | 0.989 | 0.007 |
| V2_R | 0.142 | 0.020 | 1.047 | 1.079 | 1.001 | 0.007 |
| V2_S | 0.081 | 0.020 | 1.047 | 1.049 | 1.004 | 0.007 |
| V2_T | 0.084 | 0.019 | 1.047 | 1.054 | 1.008 | 0.006 |
| V2_RS | 0.089 | 0.019 | 1.047 | 1.053 | 1.004 | 0.007 |
| V2_ST | 0.086 | 0.019 | 1.047 | 1.048 | 1.000 | 0.007 |
| I_t_ST | 0.094 | 0.023 | 1.047 | 1.041 | 0.989 | 0.008 |
| V2_J_up | 0.010 | 0.015 | 0.999 | 1.006 | 1.001 | 0.007 |
| V2_R-area | 0.140 | 0.022 | 1.047 | 1.076 | 0.999 | 0.007 |
| V2_S-area | 0.064 | 0.016 | 1.047 | 1.033 | 0.999 | 0.007 |
| V2_T-area | 0.009 | 0.016 | 0.999 | 1.003 | 0.998 | 0.006 |
| V2_t_T | 0.029 | 0.016 | 0.999 | 1.017 | 1.000 | 0.006 |
| V2_t_S | 0.032 | 0.017 | 1.047 | 1.031 | 1.014 | 0.006 |
| V2_t_R | -0.001 | 0.018 | 0.999 | 1.010 | 1.011 | 0.006 |
| V2_t_ST | 0.009 | 0.017 | 0.999 | 1.009 | 1.004 | 0.006 |
| V2_t_QT | 0.012 | 0.017 | 0.999 | 1.013 | 1.006 | 0.006 |
| I_t_QT | 0.035 | 0.016 | 1.047 | 1.024 | 1.004 | 0.006 |
| V3_R | 0.141 | 0.022 | 1.047 | 1.079 | 1.001 | 0.006 |
| V3_S | 0.142 | 0.020 | 1.047 | 1.087 | 1.009 | 0.008 |
| V3_T | 0.124 | 0.020 | 1.047 | 1.073 | 1.005 | 0.007 |
| V3_RS | 0.140 | 0.021 | 1.096 | 1.087 | 1.009 | 0.007 |
| V3_ST | 0.141 | 0.019 | 1.096 | 1.081 | 1.004 | 0.007 |
| V3_J_up | 0.036 | 0.017 | 0.999 | 1.011 | 0.991 | 0.006 |
| V3_R-area | 0.148 | 0.021 | 1.047 | 1.079 | 0.997 | 0.006 |
| V3_S-area | 0.108 | 0.017 | 1.047 | 1.057 | 0.998 | 0.007 |
| V3_T-area | -0.020 | 0.016 | 0.999 | 0.993 | 1.005 | 0.007 |
| V3_t_T | -0.006 | 0.016 | 0.999 | 1.008 | 1.012 | 0.007 |
| V3_t_S | 0.140 | 0.024 | 1.047 | 1.069 | 0.991 | 0.008 |
| V3_t_R | 0.048 | 0.016 | 0.999 | 1.027 | 1.001 | 0.006 |
| V3_t_ST | 0.025 | 0.016 | 0.999 | 1.019 | 1.005 | 0.007 |
| V3_t_QT | 0.037 | 0.014 | 0.999 | 1.022 | 1.002 | 0.006 |
| V4_R | 0.105 | 0.019 | 1.047 | 1.065 | 1.006 | 0.006 |
| V4_S | 0.174 | 0.024 | 1.096 | 1.120 | 1.024 | 0.008 |
| V4_T | 0.111 | 0.020 | 1.047 | 1.070 | 1.009 | 0.007 |
| V4_RS | 0.136 | 0.023 | 1.096 | 1.092 | 1.017 | 0.007 |
| V4_ST | 0.166 | 0.022 | 1.096 | 1.109 | 1.017 | 0.007 |
| V4_J_up | 0.082 | 0.017 | 1.047 | 1.042 | 0.996 | 0.006 |
| V4_R-area | 0.119 | 0.019 | 1.047 | 1.071 | 1.005 | 0.007 |
| V4_S-area | 0.112 | 0.021 | 1.047 | 1.073 | 1.011 | 0.008 |
| V4_T-area | 0.007 | 0.016 | 0.999 | 1.014 | 1.010 | 0.007 |
| V4_t_T | 0.018 | 0.016 | 0.999 | 1.022 | 1.012 | 0.006 |
| V4_t_S | 0.136 | 0.022 | 1.047 | 1.074 | 0.996 | 0.007 |
| V4_t_R | 0.019 | 0.017 | 0.999 | 1.020 | 1.010 | 0.006 |
| V4_t_ST | 0.040 | 0.017 | 0.999 | 1.022 | 0.999 | 0.006 |
| V4_t_QT | 0.026 | 0.017 | 0.999 | 1.019 | 1.004 | 0.006 |
| V5_R | 0.111 | 0.020 | 1.047 | 1.061 | 0.999 | 0.007 |
| V5_S | 0.152 | 0.023 | 1.096 | 1.103 | 1.018 | 0.008 |
| V5_T | 0.110 | 0.018 | 1.047 | 1.062 | 1.001 | 0.006 |
| V5_RS | 0.139 | 0.022 | 1.096 | 1.086 | 1.009 | 0.008 |
| V5_ST | 0.132 | 0.021 | 1.047 | 1.081 | 1.007 | 0.008 |
| V5_J_up | 0.155 | 0.023 | 1.096 | 1.097 | 1.009 | 0.008 |
| V5_R-area | 0.105 | 0.021 | 1.047 | 1.061 | 1.002 | 0.007 |
| V5_S-area | 0.117 | 0.021 | 1.047 | 1.075 | 1.010 | 0.007 |
| I_S | 0.070 | 0.019 | 1.047 | 1.030 | 0.991 | 0.007 |
| II_R | 0.110 | 0.019 | 1.047 | 1.067 | 1.007 | 0.007 |
| V5_T-area | 0.021 | 0.016 | 0.999 | 1.012 | 1.001 | 0.006 |
| V5_t_T | 0.015 | 0.016 | 0.999 | 1.015 | 1.007 | 0.006 |
| V5_t_S | 0.103 | 0.020 | 1.047 | 1.054 | 0.996 | 0.007 |
| V5_t_R | 0.087 | 0.022 | 1.047 | 1.041 | 0.993 | 0.007 |
| V5_t_ST | 0.064 | 0.018 | 1.047 | 1.037 | 1.002 | 0.007 |
| V5_t_QT | 0.037 | 0.018 | 0.999 | 1.020 | 1.000 | 0.007 |
| II_S | 0.093 | 0.019 | 1.047 | 1.064 | 1.012 | 0.007 |
| V6_R | 0.126 | 0.021 | 1.047 | 1.069 | 0.999 | 0.008 |
| V6_S | 0.090 | 0.020 | 1.047 | 1.063 | 1.014 | 0.007 |
| V6_T | 0.084 | 0.018 | 1.047 | 1.054 | 1.007 | 0.006 |
| V6_RS | 0.132 | 0.021 | 1.047 | 1.076 | 1.002 | 0.008 |
| V6_ST | 0.072 | 0.023 | 1.047 | 1.047 | 1.006 | 0.008 |
| V6_J_up | 0.190 | 0.026 | 1.096 | 1.127 | 1.020 | 0.008 |
| V6_R-area | 0.123 | 0.021 | 1.047 | 1.067 | 0.999 | 0.007 |
| V6_S-area | 0.089 | 0.019 | 1.047 | 1.050 | 1.001 | 0.007 |
| V6_T-area | 0.021 | 0.016 | 0.999 | 1.013 | 1.002 | 0.006 |
| II_T | 0.104 | 0.019 | 1.047 | 1.054 | 0.996 | 0.007 |
| V6_t_T | 0.038 | 0.018 | 1.047 | 1.023 | 1.002 | 0.006 |
| V6_t_S | 0.093 | 0.021 | 1.047 | 1.046 | 0.995 | 0.007 |
| V6_t_R | 0.049 | 0.019 | 0.999 | 1.027 | 0.999 | 0.007 |
| V6_t_ST | 0.084 | 0.019 | 1.047 | 1.049 | 1.002 | 0.007 |
| V6_t_QT | 0.060 | 0.017 | 0.999 | 1.026 | 0.993 | 0.007 |
| II_RS | 0.131 | 0.020 | 1.047 | 1.081 | 1.009 | 0.007 |
| II_ST | 0.108 | 0.024 | 1.047 | 1.058 | 0.997 | 0.008 |
| II_J_up | 0.151 | 0.022 | 1.096 | 1.100 | 1.018 | 0.007 |
| II_R-area | 0.094 | 0.018 | 1.047 | 1.058 | 1.006 | 0.006 |
| II_S-area | 0.096 | 0.021 | 1.047 | 1.056 | 1.002 | 0.007 |
| II_T-area | 0.003 | 0.017 | 0.999 | 1.000 | 0.999 | 0.007 |
| I_T | 0.116 | 0.030 | 1.047 | 1.063 | 0.997 | 0.009 |
| II_t_T | 0.012 | 0.018 | 0.999 | 1.004 | 0.998 | 0.007 |
| II_t_S | 0.054 | 0.017 | 1.047 | 1.038 | 1.008 | 0.006 |
| II_t_R | 0.062 | 0.020 | 0.999 | 1.030 | 0.995 | 0.007 |
| II_t_ST | 0.037 | 0.017 | 0.999 | 1.028 | 1.008 | 0.006 |
| II_t_QT | 0.051 | 0.018 | 0.999 | 1.020 | 0.992 | 0.006 |
| III_R | 0.108 | 0.018 | 1.047 | 1.061 | 1.001 | 0.007 |
| III_S | 0.164 | 0.020 | 1.096 | 1.102 | 1.012 | 0.007 |
| III_T | 0.046 | 0.017 | 1.047 | 1.033 | 1.008 | 0.007 |
| III_RS | 0.171 | 0.022 | 1.096 | 1.101 | 1.005 | 0.008 |
| III_ST | 0.167 | 0.022 | 1.096 | 1.097 | 1.005 | 0.008 |
| III_J_up | 0.121 | 0.021 | 1.047 | 1.069 | 1.001 | 0.008 |
| III_R-area | 0.073 | 0.017 | 1.047 | 1.036 | 0.997 | 0.007 |
| III_S-area | 0.120 | 0.023 | 1.047 | 1.072 | 1.005 | 0.008 |
| III_T-area | 0.047 | 0.019 | 1.047 | 1.031 | 1.005 | 0.007 |
| III_t_T | 0.066 | 0.019 | 1.047 | 1.037 | 1.001 | 0.007 |
| I_RS | 0.116 | 0.023 | 1.047 | 1.067 | 1.001 | 0.008 |
| III_t_S | 0.064 | 0.016 | 1.047 | 1.033 | 0.998 | 0.006 |
| III_t_R | 0.031 | 0.015 | 0.999 | 1.020 | 1.003 | 0.006 |
| III_t_ST | 0.043 | 0.018 | 1.047 | 1.030 | 1.006 | 0.007 |
| III_t_QT | 0.029 | 0.019 | 0.999 | 1.018 | 1.002 | 0.007 |
| aVR_R | 0.100 | 0.019 | 1.047 | 1.069 | 1.013 | 0.007 |
| aVR_S | 0.072 | 0.024 | 1.047 | 1.045 | 1.004 | 0.008 |
| I_ST | 0.095 | 0.025 | 1.047 | 1.049 | 0.996 | 0.008 |
| aVR_T | 0.119 | 0.025 | 1.047 | 1.065 | 0.999 | 0.008 |
| aVR_RS | 0.107 | 0.018 | 1.047 | 1.072 | 1.013 | 0.007 |
| aVR_ST | 0.109 | 0.034 | 1.047 | 1.058 | 0.997 | 0.010 |
| aVR_J_up | 0.192 | 0.027 | 1.096 | 1.122 | 1.016 | 0.008 |
| aVR_R-area | 0.108 | 0.020 | 1.047 | 1.063 | 1.003 | 0.007 |
| aVR_S-area | 0.064 | 0.017 | 1.047 | 1.038 | 1.003 | 0.007 |
| aVR_T-area | 0.112 | 0.024 | 1.047 | 1.066 | 1.003 | 0.008 |
| aVR_t_T | 0.053 | 0.017 | 1.047 | 1.031 | 1.002 | 0.007 |
| aVR_t_S | 0.023 | 0.019 | 0.999 | 1.023 | 1.010 | 0.007 |
| I_J_up | 0.192 | 0.029 | 1.096 | 1.116 | 1.009 | 0.008 |
| aVR_t_R | 0.051 | 0.017 | 1.047 | 1.027 | 0.998 | 0.007 |
| aVR_t_ST | 0.120 | 0.021 | 1.047 | 1.071 | 1.005 | 0.007 |
| aVR_t_QT | 0.107 | 0.019 | 1.047 | 1.072 | 1.013 | 0.007 |
| aVL_R | 0.114 | 0.026 | 1.047 | 1.057 | 0.992 | 0.008 |
| aVL_S | 0.082 | 0.017 | 1.047 | 1.037 | 0.992 | 0.006 |
| aVL_T | 0.064 | 0.019 | 1.047 | 1.037 | 1.002 | 0.007 |
| I_R-area | 0.124 | 0.025 | 1.047 | 1.063 | 0.993 | 0.008 |
| aVL_RS | 0.112 | 0.024 | 1.047 | 1.056 | 0.993 | 0.008 |
| aVL_ST | 0.078 | 0.018 | 1.047 | 1.035 | 0.992 | 0.006 |
| aVL_J_up | 0.184 | 0.024 | 1.096 | 1.109 | 1.005 | 0.008 |
| aVL_R-area | 0.098 | 0.020 | 1.047 | 1.044 | 0.989 | 0.007 |
| aVL_S-area | 0.059 | 0.017 | 0.999 | 1.022 | 0.989 | 0.006 |
| aVL_T-area | 0.059 | 0.017 | 1.047 | 1.029 | 0.996 | 0.006 |
| aVL_t_T | 0.042 | 0.017 | 0.999 | 1.025 | 1.002 | 0.006 |
| aVL_t_S | 0.038 | 0.016 | 0.999 | 1.016 | 0.995 | 0.007 |
| aVL_t_R | 0.089 | 0.019 | 1.047 | 1.037 | 0.987 | 0.007 |
| I_S-area | 0.047 | 0.018 | 1.047 | 1.021 | 0.995 | 0.007 |
| aVL_t_ST | 0.050 | 0.019 | 0.999 | 1.025 | 0.997 | 0.007 |
| aVL_t_QT | 0.049 | 0.019 | 0.999 | 1.023 | 0.996 | 0.007 |
| aVF_R | 0.100 | 0.020 | 1.047 | 1.061 | 1.006 | 0.007 |
| aVF_S | 0.118 | 0.022 | 1.047 | 1.079 | 1.014 | 0.007 |
| aVF_T | 0.087 | 0.017 | 1.047 | 1.055 | 1.007 | 0.006 |

Supplementary Table 10. The heritability ($h^{2}$) of cardiovascular diseases calculated by single trait LDSC.

| **GWAS id** | $\boldsymbol{h}^{\boldsymbol{2}}\boldsymbol{\pm se}$ | **Lambda gc** | **Mean** $\boldsymbol{\chi}^{\boldsymbol{2}}$ | **Intercept**$\boldsymbol{\pm se}$ |
| --- | --- | --- | --- | --- |
| I9_AF | 0.063$\pm$0.010 | 1.159 | 1.244 | 1.067$\pm$0.012 |
| I9_AF_REIMB | 0.085$\pm$0.015 | 1.121 | 1.182 | 1.043$\pm$0.011 |
| I9_AORTANEUR | 0.089$\pm$0.026 | 1.053 | 1.058 | 1.020$\pm$0.008 |
| I9_ATHSCLE | 0.064$\pm$0.012 | 1.086 | 1.100 | 1.035$\pm$0.008 |
| I9_CARDMPRI | 0.051$\pm$0.019 | 1.047 | 1.045 | 1.017$\pm$0.008 |
| I9_CARDMYO | 0.047$\pm$0.014 | 1.053 | 1.057 | 1.021$\pm$0.008 |
| I9_CHD | 0.053$\pm$0.009 | 1.146 | 1.203 | 1.098$\pm$0.011 |
| I9_CHD_NOREV | 0.051$\pm$0.009 | 1.127 | 1.161 | 1.075$\pm$0.010 |
| I9_CORATHER | 0.039$\pm$0.006 | 1.178 | 1.230 | 1.109$\pm$0.012 |
| I9_CVD | 0.026$\pm$0.003 | 1.210 | 1.247 | 1.129$\pm$0.010 |
| I9_HEARTFAIL | 0.023$\pm$0.005 | 1.080 | 1.091 | 1.043$\pm$0.009 |
| I9_HEARTFAIL_ALLCAUSE | 0.018$\pm$0.003 | 1.108 | 1.125 | 1.056$\pm$0.009 |
| I9_HEARTFAIL_NS | 0.018$\pm$0.003 | 1.111 | 1.125 | 1.056$\pm$0.009 |
| I9_IHD | 0.038$\pm$0.006 | 1.194 | 1.240 | 1.119$\pm$0.011 |
| I9_ISCHHEART | 0.037$\pm$0.006 | 1.178 | 1.228 | 1.114$\pm$0.011 |
| I9_MI | 0.051$\pm$0.008 | 1.121 | 1.161 | 1.064$\pm$0.009 |
| I9_MI_STRICT | 0.057$\pm$0.009 | 1.111 | 1.156 | 1.056$\pm$0.009 |
| I9_NONRHEVALV | 0.028$\pm$0.006 | 1.062 | 1.078 | 1.036$\pm$0.007 |
| I9_PAD | 0.066$\pm$0.013 | 1.071 | 1.091 | 1.030$\pm$0.008 |
| I9_VHD | 0.015$\pm$0.003 | 1.102 | 1.120 | 1.056$\pm$0.008 |
| I9_VTE | 0.030$\pm$0.009 | 1.071 | 1.106 | 1.040$\pm$0.010 |
| I9_ANGINA | 0.046$\pm$0.007 | 1.162 | 1.192 | 1.089$\pm$0.010 |

Supplementary Table 11. the significant genetic correlation ($r_{g}$) between ETCs and medication use by cross-trait LDSC. *se*: standard error.

| **ETC** | **Medication** | ***r_g_*** | ***se*** | **P-value** | **Bonferroni corrected P-value** |
| --- | --- | --- | --- | --- | --- |
| aVF_T | A10 | -0.275 | 0.074 | 2.00E-04 | 1.48E-02 |
| aVL_R-area | A10 | 0.318 | 0.081 | 8.12E-05 | 6.01E-03 |
| aVR_T-area | A10 | -0.326 | 0.073 | 8.82E-06 | 6.52E-04 |
| II_ST | A10 | -0.358 | 0.092 | 9.89E-05 | 7.32E-03 |
| II_T | A10 | -0.301 | 0.070 | 1.78E-05 | 1.32E-03 |
| V4_T | A10 | -0.219 | 0.062 | 4.00E-04 | 2.96E-02 |
| V5_T | A10 | -0.232 | 0.061 | 1.00E-04 | 7.40E-03 |
| V6_T | A10 | -0.266 | 0.070 | 2.00E-04 | 1.48E-02 |
| aVF_R | Metformin | -0.267 | 0.075 | 3.00E-04 | 2.22E-02 |
| aVF_T | Metformin | -0.316 | 0.077 | 3.86E-05 | 2.86E-03 |
| aVL_R-area | Metformin | 0.330 | 0.086 | 1.00E-04 | 7.40E-03 |
| aVR_T-area | Metformin | -0.343 | 0.094 | 3.00E-04 | 2.22E-02 |
| II_T | Metformin | -0.323 | 0.084 | 1.00E-04 | 7.40E-03 |
| V4_T | Metformin | -0.226 | 0.066 | 6.00E-04 | 4.44E-02 |

Supplementary Table 12. The cross-trait LDSC estimated the genetic correlation ($r_{g}$) between ETCs and cardiovascular diseases.

| **ETC** | **Phenocode^*^** | **Abbreviation** | **Phenotype** | $\boldsymbol{r}_{\boldsymbol{g}}\boldsymbol{\pm se}$ | ***P value*** |
| --- | --- | --- | --- | --- | --- |
| aVR_T-area | I9_CORATHER | CA | Coronary atherosclerosis | -0.368$\pm$0.095 | 1.00$\times{10}^{-4}$ |
| aVR_T-area | I9_IHD | IHD2 | Ischemic heart disease, wide definition | -0.371$\pm$0.098 | 2.00$\times{10}^{-4}$ |
| aVR_T-area | I9_ISCHHEART | IHD1 | Ischemic heart diseases | -0.382$\pm$0.101 | 2.00$\times{10}^{-4}$ |
| III_S | I9_HEARTFAIL | HF1 | Heart failure, strict | 0.438$\pm$0.116 | 2.00$\times{10}^{-4}$ |
| III_S | I9_HEARTFAIL_ALLCAUSE | HF2 | All-cause Heart Failure | 0.375$\pm$0.096 | 8.73$\times{10}^{-5}$ |
| III_S | I9_HEARTFAIL_NS | HF3 | Heart failure, not strict | 0.390$\pm$0.095 | 3.75$\times{10}^{-6}$ |

*se*: standard error; *FDR*: false discovery rate.

Supplementary Table 13. the significant genetic correlation ($r_{g}$) between medication use and cardiovascular diseases by cross-trait LDSC. *se*: standard error; *FDR*: false discovery rate.

| **CVD GWAS id (phenocode)** | **Medication** | ***r_g_*** | ***se*** | **P-value** | **FDR** | **Bonferroni corrected P-value** |
| --- | --- | --- | --- | --- | --- | --- |
| I9_AF | A10 | 0.2054 | 0.0431 | 1.86E-06 | 5.75E-06 | 1.95E-04 |
| I9_AF_REIMB | A10 | 0.17 | 0.0441 | 1.00E-04 | 2.02E-04 | 0.0105 |
| I9_ANGINA | A10 | 0.3217 | 0.0545 | 3.63E-09 | 2.12E-08 | 3.81E-07 |
| I9_ATHSCLE | A10 | 0.4811 | 0.0729 | 4.20E-11 | 3.39E-10 | 4.41E-09 |
| I9_CHD | A10 | 0.3142 | 0.0542 | 6.82E-09 | 3.58E-08 | 7.16E-07 |
| I9_CHD_NOREV | A10 | 0.3225 | 0.0582 | 2.95E-08 | 1.29E-07 | 3.10E-06 |
| I9_CORATHER | A10 | 0.3503 | 0.0481 | 3.14E-13 | 4.70E-12 | 3.29E-11 |
| I9_CVD | A10 | 0.351 | 0.0474 | 1.29E-13 | 3.32E-12 | 1.36E-11 |
| I9_HEARTFAIL | A10 | 0.4549 | 0.0864 | 1.42E-07 | 4.97E-07 | 1.49E-05 |
| I9_HEARTFAIL_ALLCAUSE | A10 | 0.5539 | 0.0744 | 1.01E-13 | 3.32E-12 | 1.06E-11 |
| I9_HEARTFAIL_NS | A10 | 0.5554 | 0.0739 | 5.80E-14 | 3.04E-12 | 6.09E-12 |
| I9_IHD | A10 | 0.36 | 0.0496 | 3.88E-13 | 5.09E-12 | 4.07E-11 |
| I9_ISCHHEART | A10 | 0.3515 | 0.0498 | 1.71E-12 | 1.80E-11 | 1.80E-10 |
| I9_MI | A10 | 0.3129 | 0.056 | 2.24E-08 | 1.02E-07 | 2.35E-06 |
| I9_MI_STRICT | A10 | 0.2838 | 0.0543 | 1.70E-07 | 5.75E-07 | 1.78E-05 |
| I9_PAD | A10 | 0.5224 | 0.0791 | 4.05E-11 | 3.39E-10 | 4.25E-09 |
| I9_VHD | A10 | 0.2426 | 0.0659 | 2.00E-04 | 3.75E-04 | 0.021 |
| I9_AF | Metformin | 0.2345 | 0.0487 | 1.49E-06 | 4.74E-06 | 1.57E-04 |
| I9_AF_REIMB | Metformin | 0.2065 | 0.0496 | 3.17E-05 | 6.79E-05 | 3.33E-03 |
| I9_ANGINA | Metformin | 0.3339 | 0.0563 | 2.93E-09 | 1.81E-08 | 3.07E-07 |
| I9_ATHSCLE | Metformin | 0.4599 | 0.0743 | 6.18E-10 | 4.06E-09 | 6.49E-08 |
| I9_CHD | Metformin | 0.323 | 0.0561 | 8.64E-09 | 4.32E-08 | 9.07E-07 |
| I9_CHD_NOREV | Metformin | 0.3518 | 0.064 | 3.80E-08 | 1.60E-07 | 3.99E-06 |
| I9_CORATHER | Metformin | 0.3602 | 0.051 | 1.69E-12 | 1.80E-11 | 1.77E-10 |
| I9_CVD | Metformin | 0.4126 | 0.0529 | 6.16E-15 | 6.47E-13 | 6.47E-13 |
| I9_HEARTFAIL | Metformin | 0.5108 | 0.0966 | 1.23E-07 | 4.44E-07 | 1.29E-05 |
| I9_HEARTFAIL_ALLCAUSE | Metformin | 0.5752 | 0.0787 | 2.68E-13 | 4.69E-12 | 2.82E-11 |
| I9_HEARTFAIL_NS | Metformin | 0.5755 | 0.078 | 1.58E-13 | 3.32E-12 | 1.66E-11 |
| I9_IHD | Metformin | 0.3661 | 0.0532 | 5.97E-12 | 5.69E-11 | 6.26E-10 |
| I9_ISCHHEART | Metformin | 0.3564 | 0.0548 | 7.72E-11 | 5.79E-10 | 8.10E-09 |
| I9_MI | Metformin | 0.336 | 0.0578 | 6.00E-09 | 3.31E-08 | 6.30E-07 |
| I9_MI_STRICT | Metformin | 0.3192 | 0.0567 | 1.85E-08 | 8.84E-08 | 1.95E-06 |
| I9_PAD | Metformin | 0.4794 | 0.0771 | 5.06E-10 | 3.54E-09 | 5.31E-08 |
| I9_VHD | Metformin | 0.2853 | 0.0777 | 2.00E-04 | 3.75E-04 | 0.021 |

Supplementary Table 14. The significant causal effect for III_S on heart failures estimated by seven MR methods. $\beta$: causal effect estimate; *se*: standard error; *FDR*: false discovery rate, liab: liability scale.

| **Outcome** | **Exposure** | **Method** | **Nsnp** | $\boldsymbol{\beta}$ | ***se*** | $\boldsymbol{\beta}_{\boldsymbol{liab}}$ | $\boldsymbol{se}_{\boldsymbol{liab}}$ | **P value** | **FDR** |
| --- | --- | --- | --- | --- | --- | --- | --- | --- | --- |
| I9_HEARTFAIL_ALLCAUSE | III_S | MR Egger | 47 | -0.021 | 0.021 | -0.008 | 0.008 | 3.19$\times{10}^{-1}$ | 3.19$\times{10}^{-1}$ |
| I9_HEARTFAIL_ALLCAUSE | III_S | Weighted median | 47 | 0.022 | 0.010 | 0.008 | 0.004 | 3.72$\times{10}^{-2}$ | 6.51$\times{10}^{-2}$ |
| I9_HEARTFAIL_ALLCAUSE | III_S | IVW | 47 | 0.017 | 0.008 | 0.006 | 0.003 | 3.26$\times{10}^{-2}$ | 6.51$\times{10}^{-2}$ |
| I9_HEARTFAIL_ALLCAUSE | III_S | Weighted mode | 47 | 0.020 | 0.016 | 0.008 | 0.006 | 2.17$\times{10}^{-1}$ | 2.53$\times{10}^{-1}$ |
| I9_HEARTFAIL_ALLCAUSE | III_S | GSMR | 47 | 0.017 | 0.007 | 0.006 | 0.003 | 1.54$\times{10}^{-2}$ | 5.38$\times{10}^{-2}$ |
| I9_HEARTFAIL_ALLCAUSE | III_S | MRlap | 47 | 0.035 | 0.018 | 0.013 | 0.007 | 4.65$\times{10}^{-2}$ | 6.51$\times{10}^{-2}$ |
| I9_HEARTFAIL_ALLCAUSE | III_S | CAUSE | 47 | 0.040 | 0.015 | 0.015 | 0.006 | 8.97$\times{10}^{-3}$ | 5.38$\times{10}^{-2}$ |
| I9_HEARTFAIL_NS | III_S | MR Egger | 47 | -0.021 | 0.021 | -0.008 | 0.008 | 3.13$\times{10}^{-1}$ | 3.32$\times{10}^{-1}$ |
| I9_HEARTFAIL_NS | III_S | Weighted median | 47 | 0.019 | 0.010 | 0.007 | 0.004 | 6.58$\times{10}^{-2}$ | 9.22$\times{10}^{-2}$ |
| I9_HEARTFAIL_NS | III_S | IVW | 47 | 0.016 | 0.008 | 0.006 | 0.003 | 3.69$\times{10}^{-2}$ | 8.75$\times{10}^{-2}$ |
| I9_HEARTFAIL_NS | III_S | Weighted mode | 47 | 0.018 | 0.018 | 0.007 | 0.007 | 3.32$\times{10}^{-1}$ | 3.32$\times{10}^{-1}$ |
| I9_HEARTFAIL_NS | III_S | GSMR | 47 | 0.016 | 0.007 | 0.006 | 0.003 | 1.75$\times{10}^{-2}$ | 8.75$\times{10}^{-2}$ |
| I9_HEARTFAIL_NS | III_S | MRlap | 47 | 0.035 | 0.018 | 0.013 | 0.007 | 4.76$\times{10}^{-2}$ | 8.75$\times{10}^{-2}$ |
| I9_HEARTFAIL_NS | III_S | CAUSE | 47 | 0.030 | 0.015 | 0.011 | 0.006 | 5.00$\times{10}^{-2}$ | 8.75$\times{10}^{-2}$ |

Supplementary Table 15. The putative causal effect for A10 and metformin on cardiovascular diseases estimated by seven MR methods.

| **Outcome**  **(phenocode)** | **Exposure** | **Method** | **Nsnp** | $\boldsymbol{\beta}$ | ***se*** | $\boldsymbol{\beta}_{\mathbf{liability}}$ | $\boldsymbol{se}_{\mathbf{liability}}$ | ***P value*** | ***FDR*** |
| --- | --- | --- | --- | --- | --- | --- | --- | --- | --- |
| I9_ANGINA | A10 | CAUSE | 60 | 1.01 | 0.40 | 0.92 | 0.36 | 1.12$\times{10}^{-2}$ | 1.57$\times{10}^{-2}$ |
| I9_ANGINA | A10 | GSMR | 62 | 1.03 | 0.22 | 0.94 | 0.20 | 3.38$\times{10}^{-6}$ | 2.37$\times{10}^{-5}$ |
| I9_ANGINA | A10 | IVW | 63 | 1.08 | 0.30 | 0.98 | 0.28 | 3.59$\times{10}^{-4}$ | 1.26$\times{10}^{-3}$ |
| I9_ANGINA | A10 | MR Egger | 63 | 0.68 | 0.73 | 0.62 | 0.66 | 3.58$\times{10}^{-1}$ | 3.58$\times{10}^{-1}$ |
| I9_ANGINA | A10 | MRlap | 63 | 0.10 | 0.03 | 0.09 | 0.03 | 7.69$\times{10}^{-4}$ | 1.79$\times{10}^{-3}$ |
| I9_ANGINA | A10 | Weighted median | 63 | 1.03 | 0.38 | 0.93 | 0.35 | 7.67$\times{10}^{-3}$ | 1.34$\times{10}^{-2}$ |
| I9_ANGINA | A10 | Weighted mode | 63 | 1.08 | 0.45 | 0.98 | 0.41 | 1.93$\times{10}^{-2}$ | 2.25$\times{10}^{-2}$ |
| I9_ATHSCLE | A10 | CAUSE | 60 | 5.26 | 1.50 | 4.59 | 1.31 | 4.54$\times{10}^{-4}$ | 6.36$\times{10}^{-4}$ |
| I9_ATHSCLE | A10 | GSMR | 62 | 4.66 | 0.78 | 4.07 | 0.68 | 2.37$\times{10}^{-9}$ | 1.66$\times{10}^{-8}$ |
| I9_ATHSCLE | A10 | IVW | 63 | 4.56 | 0.88 | 3.99 | 0.77 | 2.08$\times{10}^{-7}$ | 4.85$\times{10}^{-7}$ |
| I9_ATHSCLE | A10 | MR Egger | 63 | 4.67 | 2.12 | 4.08 | 1.86 | 3.16$\times{10}^{-2}$ | 3.16$\times{10}^{-2}$ |
| I9_ATHSCLE | A10 | MRlap | 63 | 0.13 | 0.02 | 0.11 | 0.02 | 1.83$\times{10}^{-7}$ | 4.85$\times{10}^{-7}$ |
| I9_ATHSCLE | A10 | Weighted median | 63 | 5.21 | 1.24 | 4.55 | 1.09 | 2.76$\times{10}^{-5}$ | 4.83$\times{10}^{-5}$ |
| I9_ATHSCLE | A10 | Weighted mode | 63 | 4.99 | 1.63 | 4.36 | 1.42 | 3.17$\times{10}^{-3}$ | 3.70$\times{10}^{-3}$ |
| I9_CHD | A10 | CAUSE | 60 | 1.08 | 0.38 | 1.14 | 0.41 | 4.77$\times{10}^{-3}$ | 6.68$\times{10}^{-3}$ |
| I9_CHD | A10 | GSMR | 62 | 0.99 | 0.19 | 1.04 | 0.20 | 2.38$\times{10}^{-7}$ | 1.67$\times{10}^{-6}$ |
| I9_CHD | A10 | IVW | 63 | 1.05 | 0.26 | 1.11 | 0.27 | 4.83$\times{10}^{-5}$ | 1.14$\times{10}^{-4}$ |
| I9_CHD | A10 | MR Egger | 63 | 1.17 | 0.62 | 1.24 | 0.66 | 6.53$\times{10}^{-2}$ | 7.10$\times{10}^{-2}$ |
| I9_CHD | A10 | MRlap | 63 | 0.12 | 0.03 | 0.12 | 0.03 | 4.88$\times{10}^{-5}$ | 1.14$\times{10}^{-4}$ |
| I9_CHD | A10 | Weighted median | 63 | 0.93 | 0.32 | 0.98 | 0.34 | 3.44$\times{10}^{-3}$ | 6.02$\times{10}^{-3}$ |
| I9_CHD | A10 | Weighted mode | 63 | 0.81 | 0.44 | 0.85 | 0.46 | 7.10$\times{10}^{-2}$ | 7.10$\times{10}^{-2}$ |
| I9_CHD_NOREV | A10 | CAUSE | 60 | 1.26 | 0.43 | 1.33 | 0.45 | 3.28$\times{10}^{-3}$ | 4.59$\times{10}^{-3}$ |
| I9_CHD_NOREV | A10 | GSMR | 62 | 1.20 | 0.24 | 1.27 | 0.25 | 3.75$\times{10}^{-7}$ | 2.63$\times{10}^{-6}$ |
| I9_CHD_NOREV | A10 | IVW | 63 | 1.27 | 0.30 | 1.35 | 0.32 | 2.71$\times{10}^{-5}$ | 8.10$\times{10}^{-5}$ |
| I9_CHD_NOREV | A10 | MR Egger | 63 | 1.21 | 0.73 | 1.28 | 0.78 | 1.03$\times{10}^{-1}$ | 1.03$\times{10}^{-1}$ |
| I9_CHD_NOREV | A10 | MRlap | 63 | 0.12 | 0.03 | 0.12 | 0.03 | 3.47$\times{10}^{-5}$ | 8.10$\times{10}^{-5}$ |
| I9_CHD_NOREV | A10 | Weighted median | 63 | 1.27 | 0.40 | 1.35 | 0.42 | 1.47$\times{10}^{-3}$ | 2.57$\times{10}^{-3}$ |
| I9_CHD_NOREV | A10 | Weighted mode | 63 | 1.11 | 0.54 | 1.17 | 0.57 | 4.28$\times{10}^{-2}$ | 4.99$\times{10}^{-2}$ |
| I9_CORATHER | A10 | CAUSE | 60 | 1.12 | 0.32 | 0.98 | 0.28 | 4.93$\times{10}^{-4}$ | 6.90$\times{10}^{-4}$ |
| I9_CORATHER | A10 | GSMR | 62 | 0.86 | 0.16 | 0.75 | 0.14 | 1.59$\times{10}^{-7}$ | 1.11$\times{10}^{-6}$ |
| I9_CORATHER | A10 | IVW | 63 | 0.89 | 0.22 | 0.78 | 0.19 | 4.27$\times{10}^{-5}$ | 1.49$\times{10}^{-4}$ |
| I9_CORATHER | A10 | MR Egger | 63 | 0.71 | 0.53 | 0.62 | 0.46 | 1.82$\times{10}^{-1}$ | 1.82$\times{10}^{-1}$ |
| I9_CORATHER | A10 | MRlap | 63 | 0.12 | 0.03 | 0.10 | 0.03 | 6.80$\times{10}^{-5}$ | 1.59$\times{10}^{-4}$ |
| I9_CORATHER | A10 | Weighted median | 63 | 1.00 | 0.29 | 0.88 | 0.25 | 4.67$\times{10}^{-4}$ | 6.90$\times{10}^{-4}$ |
| I9_CORATHER | A10 | Weighted mode | 63 | 0.97 | 0.38 | 0.85 | 0.33 | 1.29$\times{10}^{-2}$ | 1.51$\times{10}^{-2}$ |
| I9_IHD | A10 | CAUSE | 60 | 0.77 | 0.21 | 0.76 | 0.21 | 2.32$\times{10}^{-4}$ | 4.06$\times{10}^{-4}$ |
| I9_IHD | A10 | GSMR | 62 | 0.61 | 0.11 | 0.60 | 0.11 | 6.86$\times{10}^{-8}$ | 4.80$\times{10}^{-7}$ |
| I9_IHD | A10 | IVW | 63 | 0.65 | 0.16 | 0.64 | 0.16 | 3.90$\times{10}^{-5}$ | 1.37$\times{10}^{-4}$ |
| I9_IHD | A10 | MR Egger | 63 | 0.45 | 0.38 | 0.44 | 0.38 | 2.45$\times{10}^{-1}$ | 2.45$\times{10}^{-1}$ |
| I9_IHD | A10 | MRlap | 63 | 0.12 | 0.03 | 0.12 | 0.03 | 7.75$\times{10}^{-5}$ | 1.81$\times{10}^{-4}$ |
| I9_IHD | A10 | Weighted median | 63 | 0.64 | 0.20 | 0.63 | 0.19 | 1.03$\times{10}^{-3}$ | 1.44$\times{10}^{-3}$ |
| I9_IHD | A10 | Weighted mode | 63 | 0.51 | 0.24 | 0.50 | 0.24 | 3.87$\times{10}^{-2}$ | 4.52$\times{10}^{-2}$ |
| I9_ISCHHEART | A10 | CAUSE | 60 | 0.84 | 0.22 | 0.83 | 0.22 | 1.83$\times{10}^{-4}$ | 3.20$\times{10}^{-4}$ |
| I9_ISCHHEART | A10 | GSMR | 62 | 0.69 | 0.12 | 0.68 | 0.12 | 5.14$\times{10}^{-9}$ | 3.60$\times{10}^{-8}$ |
| I9_ISCHHEART | A10 | IVW | 63 | 0.72 | 0.17 | 0.71 | 0.16 | 1.25$\times{10}^{-5}$ | 4.38$\times{10}^{-5}$ |
| I9_ISCHHEART | A10 | MR Egger | 63 | 0.56 | 0.40 | 0.55 | 0.39 | 1.67$\times{10}^{-1}$ | 1.67$\times{10}^{-1}$ |
| I9_ISCHHEART | A10 | MRlap | 63 | 0.13 | 0.03 | 0.13 | 0.03 | 2.35$\times{10}^{-5}$ | 5.48$\times{10}^{-5}$ |
| I9_ISCHHEART | A10 | Weighted median | 63 | 0.67 | 0.21 | 0.66 | 0.21 | 1.61$\times{10}^{-3}$ | 2.25$\times{10}^{-3}$ |
| I9_ISCHHEART | A10 | Weighted mode | 63 | 0.57 | 0.24 | 0.56 | 0.24 | 2.18$\times{10}^{-2}$ | 2.54$\times{10}^{-2}$ |
| I9_MI | A10 | CAUSE | 60 | 1.95 | 0.58 | 1.67 | 0.49 | 7.19$\times{10}^{-4}$ | 1.26$\times{10}^{-3}$ |
| I9_MI | A10 | GSMR | 62 | 1.55 | 0.33 | 1.33 | 0.28 | 2.78$\times{10}^{-6}$ | 1.95$\times{10}^{-5}$ |
| I9_MI | A10 | IVW | 63 | 1.65 | 0.42 | 1.41 | 0.36 | 1.01$\times{10}^{-4}$ | 3.54$\times{10}^{-4}$ |
| I9_MI | A10 | MR Egger | 63 | 1.40 | 1.02 | 1.20 | 0.87 | 1.75$\times{10}^{-1}$ | 1.75$\times{10}^{-1}$ |
| I9_MI | A10 | MRlap | 63 | 0.11 | 0.03 | 0.09 | 0.02 | 2.09$\times{10}^{-4}$ | 4.88$\times{10}^{-4}$ |
| I9_MI | A10 | Weighted median | 63 | 1.76 | 0.61 | 1.50 | 0.52 | 4.11$\times{10}^{-3}$ | 5.75$\times{10}^{-3}$ |
| I9_MI | A10 | Weighted mode | 63 | 1.68 | 0.73 | 1.43 | 0.62 | 2.51$\times{10}^{-2}$ | 2.93$\times{10}^{-2}$ |
| I9_MI_STRICT | A10 | CAUSE | 60 | 1.90 | 0.62 | 1.62 | 0.53 | 2.09$\times{10}^{-3}$ | 3.66$\times{10}^{-3}$ |
| I9_MI_STRICT | A10 | GSMR | 62 | 1.48 | 0.38 | 1.26 | 0.32 | 8.31$\times{10}^{-5}$ | 5.82$\times{10}^{-4}$ |
| I9_MI_STRICT | A10 | IVW | 63 | 1.58 | 0.46 | 1.35 | 0.40 | 6.53$\times{10}^{-4}$ | 2.08$\times{10}^{-3}$ |
| I9_MI_STRICT | A10 | MR Egger | 63 | 1.58 | 1.12 | 1.35 | 0.96 | 1.66$\times{10}^{-1}$ | 1.66$\times{10}^{-1}$ |
| I9_MI_STRICT | A10 | MRlap | 63 | 0.09 | 0.03 | 0.08 | 0.02 | 8.90$\times{10}^{-4}$ | 2.08$\times{10}^{-3}$ |
| I9_MI_STRICT | A10 | Weighted median | 63 | 1.90 | 0.67 | 1.62 | 0.57 | 4.51$\times{10}^{-3}$ | 6.31$\times{10}^{-3}$ |
| I9_MI_STRICT | A10 | Weighted mode | 63 | 1.66 | 0.75 | 1.42 | 0.64 | 3.10$\times{10}^{-2}$ | 3.62$\times{10}^{-2}$ |
| I9_PAD | A10 | CAUSE | 60 | 5.59 | 1.45 | 5.27 | 1.37 | 1.21$\times{10}^{-4}$ | 1.41$\times{10}^{-4}$ |
| I9_PAD | A10 | GSMR | 62 | 5.38 | 0.70 | 5.08 | 0.66 | 1.70$\times{10}^{-14}$ | 1.19$\times{10}^{-13}$ |
| I9_PAD | A10 | IVW | 63 | 5.33 | 0.81 | 5.03 | 0.77 | 5.40$\times{10}^{-11}$ | 1.89$\times{10}^{-10}$ |
| I9_PAD | A10 | MR Egger | 63 | 6.87 | 1.95 | 6.49 | 1.84 | 8.13$\times{10}^{-4}$ | 8.13$\times{10}^{-4}$ |
| I9_PAD | A10 | MRlap | 63 | 0.16 | 0.03 | 0.15 | 0.02 | 2.48$\times{10}^{-10}$ | 5.79$\times{10}^{-10}$ |
| I9_PAD | A10 | Weighted median | 63 | 6.47 | 1.17 | 6.10 | 1.10 | 2.90$\times{10}^{-8}$ | 5.08$\times{10}^{-8}$ |
| I9_PAD | A10 | Weighted mode | 63 | 6.73 | 1.50 | 6.35 | 1.42 | 3.21$\times{10}^{-5}$ | 4.49$\times{10}^{-5}$ |
| I9_ATHSCLE | metformin | CAUSE | 56 | 4.72 | 1.52 | 4.55 | 1.47 | 1.91$\times{10}^{-3}$ | 2.67$\times{10}^{-3}$ |
| I9_ATHSCLE | metformin | GSMR | 52 | 4.69 | 0.76 | 4.53 | 0.73 | 7.11$\times{10}^{-10}$ | 4.98$\times{10}^{-9}$ |
| I9_ATHSCLE | metformin | IVW | 54 | 4.56 | 0.82 | 4.39 | 0.79 | 2.98$\times{10}^{-8}$ | 7.30$\times{10}^{-8}$ |
| I9_ATHSCLE | metformin | MR Egger | 54 | 3.01 | 2.28 | 2.90 | 2.20 | 1.93$\times{10}^{-1}$ | 1.93$\times{10}^{-1}$ |
| I9_ATHSCLE | metformin | MRlap | 54 | 0.18 | 0.03 | 0.17 | 0.03 | 3.13$\times{10}^{-8}$ | 7.30$\times{10}^{-8}$ |
| I9_ATHSCLE | metformin | Weighted median | 54 | 4.55 | 1.17 | 4.39 | 1.13 | 1.07$\times{10}^{-4}$ | 1.87$\times{10}^{-4}$ |
| I9_ATHSCLE | metformin | Weighted mode | 54 | 4.34 | 1.50 | 4.18 | 1.45 | 5.64$\times{10}^{-3}$ | 6.58$\times{10}^{-3}$ |
| I9_CHD | metformin | CAUSE | 56 | 1.14 | 0.36 | 1.33 | 0.42 | 1.65$\times{10}^{-3}$ | 2.32$\times{10}^{-3}$ |
| I9_CHD | metformin | GSMR | 52 | 0.94 | 0.19 | 1.10 | 0.22 | 3.90$\times{10}^{-7}$ | 2.73$\times{10}^{-6}$ |
| I9_CHD | metformin | IVW | 54 | 1.00 | 0.26 | 1.17 | 0.30 | 1.00$\times{10}^{-4}$ | 3.50$\times{10}^{-4}$ |
| I9_CHD | metformin | MR Egger | 54 | 1.53 | 0.71 | 1.79 | 0.84 | 3.67$\times{10}^{-2}$ | 3.67$\times{10}^{-2}$ |
| I9_CHD | metformin | MRlap | 54 | 0.15 | 0.04 | 0.18 | 0.05 | 2.58$\times{10}^{-4}$ | 6.02$\times{10}^{-4}$ |
| I9_CHD | metformin | Weighted median | 54 | 1.04 | 0.33 | 1.21 | 0.39 | 1.66$\times{10}^{-3}$ | 2.32$\times{10}^{-3}$ |
| I9_CHD | metformin | Weighted mode | 54 | 0.90 | 0.39 | 1.05 | 0.45 | 2.38$\times{10}^{-2}$ | 2.78$\times{10}^{-2}$ |
| I9_CHD_NOREV | metformin | CAUSE | 56 | 1.34 | 0.41 | 1.57 | 0.48 | 1.03$\times{10}^{-3}$ | 1.80$\times{10}^{-3}$ |
| I9_CHD_NOREV | metformin | GSMR | 52 | 1.07 | 0.23 | 1.25 | 0.27 | 2.97$\times{10}^{-6}$ | 2.08$\times{10}^{-5}$ |
| I9_CHD_NOREV | metformin | IVW | 54 | 1.15 | 0.29 | 1.34 | 0.34 | 1.01$\times{10}^{-4}$ | 3.54$\times{10}^{-4}$ |
| I9_CHD_NOREV | metformin | MR Egger | 54 | 1.52 | 0.82 | 1.77 | 0.96 | 7.09$\times{10}^{-2}$ | 7.09$\times{10}^{-2}$ |
| I9_CHD_NOREV | metformin | MRlap | 54 | 0.14 | 0.04 | 0.17 | 0.05 | 2.32$\times{10}^{-4}$ | 5.41$\times{10}^{-4}$ |
| I9_CHD_NOREV | metformin | Weighted median | 54 | 1.21 | 0.38 | 1.42 | 0.45 | 1.64$\times{10}^{-3}$ | 2.30$\times{10}^{-3}$ |
| I9_CHD_NOREV | metformin | Weighted mode | 54 | 1.06 | 0.48 | 1.24 | 0.56 | 2.98$\times{10}^{-2}$ | 3.48$\times{10}^{-2}$ |
| I9_CORATHER | metformin | CAUSE | 56 | 0.84 | 0.32 | 0.81 | 0.31 | 7.92$\times{10}^{-3}$ | 1.11$\times{10}^{-2}$ |
| I9_CORATHER | metformin | GSMR | 52 | 0.69 | 0.16 | 0.67 | 0.15 | 1.23$\times{10}^{-5}$ | 8.61$\times{10}^{-5}$ |
| I9_CORATHER | metformin | IVW | 54 | 0.75 | 0.21 | 0.72 | 0.20 | 2.79$\times{10}^{-4}$ | 9.77$\times{10}^{-4}$ |
| I9_CORATHER | metformin | MR Egger | 54 | 1.14 | 0.57 | 1.10 | 0.55 | 5.11$\times{10}^{-2}$ | 5.85$\times{10}^{-2}$ |
| I9_CORATHER | metformin | MRlap | 54 | 0.13 | 0.04 | 0.13 | 0.04 | 6.08$\times{10}^{-4}$ | 1.42$\times{10}^{-3}$ |
| I9_CORATHER | metformin | Weighted median | 54 | 0.88 | 0.28 | 0.85 | 0.27 | 1.87$\times{10}^{-3}$ | 3.27$\times{10}^{-3}$ |
| I9_CORATHER | metformin | Weighted mode | 54 | 0.75 | 0.39 | 0.73 | 0.38 | 5.85$\times{10}^{-2}$ | 5.85$\times{10}^{-2}$ |
| I9_IHD | metformin | CAUSE | 56 | 0.62 | 0.22 | 0.68 | 0.24 | 4.71$\times{10}^{-3}$ | 8.24$\times{10}^{-3}$ |
| I9_IHD | metformin | GSMR | 52 | 0.50 | 0.11 | 0.55 | 0.12 | 4.53$\times{10}^{-6}$ | 3.17$\times{10}^{-5}$ |
| I9_IHD | metformin | IVW | 54 | 0.55 | 0.15 | 0.60 | 0.17 | 3.94$\times{10}^{-4}$ | 1.38$\times{10}^{-3}$ |
| I9_IHD | metformin | MR Egger | 54 | 0.81 | 0.43 | 0.88 | 0.47 | 6.64$\times{10}^{-2}$ | 6.64$\times{10}^{-2}$ |
| I9_IHD | metformin | MRlap | 54 | 0.14 | 0.04 | 0.15 | 0.05 | 9.35$\times{10}^{-4}$ | 2.18$\times{10}^{-3}$ |
| I9_IHD | metformin | Weighted median | 54 | 0.52 | 0.20 | 0.57 | 0.21 | 8.16$\times{10}^{-3}$ | 1.14$\times{10}^{-2}$ |
| I9_IHD | metformin | Weighted mode | 54 | 0.46 | 0.22 | 0.51 | 0.24 | 3.90$\times{10}^{-2}$ | 4.55$\times{10}^{-2}$ |
| I9_ISCHHEART | metformin | CAUSE | 56 | 0.67 | 0.24 | 0.73 | 0.26 | 5.21$\times{10}^{-3}$ | 7.94$\times{10}^{-3}$ |
| I9_ISCHHEART | metformin | GSMR | 52 | 0.57 | 0.11 | 0.62 | 0.12 | 6.92$\times{10}^{-7}$ | 4.84$\times{10}^{-6}$ |
| I9_ISCHHEART | metformin | IVW | 54 | 0.61 | 0.16 | 0.66 | 0.17 | 1.48$\times{10}^{-4}$ | 5.18$\times{10}^{-4}$ |
| I9_ISCHHEART | metformin | MR Egger | 54 | 0.93 | 0.44 | 1.02 | 0.49 | 4.14$\times{10}^{-2}$ | 4.14$\times{10}^{-2}$ |
| I9_ISCHHEART | metformin | MRlap | 54 | 0.15 | 0.04 | 0.16 | 0.05 | 3.32$\times{10}^{-4}$ | 7.75$\times{10}^{-4}$ |
| I9_ISCHHEART | metformin | Weighted median | 54 | 0.57 | 0.21 | 0.62 | 0.22 | 5.67$\times{10}^{-3}$ | 7.94$\times{10}^{-3}$ |
| I9_ISCHHEART | metformin | Weighted mode | 54 | 0.51 | 0.22 | 0.56 | 0.24 | 2.40$\times{10}^{-2}$ | 2.80$\times{10}^{-2}$ |
| I9_MI | metformin | CAUSE | 56 | 1.87 | 0.60 | 1.76 | 0.56 | 1.73$\times{10}^{-3}$ | 3.03$\times{10}^{-3}$ |
| I9_MI | metformin | GSMR | 52 | 1.46 | 0.32 | 1.37 | 0.30 | 6.68$\times{10}^{-6}$ | 4.68$\times{10}^{-5}$ |
| I9_MI | metformin | IVW | 54 | 1.54 | 0.39 | 1.45 | 0.37 | 9.67$\times{10}^{-5}$ | 3.38$\times{10}^{-4}$ |
| I9_MI | metformin | MR Egger | 54 | 1.74 | 1.10 | 1.64 | 1.04 | 1.20$\times{10}^{-1}$ | 1.20$\times{10}^{-1}$ |
| I9_MI | metformin | MRlap | 54 | 0.14 | 0.04 | 0.13 | 0.04 | 1.72$\times{10}^{-4}$ | 4.01$\times{10}^{-4}$ |
| I9_MI | metformin | Weighted median | 54 | 1.51 | 0.54 | 1.42 | 0.51 | 5.40$\times{10}^{-3}$ | 7.56$\times{10}^{-3}$ |
| I9_MI | metformin | Weighted mode | 54 | 1.81 | 0.69 | 1.71 | 0.65 | 1.09$\times{10}^{-2}$ | 1.27$\times{10}^{-2}$ |
| I9_MI_STRICT | metformin | CAUSE | 56 | 1.94 | 0.58 | 1.83 | 0.55 | 8.52$\times{10}^{-4}$ | 1.49$\times{10}^{-3}$ |
| I9_MI_STRICT | metformin | GSMR | 52 | 1.44 | 0.37 | 1.35 | 0.35 | 8.98$\times{10}^{-5}$ | 6.29$\times{10}^{-4}$ |
| I9_MI_STRICT | metformin | IVW | 54 | 1.55 | 0.42 | 1.46 | 0.40 | 2.40$\times{10}^{-4}$ | 8.40$\times{10}^{-4}$ |
| I9_MI_STRICT | metformin | MR Egger | 54 | 1.59 | 1.18 | 1.50 | 1.11 | 1.83$\times{10}^{-1}$ | 1.83$\times{10}^{-1}$ |
| I9_MI_STRICT | metformin | MRlap | 54 | 0.13 | 0.04 | 0.12 | 0.03 | 4.58$\times{10}^{-4}$ | 1.07$\times{10}^{-3}$ |
| I9_MI_STRICT | metformin | Weighted median | 54 | 1.60 | 0.66 | 1.51 | 0.62 | 1.51$\times{10}^{-2}$ | 1.76$\times{10}^{-2}$ |
| I9_MI_STRICT | metformin | Weighted mode | 54 | 1.97 | 0.77 | 1.86 | 0.73 | 1.34$\times{10}^{-2}$ | 1.76$\times{10}^{-2}$ |
| I9_PAD | metformin | CAUSE | 56 | 4.98 | 1.19 | 5.19 | 1.24 | 2.80$\times{10}^{-5}$ | 3.92$\times{10}^{-5}$ |
| I9_PAD | metformin | GSMR | 52 | 4.93 | 0.68 | 5.14 | 0.71 | 4.84$\times{10}^{-13}$ | 3.39$\times{10}^{-12}$ |
| I9_PAD | metformin | IVW | 54 | 4.82 | 0.71 | 5.02 | 0.74 | 9.23$\times{10}^{-12}$ | 2.92$\times{10}^{-11}$ |
| I9_PAD | metformin | MR Egger | 54 | 4.41 | 1.97 | 4.59 | 2.06 | 2.97$\times{10}^{-2}$ | 2.97$\times{10}^{-2}$ |
| I9_PAD | metformin | MRlap | 54 | 0.21 | 0.03 | 0.22 | 0.03 | 1.25$\times{10}^{-11}$ | 2.92$\times{10}^{-11}$ |
| I9_PAD | metformin | Weighted median | 54 | 5.43 | 1.02 | 5.66 | 1.07 | 1.09$\times{10}^{-7}$ | 1.91$\times{10}^{-7}$ |
| I9_PAD | metformin | Weighted mode | 54 | 6.06 | 1.38 | 6.31 | 1.44 | 5.40$\times{10}^{-5}$ | 6.30$\times{10}^{-5}$ |

Phenocode: phenotype code in FinnGen; $\beta$: causal effect estimate; *se*: standard error; *FDR*: false discovery rate; IVW: inverse variance weighted.
